# No safe level of alcohol consumption for brain health: observational cohort study of 25,378 UK Biobank participants

**DOI:** 10.1101/2021.05.10.21256931

**Authors:** Anya Topiwala, Klaus P. Ebmeier, Thomas Maullin-Sapey, Thomas E. Nichols

## Abstract

**Objectives:** To estimate the relationship between moderate alcohol consumption and brain health, determining the threshold intake for harm and identifying whether population subgroups are at differential risk.

**Design:** Observational cohort study. Alcohol consumption was determined at baseline assessment visit using touchscreen questionnaire (2016-10). Multi-modal MRI brain and cognitive testing were performed subsequently (2014-20). Clinical data was extracted from linked Hospital Episode Statistics.

**Setting:** UK Biobank study. Brain imaging was performed on identical scanners with identical protocols at three UK centres (2014-20).

**Participants:** 25,378 participants (mean age 54.9±7.4 years).

**Main outcome measures:** Brain health as defined by structural and functional MRI brain measures.

**Results:** Alcohol consumption was negatively linearly associated with global brain grey matter volume (beta= -0.1, 95%CI= -0.11 to -0.09, p<2×10^−16^). The association with alcohol was stronger than other modifiable factor tested and robust to unobserved confounding. Widespread negative associations were observed with white matter microstructure (beta= -0.08, 95%CI= -0.09 to -0.06, p<2×10^−16^) and positive correlations with functional connectivity. Higher blood pressure and body mass index increased risk of alcohol-related harm (SBP*alcohol: beta= - 0.01, 95%CI = -0.02 to -0.004, p=0.005; BMI*alcohol: beta= -0.01, 95%CI = -0.02 to -0.002, p=0.02). Binging on alcohol had additive negative effects on brain structure on top of the absolute volume consumed (daily compared to never binging: beta= -0.19, 95%CI= -0.30 to -0.08, p<0.01). No evidence was found for differential effects of drinking wine, beer or spirits.

**Conclusions:** No safe dose of alcohol for the brain was found. Moderate consumption is associated with more widespread adverse effects on the brain than previously recognised. Individuals who binge drink or with high blood pressure and BMI may be more susceptible. Detrimental effects of drinking appear to be greater than other modifiable factors. Current ‘low risk’ drinking guidelines should be revisited to take account of brain effects.

**Funding:** AT is supported by a Wellcome Trust fellowship (216462/Z/19/Z). KPE is supported by the UK Medical Research Council (G1001354) and the European Commission (Horizon 2020 732592). This work was also supported by the Li Ka Shing Centre for Health Information and Discovery and NIH grant (TMS, TN: R01EB026859) and a Wellcome Trust award (TN: 100309/Z/12/Z).

## Introduction

Moderate alcohol consumption is common [1] and often viewed as harmless to brain health [2, 3]. Without disease-modifying treatments for neurodegenerative disease, there is a necessary focus on modifiable risk factors such as alcohol. Thus even small adverse effects of moderate drinking on the brain have substantial public health relevance. Differing approaches have been used to ascertain brain health [4]. Advantages of using MRI to determine preservation of optimal brain integrity include the quantitative and sensitive measurement of changes detectable years before clinical symptoms. Finding robust associations with dementia-related imaging phenotypes would provide a biological pathway for a causal link between alcohol and neurodegenerative disease. Alcohol guidelines could be amended to reflect evidence about brain health rather than rely solely on that from current cardiovascular disease and cancer research as currently. There is a pressing need for stratified public health, with personalised risk predictions that would allow focused interventions to those at greatest need [5].

Whilst chronic heavy alcohol intake damages brain health [6, 7], the impact of ‘moderate’ consumption [defined variably] continues to be contentious. Epidemiological studies have produced conflicting results [8, 9]. A J-shaped curve has been suggested to link alcohol to brain outcomes. Our previous work found novel associations between moderate consumption of alcohol and multiple adverse brain outcomes [10]. Recent studies have supported these findings but were unable to fully examine the shape of the relationship, accounted for few potential confounders, or examined selected regions of interest rather than the whole brain [11, 12]. Further unanswered questions remain. The threshold of alcohol intake necessary for brain harm is unknown. The impact of moderate drinking on brain connectivity is unclear. Also uncertain is whether specific population subgroups are at particularly increased risk. The effect of alcohol on cardiovascular outcomes is reported to vary according to sex and age [13]. Whether age and sex affect the risk of brain harm remains unclear. Understanding how medical comorbidities, such as hypertension and liver disease, interplay is also limited. Whilst blood pressure and BMI are linked to brain harm, any interaction with alcohol is undetermined [14, 15]. ApoE4 genotype is a well-established risk factor for Alzheimer’s disease [16], but it is unknown whether alcohol intake interacts. There are little substantiated claims that red wine has beneficial effects due to its concentration of polyphenols (in particular resveratrol) [17]. Conversely it is thought that certain drinking patterns, such as binging, may worsen the impact of drinking on the brain [18]. Imaging studies to date have been highly selected with insufficient power to answer these questions.

For the first time, in UK Biobank, the world’s largest imaging sample, we have a sufficiently large sample to clarify these important public health issues. We estimated the relationship between alcohol consumption and brain health. Furthermore, we investigated whether certain population subgroups, defined by sociodemographic, clinical and drinking factors, demonstrated increased susceptibility to alcohol-related brain effects. Our hypotheses were as follows: 1) the threshold intake for adverse brain outcomes is lower than currently defined as “low risk” drinking (<14 units weekly); 2) older age, female sex, vascular risk, liver dysfunction and ApoE4 genotype increase the risk of harm; 3) binge-drinking is associated with worse outcomes; 4) type of beverage per se has little impact on outcomes.

## Methods

### Sample

UKB is the world’s largest neuroimaging resource, with over 40,000 subjects imaged among the ∼500,000 adults of the core study (aged 40-69 years at initial recruitment in 2006-10). Data used in this study include: clinical data (alcohol consumption and confounders), linked Hospital Episode Statistics (electronic health records with clinician-coded diagnoses), brain imaging, cognitive testing and genotyping. Subjects with at least one brain MRI by 28.1.21 (n=43,572) were included in the study. Exclusions were due to missing imaging, alcohol or confounder data, or those with images of insufficient quality for analyses (supplementary Figure 1).

### Data

Tabular variables were extracted from UKB files using FSL’s funpack [19].

#### Alcohol consumption measures

Subjects were asked at baseline their alcohol intake in an average week for those drinking at least weekly, and average monthly intake for those drinking less frequently. Numbers of glasses were asked for, and subjects were given guidance about how many glasses in the normal bottle. Glasses were converted to UK units as follows [20]: red or white wine = 1.7 units; fortified wine=1.2 units; pint = 2.4 units; spirits = 1 unit; other (e.g. alcopops) =1.2 units. Amounts were also converted to grams pure ethanol (1 UK unit=8g) to aid international comprehension and comparison. For monthly intake, units were divided by 4.3 to estimate a weekly amount. Amounts were summed across beverage types and weekly and monthly intakes to generate a total weekly alcohol unit intake used for further analyses. This weekly total was additionally divided into quantiles for selected analyses. Subjects who self-reported as “drinkers” but then reported 0 units weekly (n=3760) were excluded from the analyses to avoid misclassification. Non-drinkers were subdivided into former and never drinkers based on a separate alcohol status question. Current drinkers reporting solely drinking wine, beer, or spirits (rather than a combination) were also sub-classified by the beverage type they consumed. Subjects who had data on binge drinking (defined as greater than six units of alcohol consumed in one episode) frequency at baseline assessment were included in a pre-specified sensitivity analysis to assess whether binging frequency, independently of alcohol volume, was associated with brain structure. Those who reported being current non-drinkers but reported any frequency of binge drinking (n=22), and lowest quartile drinkers who reported daily binging (n=5), were assumed to have missing data and were excluded from the analysis.

#### Medical diagnoses

Selected medical conditions (heart disease, liver disease, depressive disorder, alcohol dependence) were defined using ICD 9 & 10 diagnoses in linked Hospital Episode Statistics (HES). Primary care records were not used as only half the UKB sample has linked records thus far. Specific diagnosis codes used and numbers of subjects with such codes are available in the supplementary materials. Diabetes diagnoses were generated by a UKB algorithm using self-report, hospital care records, and death certificates. Subtypes of diabetes mellitus (insulin-dependent, noninsulin-dependent, unspecified were combined to generate a binary variable (diabetes present/absent). Depression cases were defined using ICD 9 & 10 codes for single or recurrent episodes of at least moderate severity.

#### Cognitive measures

Cognitive function was assessed at the time of imaging, for a subset of the sample, using the following tests:

trail-making (numerical – ‘a’, and alpha-numeric – ‘b’), tower rearranging, digit span, digit substitution, pairs matching, matrix pattern completion, paired association.

#### Genetics

Genotypes for two SNPs of interest – ApoE4 (rs429358 & rs7412) were extracted from v3 imputed genotype data for UKB subjects using qctool (version 2.0.7). ApoE4 is the strongest genetic risk factor for late-onset dementia [21]. Subjects were classified according to their number of E4 alleles (0-2) for ApoE.

#### Other variables

Covariates were chosen because of associations with brain imaging phenotypes in the literature [22-25]. On this basis they were included as potential confounders. Baseline data on age, sex, smoking status, educational qualifications, systolic (SBP) and diastolic (DBP) blood pressure, body mass index (BMI), Townsend Deprivation Index (TDI) and weekly exercise (MET minutes) were used. Gamma glutamyl transferase (GGT) and cholesterol levels were derived from a blood sample at baseline. In all analyses imaging-derived confounders included imaging site and head size (T1 scaling factor). In sensitivity analyses the following additional imaging-related factors were included: head motion (structural images), table positioning, acquisition parameters (software version, head coil replacement, cold head replacement, service pack, minor protocol changes, other hardware events).

#### MRI acquisition and pre-processing

Participants were scanned at three imaging centres with identical Siemens Skyra 3T scanners (software VD13) using a standard 32-channel head coil. T1-weighted structural images, diffusion tensor and resting state functional images were utilized in this study. Full details of the image pre-processing and quality control pipeline are described in supplementary methods [26].

#### Image-derived phenotypes (IDPs)

IDPs were pre-specified on the basis of their previous association with alcohol [10] and their relation to clinical phenotypes: 1) hippocampal volume (left and right, determined by FIRST [27]), 2) corpus callosum fractional anisotropy (a tract-averaged marker of white matter microstructural integrity), 3) default mode network functional connectivity (determined by resting state functional MRI). Hippocampal atrophy is a validated biomarker of Alzheimer’s disease [28] and has been previously associated with alcohol [10]. Corpus callosum fractional anisotropy is a marker of white matter integrity and vascular damage, linked to vascular dementia [29]. Multiple changes of DMN connectivity have been reported in dementia [30].

Functional connectivity of the brain at rest was determined using node amplitudes (standard deviation of time courses; n=21 deemed “good” rather than noise), and ‘edges’ (partial correlations between nodes from rfMRI netmats; n=210) from d25 components were extracted from IDPs processed by the UKB imaging team. The following additional IDPs were used in post-hoc sensitivity analyses: lingual gyrus grey matter volume (from FAST [31], to explore voxel-based morphometry results, right and left thalamus and putamen volumes (extracted using FIRST), and additional metrics from diffusion tensor imaging including NODDI [32](mean diffusivity, radial diffusivity, mode, intra- and extra-cellular volume fractions, neurite orientation).

### MRI analyses

#### Grey matter

Relationships between alcohol use and grey matter were examined in a brain-wide hypothesis-free manner using voxel-based morphometry, an objective method to compare grey matter density between individuals in each voxel (smallest distinguishable image volume) of the structural image. Structural data were analysed with FSL-VBM [33](http://fsl.fmrib.ox.ac.uk/fsl/fslwiki/FSLVBM), an optimised VBM protocol [34] carried out with FSL tools [35]. First, structural images were brain-extracted and grey matter-segmented before being registered to the MNI 152 standard space using non-linear registration [36]. The resulting images were averaged and flipped along the x-axis to create a left-right symmetric, study-specific grey matter template. Second, all native grey matter images were non-linearly registered to this study-specific template and “modulated” to correct for local expansion (or contraction) due to the non-linear component of the spatial transformation. The modulated grey matter images were then smoothed with an isotropic Gaussian kernel with a sigma of 2 mm.

Alcohol intake and covariates were demeaned (to avoid the mean signal being shared amongst many covariates) for the design matrix. An explicit grey matter mask was created by thresholding (at 0.01) a mean image of unsmoothed T1 images for included subjects.

#### White matter microstructure

Voxelwise statistical analysis of fractional anisotropy (FA), axial diffusivity (L1), radial diffusivity (L2, L3), mean diffusivity (MD) and mode (MO) data was carried out using Tract-Based Spatial Statistics (TBSS) [37]. This involves non-linear registration followed by projection onto an alignment-invariant tract representation (the “mean FA skeleton”). This avoids alignment problems for multiple subjects and avoids arbitrariness of spatial smoothing extent, improving the sensitivity, objectivity and interpretability of analysis of multi-subject diffusion imaging studies [38]. Multiple diffusion indices were analysed to allow a richer investigation of localised connectivity related changes.

Diffusion images were corrected for head movement and eddy currents (eddy_correct) and brain masks generated using BET. Fractional anisotropy, mean diffusivity, axial diffusivity and radial diffusivity maps were generated using DTIFit (http://fsl.fmrib.ox.ac.uk/fsl/fdt) that fits a diffusion tensor model at each voxel. Tract-based spatial statistics (TBSS) were used in a 4-stage process. Pre-processing prepared images for registration to standard space. Mean FA, L2, L3, MD, MO, and corresponding skeletonized images were created, and thresholded. Lastly each image was projected onto the relevant skeleton. A sensitivity analyses excluded non-drinkers.

### Statistical analyses

The sample was characterized using means and standard deviations for continuous variables, numbers and percentages for categorical variables, split by current alcohol intake status. We examined differences in sociodemographic and clinical factors between participants drinking category using one-way ANOVA (continuous variables) or χ^2^ tests of independence (categorical variables). Alcohol intake for the sample was visualized using cumulative distribution plots, separately by sex. These processes were repeated for sub-analyses by beverage type consumption.

#### Mass univariate analyses

The Big Linear Model toolbox [39] was used to perform mass univariate OLS regression (parametric inference) voxelwise. A missingness threshold of 80% was employed (i.e. voxels with recorded data for less than 80% of subjects were discarded from the analysis), and two T contrasts (positive and negative correlation with alcohol) and an F contrast were computed. A p-value threshold that capped the False Discovery Rate (FDR) at 0.05 was generated using FSL’s FDR (https://fsl.fmrib.ox.ac.uk/fsl/fslwiki/FDR). This was used to threshold T statistic images. Sensitivity analyses excluded non-drinkers.

#### Univariate analyses

Regression models were fitted with the IDPs as dependent variables and alcohol intake as an independent variable. All variables were standardized (z scores) for comparability of estimates. Alcohol was fitted using linear and non-linear models. The latter comprised restricted cubic splines (RCS) being applied to alcohol intake (R package rms v.6.2-0). The latter models the effect of alcohol parametrized with a cubic spline with 5 knots at 5^th^, 25^th^, 50^th^, 75^th^ and 95^th^ percentiles (0.2, 1.9, 10.1, 20.3, 49.2 units) with the tails restricted to linearity (for stability). The knot number and position used are recommended in the literature and were empirically tested for this data using AIC criteria [40]. A spline model avoids the loss of power and arbitrary cut-points of categorization, and fits threshold effects better than polynomials. It offers a flexible approach to estimate the shape of the exposure-outcome curve which was of key interest in this study. Non-linearity was formally tested (H0:β2=β3=…=βk-1= 0) with an F-test. The same confounders were included in all analyses. Polynomic terms for age (age^2^ and age^3^), and age x sex, age^2^ x sex interactions were also included [41]. Alcohol can impact blood pressure and cholesterol, and therefore to investigate the possibility that these covariates could be mediators rather than confounders in the alcohol-brain pathway, we examined regression models with and without these covariates. Interactions between alcohol intake (weekly units, continuous) and age, sex, diagnosis of heart disease, blood pressure, BMI and ApoE4 genotype were also tested. Given how widespread the associations with alcohol were, total grey matter volume (normalized to head size) was used as the dependent variable in these analyses. There were insufficient numbers of subjects with liver disease or depressive disorder diagnoses to test interactions (see supplementary materials).

We performed a sensitivity analysis to test how robust associations were to unobserved confounding, a concern in all observational studies. Partial R^2^ and robustness values were calculated using R’s sensemakr package, which estimate the necessary strength of an unobserved confounder required to fully account for the alcohol effect on brain health [42]. The impact of an unobserved confounder depends on two measures: its association with brain health (the outcome) and its association with alcohol (the exposure of interest). The strength of these associations can be measured in partial R2. The robustness value is the level of outcome and exposure confounding (assumed to be equal for these purposes) required to zero-out the alcohol-brain health association if we actually could control for the unobserved confound. Plausible outcome and exposure confounding partial R2’s can be computed for known confounds. This allows the impact of an unobserved confound to be calibrated by the severity of the strongest existing confounds. We used age, sex and smoking (the strongest known confounders for grey matter volume and alcohol intake) for this purpose.

In a sub-analysis, regression models were re-run amongst three separate groups of drinkers consuming solely wine, beer, or spirits. The association of binge drinking frequency (distinct from total alcohol intake volume) on grey matter volume was examined, by adding binging frequency as a categorical variable in the regression model, whilst controlling for alcohol units consumed weekly as a linear covariate. Associations between resting fMRI connectivity (node amplitude) and cognitive test performance at the time of scanning were examined using regression models, which included the same covariates as previously.

Standardised regression coefficients (generated by converting outcomes to z scores) for were plotted using R’s jtools package (v2.1.3). Spline models were plotted graphically using R’s predict function to show how each brain IDP (z-score) would be expected to change with alcohol consumption, keeping other independent variables at a fixed level. Manhattan plots (implemented in R’s qqman package v0.1.8) were used to display associations between ‘edges’ (functional connectivity between nodes) and alcohol intake. All analyses were completed in R (v3.6.0) unless otherwise stated.

#### Patient involvement

Participants are from the UK Biobank study. No patients were involved in setting the research question or the outcome measures, nor were they involved in the design, recruitment to, or conduct of the study. No patients were asked to advise on interpretation or writing up of results. Participants are thanked in the acknowledgements.

## Results

Baseline characteristics are given in Table 1. Alcohol consumption was very common amongst the sample. A small minority were non-drinkers (5.2%), who could be divided approximately equally into never drinkers (52.8%) and ex-drinkers (47.2%; for cumulative distribution see supplementary Figure 2). The non-drinking groups comprised a higher percentage of females, reported lower rates of smoking, higher indices of deprivation, and lower educational qualifications than drinkers. Current drinkers had higher systolic and diastolic blood pressure and HDL levels but lower total cholesterol and BMI than non-drinkers.

**Table 1:** Baseline characteristics for included sample (n=25,377*) by drinking status.

| Baseline characteristic | Never drinkers<br>(N=691) | Former drinkers<br>(N=617) | Current drinkers<br>(N=24,069) | Group differences<sup>3</sup> |
| --- | --- | --- | --- | --- |
| **Age band, N(%)** |  |  |  |  |
| 40- <50 years | 206<br>(29.8) | 171<br>(27.7) | 6083 (25.3) | X<sup>2</sup>=16.1,<br>df=4,p=0.003 |
| 50- <60 years | 215<br>(31.1) | 241<br>(39.1) | 8892 (36.9) |  |
| 60- <70 years | 200<br>(28.9) | 153<br>(24.8) | 6756 (28.1) |  |
| **Sex, female N(%)** | 457<br>(66.1) | 320<br>(51.9) | 11,477<br>(47.7) | X<sup>2</sup>=94.8,<br>df=2,p<2.2x10<sup>-16</sup> |
| **Smoking status, N(%)** |  |  |  |  |
| *Never* | 603<br>(87.3) | 315<br>(51.1) | 14,119<br>(58.7) | X<sup>2</sup>=250.5, df=4,<br>p<2.2x10<sup>-16</sup> |
| *Previous* | 61 (8.8) | 253<br>(41.0) | 8428 (35.0) |  |
| *Current* | 27 (3.9) | 49 (2.9) | 1522 (6.3) |  |
| **Qualifications<sup>4</sup>, N(%)** |  |  |  |  |
| *None* | 66<br>(9.6) | 54<br>(8.8) | 1264 (5.3) | X<sup>2</sup>=55.9, df=12,<br>p=1.24x10<sup>-7</sup> |
| *A level* | 86 (12.5) | 87 (14.1) | 3145 (13.1) |  |
| *Degree* | 301<br>(43.6) | 263<br>(42.6) | 11,813<br>(49.1) |  |
| **Systolic BP<sup>2</sup>, mmHg** | 134.28<br>(18.8) | 134.02<br>(17.8) | 137.3 (18.7) | F(2,25374)=17.4,<br>p=2.9x10<sup>-8</sup> |
| **Diastolic BP<sup>2</sup>, mmHg** | 79.7<br>(10.8) | 79.7<br>(10.2) | 81.7 (10.5) | F(2,25374)=21.5,<br>p=4.9x10<sup>-10</sup> |
| **Body Mass Index<sup>2</sup>, kg/m<sup>2</sup>** | 26.7<br>(4.7) | 27.0<br>(4.7) | 26.5 (4.0) | F(2,25374)=5.7,<br>p=0.003 |
| **Total cholesterol<sup>2</sup>, mmol/L** | 5.6 (1.2) | 5.54<br>(1.2) | 5.73 (1.1) | F(2,25374)=13.1,<br>p=2.2x10<sup>-6</sup> |
| **Non-high Density cholesterol<sup>2</sup>, mmol/L** | 4.2 (1.0) | 4.2 (1.1) | 4.3(1.1) | F(2,25374)=0.001,<br>p=1.0 |
| **Diabetes Mellitus<sup>5</sup>, N(%)** | 57<br>(8.3) | 40<br>(6.5) | 1139 (4.7) | X<sup>2</sup>=21.5, df=2,<br>p=2.2x10<sup>-5</sup> |
| **Townsend Deprivation Index<sup>2</sup>** | -1.4 (3.0) | -0.8 (3.2) | -2.0 (2.6) | F(2,25374)=72.7,<br>p=2.0x10<sup>-16</sup> |
| **Exercise<sup>2</sup>, minutes weekly<sup>1</sup>** | 120.8<br>(103.2) | 125.7<br>(93.8) | 121.8 (90.6) | F(2,25374)=0.6,<br>p=0.5 |

<sup>1</sup> Minutes of walking, moderate or vigorous activity. Derived from Metabolic Equivalent Task (MET) based on International Physical Activity Questionnaire guidelines.
<sup>2</sup> Mean (standard deviation).
<sup>3</sup> One-way ANOVA for continuous variables, chi-square test for categorical variables.
<sup>4</sup> Only selected qualification categories are presented for brevity.
<sup>5</sup> Defined using self-report, hospital care records, and death certificates.
\* N=1 subject included in the VBM analysis was excluded here due absence of alcohol status data.

Median alcohol intake was 13.5 units (102g) weekly (IQR=17.3). Almost half the sample (48.2%) were drinking above current UK ‘low risk’ guidelines (14 units (112g) weekly), but few subjects drank very heavily (>50 units (400g) weekly). 31 subjects had an ICD 9 or 10 diagnosis of alcohol dependence in their linked HES records, although n=403 self-reported having previously been addicted to alcohol at the UKB baseline questionnaire. More frequent binge drinking was associated with younger age, male sex, more educational qualifications, higher deprivation score, and current smoking, independent of total alcohol consumed weekly (supplementary Table 1). Of those who reported drinking solely one type of alcoholic beverage (n=6602), most drank wine (76.9%) (supplementary Figure 2). Wine drinkers were significantly older, better educated, had lower BMI, deprivation, and smoking levels (see supplementary Table 2).

### Alcohol and grey matter

Higher volume of alcohol consumption per week was associated with lower grey matter density almost globally (Figure 1). These findings were adjusted for all known potential confounders and multiple comparisons. Results were unchanged in a sensitivity analyses (n=22,536 subjects) additionally adjusted for a further seven MRI acquisition, four table position parameters and head motion during the structural scan [41] (supplementary Figure 4). Similarly, excluding non-drinkers from the sample did not change the findings (supplementary Figure 5).

**Figure 1:**
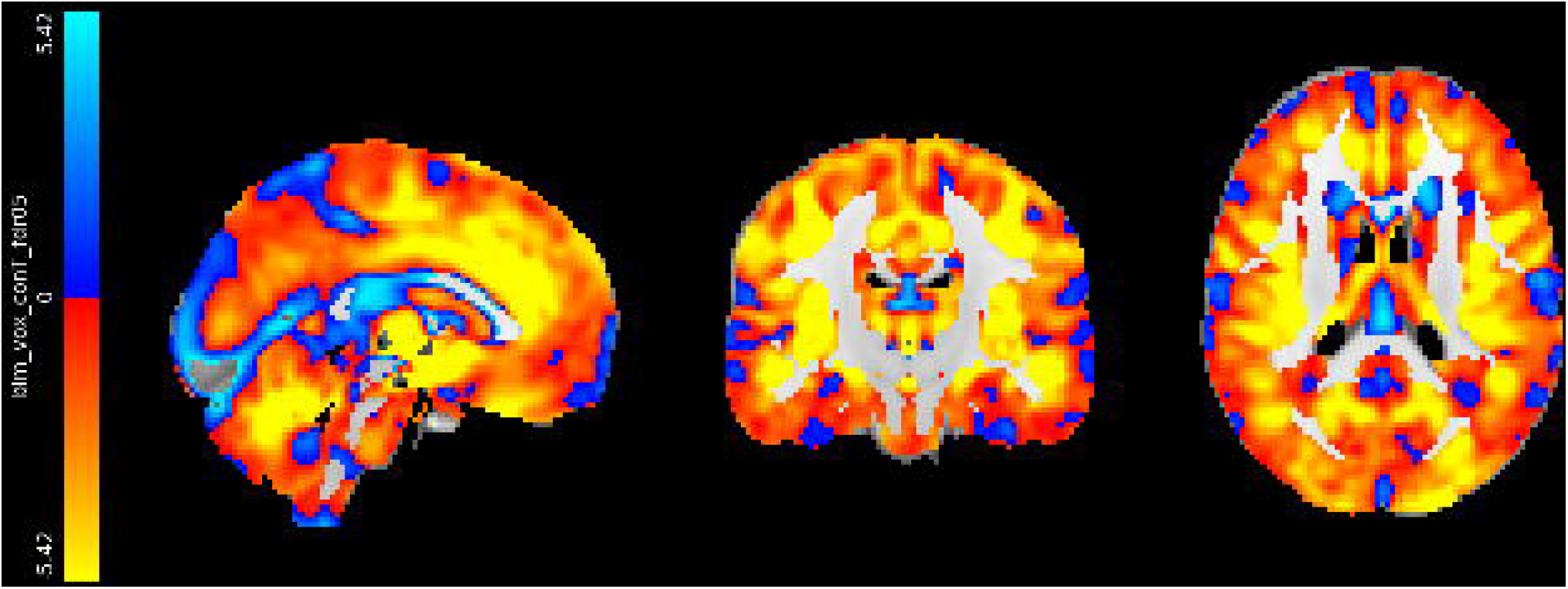
**Association between weekly alcohol intake and grey matter density generated using voxel-based morphometry. Colours show FDR-adjusted T statistics from regression models with alcohol as independent variable. Adjusted for: age, sex, age^2^, age^3^, age x sex, age^2^ x sex, imaging site, SBP, DBP, cholesterol, HDL, Diabetes Mellitus, smoking, BMI, exercise, TDI, depression, qualifications, head size. N= 25,378. Voxel location: 45, 54, 45. Coordinates (anatomically aligned): 9.9×10^−5^, -18.0, 18.0.**

Alcohol explained up to 0.8% of grey matter volume variance (change in adjusted R^2^). While this is a small effect size (supplementary Figure 6) in comparison to age (R^2^=27%), alcohol made a larger contribution than any other modifiable risk factor tested, including smoking (supplementary Table 3). Removing blood pressure and cholesterol from the models made no difference to estimates for alcohol (supplementary Table 5). In a sensitivity analysis, we estimated that to nullify the estimate of alcohol, an unobserved confounder would need to explain both >12% of grey matter and >12% of alcohol intake variation (robustness value=0.12). The presence of an unobserved confounder which achieves this seems implausible given examination of the strongest known confounders. For example, whilst age explains 54% of grey matter, it only explains 0.1% alcohol intake, and an unobserved confounder with such characteristics would be unable to explain-away the alcohol-brain relationship (supplementary Figure 7). Similarly sex and smoking each explain only 0.2% of grey matter and 4% of alcohol intake.

The VBM analysis highlighted some voxels with positive associations with alcohol consumption, but these were not replicated in post-hoc region-of-interest analyses, so are likely artefactual. The spline model did not offer significant improved fit over a model with a linear effect of alcohol (grey matter volume: df=3, F=1.6, p=0.2) (Figure 2). The positive slopes between 0-5 alcohol units in Figure 2 for several subcortical regions flattened upon excluding previous drinkers (supplementary Figure 8) suggesting a “sick quitter” effect i.e. previous drinkers may have other health concerns or risk factors for poorer brain health. Of the subcortical regions tested, the strongest associations with alcohol consumption were found with bilateral thalami volume.

**Figure 2:**
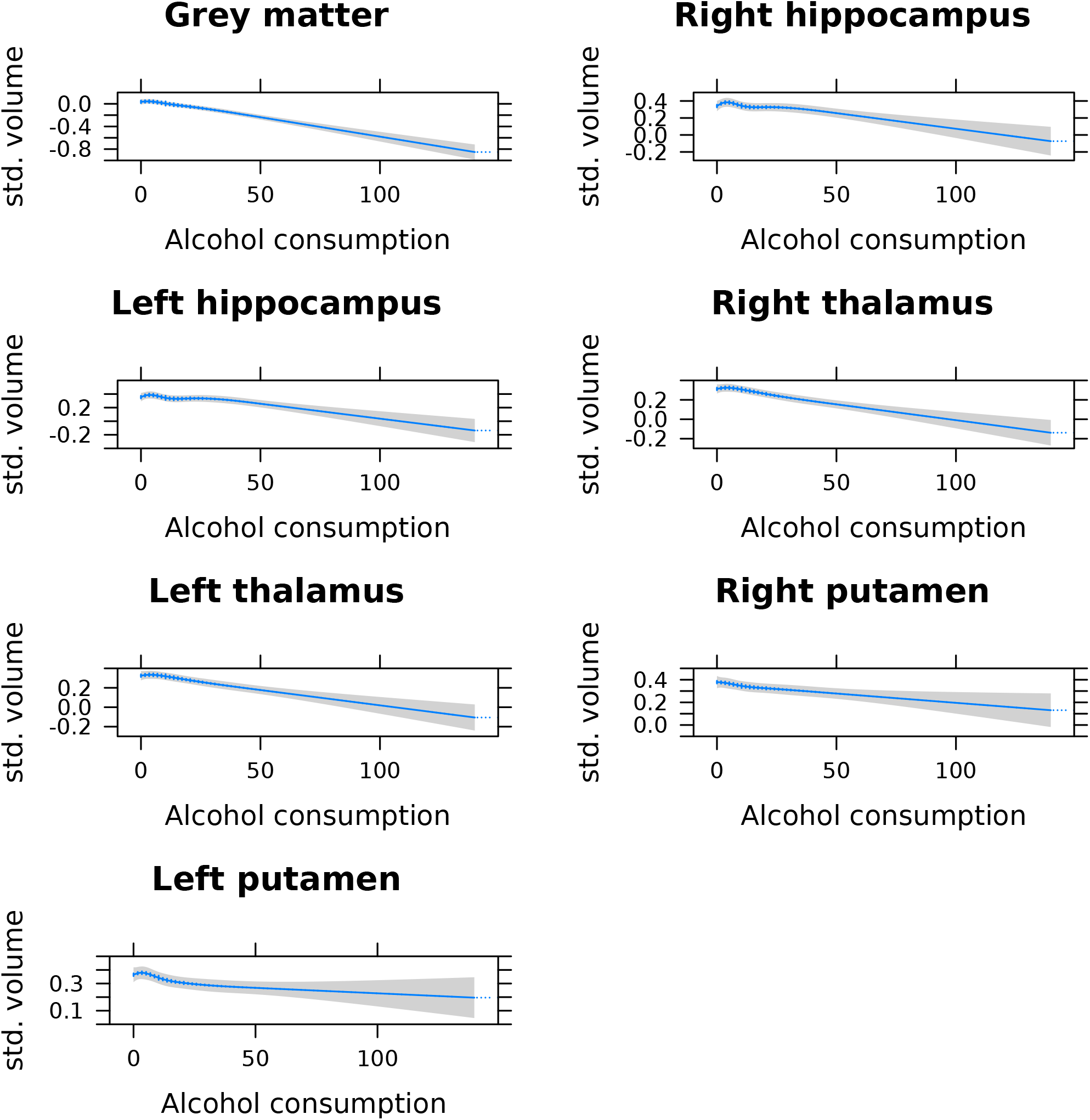
**Predicted change in selected brain volumes according to weekly alcohol intake in units (grams conversion: 50u=400g, 100u=800g). Predictions are based on regression models with alcohol (spline fit with knots at 5^th^, 25^th^, 50^th^, 75^th^ and 95^th^ percentiles) as an independent variable and standardized brain volume as the dependent variable with n=22,254 subjects. Models are adjusted for: age, sex, age^2^, age^3^, age x sex, age^2^ x sex, SBP, DBP, TDI, smoking, BMI, non-HDL cholesterol, Diabetes Mellitus, head size, exercise. 95% confidence intervals are shaded.**

### Alcohol and white matter

Widespread negative associations were also found between alcohol consumption and all the white matter integrity measures assessed (Figure 3 & supplementary Figures 9-13). Results were unchanged when additional image-related confounders were adjusted for (supplementary Figure 14) or only current drinkers included (supplementary Figure 15). A particularly vulnerable region appeared to be the anterior corpus callosum (genu). In a post-hoc analysis, the DTI metrics which were most tightly coupled to alcohol intake were measures of intra-cellular and extra-cellular water volume (isovf and icvf), markers of neurite density (supplementary Figure 9). Again, spline models provided no better fit for associations with white matter microstructure than linear models.

**Figure 3:**
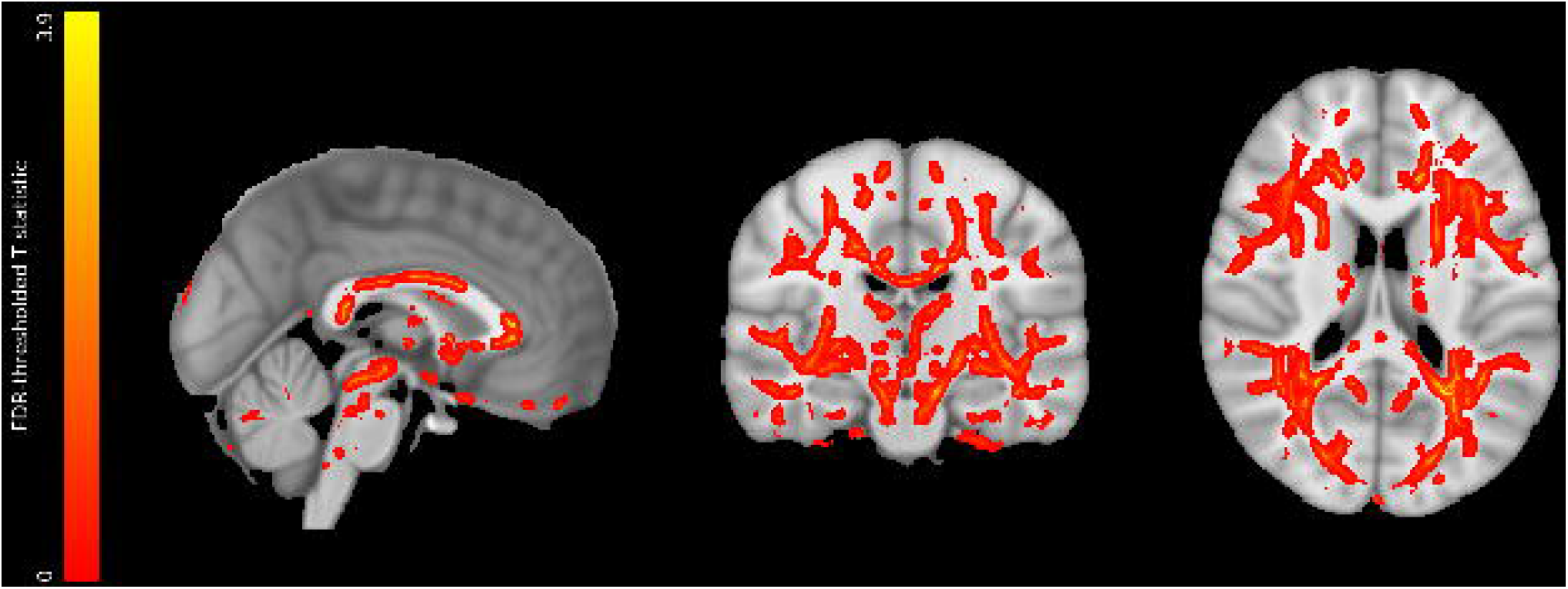
**Negative associations between weekly alcohol intake and fractional anisotropy – a diffusion tensor imaging measure of white matter integrity. Red-yellow voxels indicate FDR-thresholded T statistics. Adjusted for: age, sex, age^2^, age^3^, age x sex, age^2^ x sex, imaging site, SBP, DBP, cholesterol, HDL, Diabetes Mellitus, smoking, BMI, exercise, TDI. N=24,030. Voxel location: 90, 108, 90. Anatomical coordinates: -0.5, -17.5, 18.5.**

### Population subgroups at higher risk

High blood pressure and BMI steepened the negative association between alcohol and brain health. This was determined by significant interactions with alcohol intake in predicting grey matter volume (SBP*alcohol: beta= -0.01, 95% CI = -0.02 to -0.004, p=0.005; DBP*alcohol: beta= -0.01, 95%CI = -0.02 to -0.004, p=0.006; BMI*alcohol: beta= -0.01, 95% CI = -0.02 to -0.002, p=0.02) (Figure 4). In contrast, there were no significant interactions between alcohol and age, sex, GGT, heart disease or ApoE4 genotype that would survive multiple testing correction.

**Figure 4:**
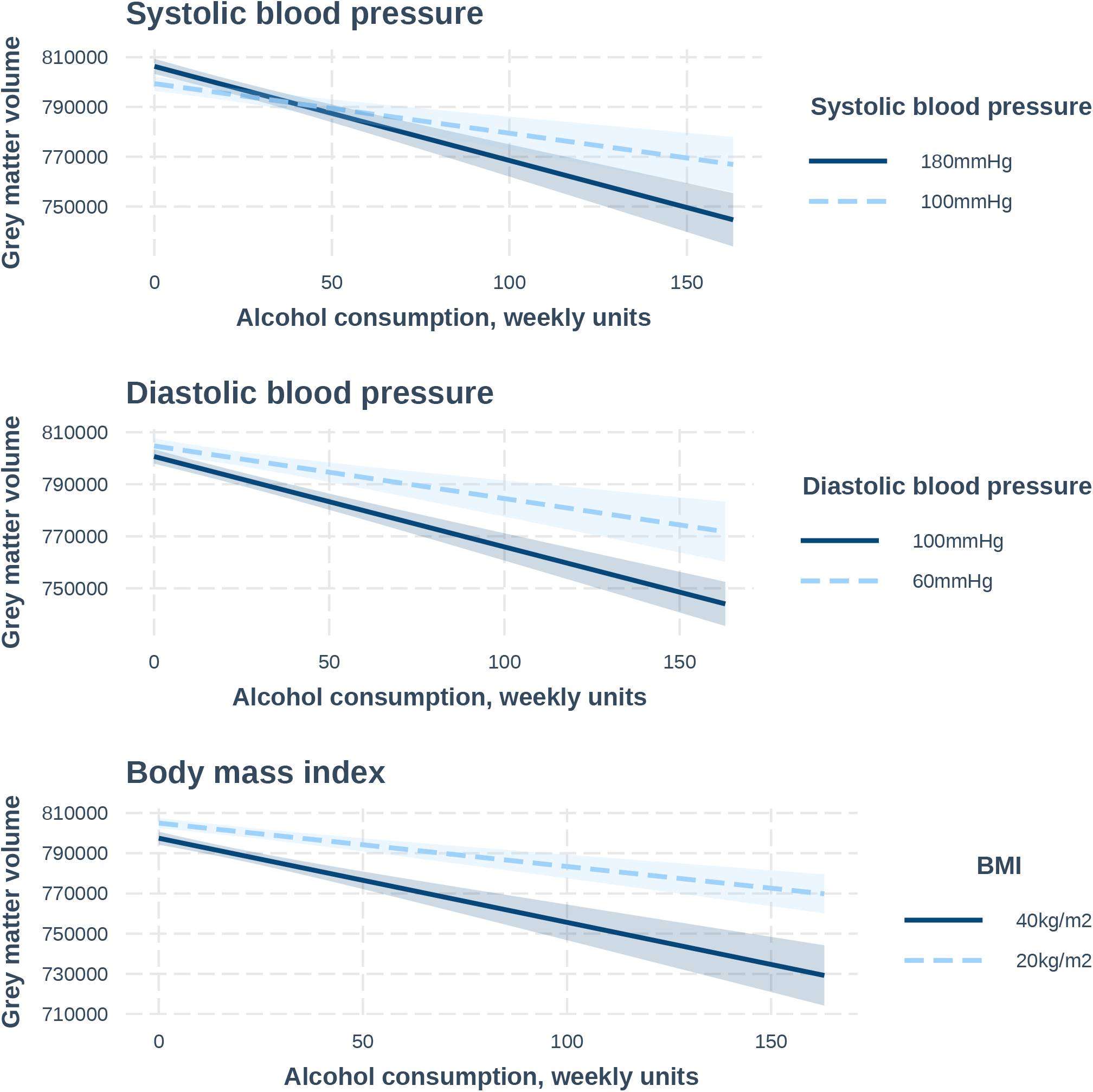
**Predicted change in grey matter volume according to alcohol intake at different blood pressure and BMI levels, when all other confounders are held constant. Predictions generated based on regression models with n=19,617 subjects adjusted for: age, age^2^, age^3,^ sex, age x sex, age^2^ x sex, Diabetes Mellitus, SBP, DBP, BMI, cholesterol, HDL, imaging site, exercise, smoking status, qualifications, TDI, head size. 95% confidence intervals are shaded.**

Those binging daily had a significantly lower total grey matter volume than never bingers, even after controlling for total alcohol volume consumed weekly (Figure 5). The impact of binging was apparent in those drinking greater than 18 units (144g) weekly (quantiles 4 & 5). Amongst those subjects drinking 18-28 units (144-224g) weekly (quartile 4), those binging at frequencies of less than monthly, monthly, or daily had significantly lower grey matter density compared to never bingers. Amongst the highest-level drinkers (quantile 5) only daily bingers had less grey matter than never bingers. Interactions between alcohol intake in units and binging frequency were not significant, suggesting additive negative effects of alcohol amount and frequent binging on the brain. The association between alcohol intake and brain health was not significantly different when the weekly units were consumed as wine, beer, or spirits (see overlapping 95% CI in supplementary Figure 16). Hence, we found no evidence that risk of alcohol-related brain harm differs according to alcoholic beverage type.

**Figure 5:**
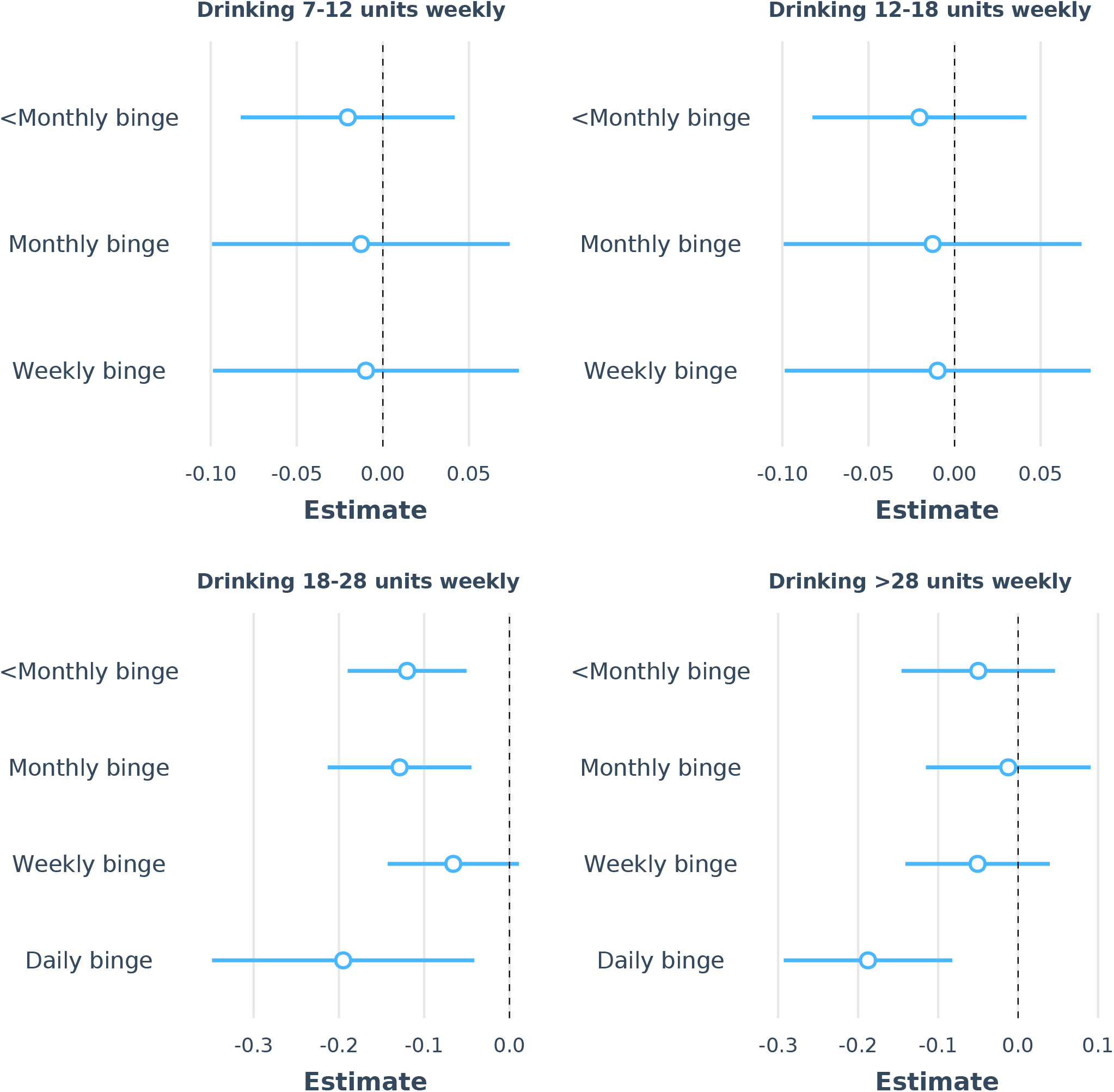
**Relation between binging frequency (>6 units/48g alcohol in one episode) and grey matter volume (normalized to head size), independent of alcohol consumption in units. N=12,812. Points show standardized regression coefficients (estimates and their 95% confidence intervals) for binging frequency category compared to the reference category (never binging) generated from regression models with grey matter volume as the dependent variable. Results are shown separately according to subjects’ weekly alcohol intake (divided into quantiles): 1) 6.8-11.6 units (54.1-92.8g), 2) 11.6-17.8 units (92.8-142.4g), 3) 17.8-28.4 units (142.4-227.2g), 4) 28.4-163 units (227.2-1304g). N=14,685. Regression models were adjusted for: alcohol consumption in weekly units, age, age^2^, age^3,^ sex, age x sex, age^2^ x sex, Diabetes Mellitus, SBP, DBP, BMI, non-HDL cholesterol, smoking status, imaging site, exercise minutes weekly, TDI, head size.**

### Alcohol and functional connectivity

Alcohol consumption was significantly associated with resting state functional connectivity of seven ‘nodes’ (supplementary Figure 18) thought to represent *within* network connectivity. Six of the seven nodes, all within the default mode network, demonstrated increased connectivity with higher alcohol intake. In turn, increased connectivity in two of these nodes predicted higher cognitive performance on tests of executive function and working memory (supplementary Figure 19). Alcohol intake was additionally associated with multiple ‘edges’ thought to represent functional connectivity *between* resting state networks (supplementary Figure 20).

## Discussion

### Statement of principal findings

Alcohol consumption was linearly and negatively associated with indices of brain health across most of the brain. There was a multiplicative interaction between alcohol and blood pressure and BMI in predicting grey matter volume. Additive harmful effects of alcohol volume and frequent binging were observed. In contrast there was no evidence that effects can be differentiated according to alcohol beverage type. Brain functional connectivity, related to cognitive function, was also associated with alcohol intake.

### Strengths and weaknesses of this study

Our study represents one of the largest imaging investigations into the impact of alcohol consumption on brain health to date. The very large sample size provided great statistical power to detect associations across almost the whole cortex, subcortical structures and cerebellum that have previously been uncharacterized, as well as extensively explore interactions with clinical and drinking behaviours. It used state-of-the-art neuroimaging and results were stringently controlled for more potential confounders than ever before, as well as multiple testing which increases confidence in the findings. Furthermore, sensitivity analyses included exploration of the robustness of the findings against unobserved confounders. Results were replicated using a voxel-wise whole brain approach as well as image-derived phenotypes. Medical diagnoses reflected real-world diagnoses made by clinicians. This also represents the first study to examine the relation of moderate drinking with functional connectivity in the brain.

It is important to acknowledge some limitations. UKB was selective with invitations generating only a 5% response rate. The sample is healthier, better educated, less deprived with less ethnic diversity than the general population [43]. This is likely to be an issue with any study requiring the intense participation necessary to investigate our research question (brain imaging, blood samples, multitude of questions) but raises the possibility that collider bias could cause spurious associations between alcohol and MRI [44]. However, we have examined the distribution of alcohol intake from UKB and Health Survey England and found no statically significant differences in proportions of daily and never drinkers, lessening the possibility of collider bias. Use of HES ICD diagnoses to define medical comorbidities means some cases would not have been captured. These would be likely to be of milder severity. The low numbers with serious liver disease and clinically significant depression in the sample limited our power to detect interactions with alcohol.

As with any observational study, we cannot infer causality from association. Traditional regression approaches assume no residual confounding (unverifiable for unmeasured variables) or reverse causality. Whilst we controlled analyses for all known confounders and were more thorough in this than any previously published study, we cannot exclude the possible of residual confounding. However, the sensitivity analyses estimated that unobserved confounding would need to be of a greater strength than any recognized observed confounder, including age and smoking, to obviate the association between alcohol and brain health, which seems implausible. The age when alcohol was self-reported limits the interpretation of the estimates to the impact of mid- to late-life consumption. Self-reported alcohol consumption may be liable to misclassification bias, but it remains the only realistic method of estimating intake on the large scale necessary for our research question. Blood measures are only sensitive to consistent heavy intake [45] and may reflect unrelated processes, and direct metabolites of alcohol such as phosphatidylethanol, ethyl glucuronide and ethyl sulphate are insensitive to moderate drinking and the cost would not justify use [46, 47]. Moreover, self-report is the method used in clinical practice, making findings from our study clinically translatable. Furthermore, random measurement error would bias associations towards the null outcome. Neuroimaging was cross-sectional and therefore we cannot examine the impact of alcohol on changes in brain measures over time. However, longitudinal imaging of sufficient duration to observe the impact of moderate drinking is likely to ‘suffer’ from MRI progress making scans incomparable. Reverse causation is unlikely because the earliest detectable brain changes occur in the late 40’s, by which time there have usually been decades of alcohol exposure. Alcohol is vasoactive [48], therefore it can disrupt coupling between neuronal activity and cerebral blood flow [49] used to make inferences in functional MRI. Resting state fMRI may be less sensitive to effects of vasodilation, but we cannot exclude such a phenomenon [50]. The proportion of participants who were drinking very heavily, or were alcohol dependent, was extremely low. Hence extrapolation of our results to this end of the intake spectrum is limited. Although there were substantial numbers binge-drinking to some extent, the middle- to older-aged UKB subjects may not be the ideal population in which to study binge-drinking, a more prevalent pattern of drinking in younger subjects. However, our primary focus was on more moderate alcohol intakes. When interpreting the results of our beverage type analysis, we are mindful that the group sizes were different, therefore giving greater power to detect associations amongst wine drinkers than amongst spirit drinkers. Additionally, whilst we controlled for multiple confounders, we cannot exclude the possibility that there were subtle differences between subjects drinking different alcoholic beverages that were not accounted for.

Whilst our outcomes of interest were quantitative and objective MRI measures rather than clinical phenotypes, they were selected because of their known dementia associations [28, 29, 51]. This gave us much greater power to detect non-linear relationships and investigate interactions than would have been possible with the low numbers in UKB with dementia diagnoses so far. UKB has a very limited cognitive battery of non-standard tests that would not necessarily capture deficits expected in relation to alcohol damage. There are also concerns about re-test reliability and significant floor effects in some tests [52].

### Strengths and weaknesses in relation to other studies

In this study, alcohol intake was associated with poorer brain integrity over almost the whole brain. Perhaps this is not a surprise, given the lipophilic characteristics of ethanol that allow passive diffusion across brain endothelial cell membranes and astrocyte wraps. We replicated the inverse associations between alcohol and hippocampal size we found in the Whitehall II study [10]. Our findings are consistent with results in the cingulate cortex and subcortical volumes reported by two other UKB analyses [11, 12]. However, our voxel-wise analyses enabled us to examine and spatially localise effects on grey and white matter across the whole brain. The persistence of associations with alcohol, despite adjustment for a greater number of potential confounders and the more punitive multiple testing correction necessitated by our voxel-wise approach, gives more confidence in the robustness of the findings. Formally testing more flexible statistical models enabled us to fully examine alcohol-brain health relationships and ascertain that the effects were linear rather than J-shaped. Adjustment for cardiovascular risk factors did not make a material difference to the strength of the associations, suggesting in this particular study they were neither confounders nor on the causal pathway. Associations in areas of the brain not previously implicated at lower levels of alcohol, including the basal ganglia and cerebellum, mirror findings in chronic heavy alcohol intake [53]. We suspect the discovery of these new linear associations result from our greater power to detect small effects. Similarly, the white matter associations we found replicated those we previously found in the corpus callosum [10], but extended to almost the whole white matter skeleton. In our case, analysis of different diffusion metrics revealed that a reduction in intracellular water and an increase in extracellular water were driving the fractional anisotropy results. This implies that a reduction in neurite density explains the white matter microstructural findings.

Higher blood pressure and BMI have been previously implicated in reducing grey matter volumes [25, 54]. To our knowledge their interaction with alcohol in predicting brain harm has not previously been explored. One mechanistic hypothesis for our findings is that hypertension may exacerbate alcohol-related brain harm by facilitating the diffusion of ethanol throughout brain tissue. Animal models have demonstrated increased cerebral blood flow (through arteriolar dilatation) and dysfunction of the blood brain barrier (BBB) in hypertension [55, 56]. Interestingly however, we found no significant interaction between alcohol and ApoE4 genotype, another factor which can break down the blood-brain barrier [57], although the relatively few E4 homozygotes may have limited our power to detect an effect. BMI may increase the adverse impact of alcohol via generation of toxic ceramides through a liver-brain axis [58]. Animal models have shown synergistic effects of obesity and alcohol on steatohepatitis [59].

Our findings that frequent binging has a negative impact on the brain additive to the effect of total alcohol consumed are in keeping with other health outcomes, including mortality [60], breast cancer [61], and cardiovascular disease [62]. We have two hypotheses to explain the findings. First, the peak ethanol (and its toxic breakdown product acetelaldehyde) level in the brain is likely to be higher during a binge. Since we found dose-dependent alcohol associations this would lead to more harm. Second, binging is often followed by a period of abstinence precipitating withdrawal. Alcohol withdrawal increases glutamate release, as well as activation of microglial cells and release of inflammatory cytokines. Both result in neuronal death. Repeated cycles of binging and withdrawal could heighten effects, as is postulated in the kindling theory [63]. Whilst most of our subjects were not alcohol-dependent, many did report binge drinking which could result in subclinical withdrawal with consequent neurochemical changes.

Studies of a range of health outcomes, including mortality, have postulated that the protective effects of moderate drinking are strongest in wine compared to beer or spirits [64], perhaps due to higher levels of polyphenols, in particular resveratrol in grape skins [17]. We found no evidence to suggest alcoholic beverage type confers differences in risks to the brain. This supports the hypothesis that it is ethanol itself, rather than other compounds in the beverage, that is on the biological pathway to damage. The associations of wine-drinking with higher educational level and socioeconomic status may explain the apparent health benefits [65].

This is the first study, to our knowledge, to find a relationship between alcohol consumption at the time of scanning and resting state functional connectivity in moderate drinkers. Resting state connectivity reflects current brain activity, which may be associated with contemporary alcohol consumption in a direct way, for instance by rebound from previous day, rather than chronic use. Participants were unlikely to have been under the influence of alcohol during the resting scan, although alcohol levels were not measured. Higher weekly consumption while sober during the scan could theoretically be associated with intermittent mild or subclinical withdrawal symptoms or some more persistent “silent” change in brain metabolism that is reflected in changed activity patterns. Interpretation of the finding of increased DMN connectivity with alcohol consumption is difficult, particularly as it is in the opposite direction to that reported in aging and Alzheimer’s disease [66]. The positive association of DMN connectivity with performance on cognitive testing at the time of scanning raises the possibility of a compensatory effect. However, given the limited number of cognitive tests associated, we caution not to over-interpret these findings. Of the few studies in alcohol dependent individuals, several have reported reduced functional connectivity in a range of networks, including visual, executive [67], salience [68, 69] and default mode networks [50] [70].

### Meaning of study for clinicians/policymakers

Our findings suggest that there is no safe level of alcohol consumption for brain health. Current low risk guidelines do not take account of the brain impact of drinking but should now be reviewed. Those with higher blood pressure and BMI, as well as those binge drinking, may be at increased risk of alcohol-related brain damage. Focusing interventions on these subgroups could represent a high yield strategy for harm reduction.

### Unanswered questions and future research

What remains unclear is the duration of drinking needed to cause an effect on the brain. There may be particular life periods, such as adolescence and older age, where dynamic brain changes occur that may lead to heightened vulnerability [71]. Studies in alcohol-dependent drinkers suggest at least some damage is reversible upon abstinence [72]. We do not know whether the same follows for moderate intakes.

## Supporting information

ICD codes

STROBE checklist

## Data Availability

Data is available to researchers affiliated with academic centres upon successful application to the UK access committee.

## Acknowledgements

We would like to thank the UKB participants for their participation in the study. We also give thanks to Professor David Leon who provided input to the study conceptualization and planning, as well as very helpful comments on the first draft of the manuscript.

## Contributions

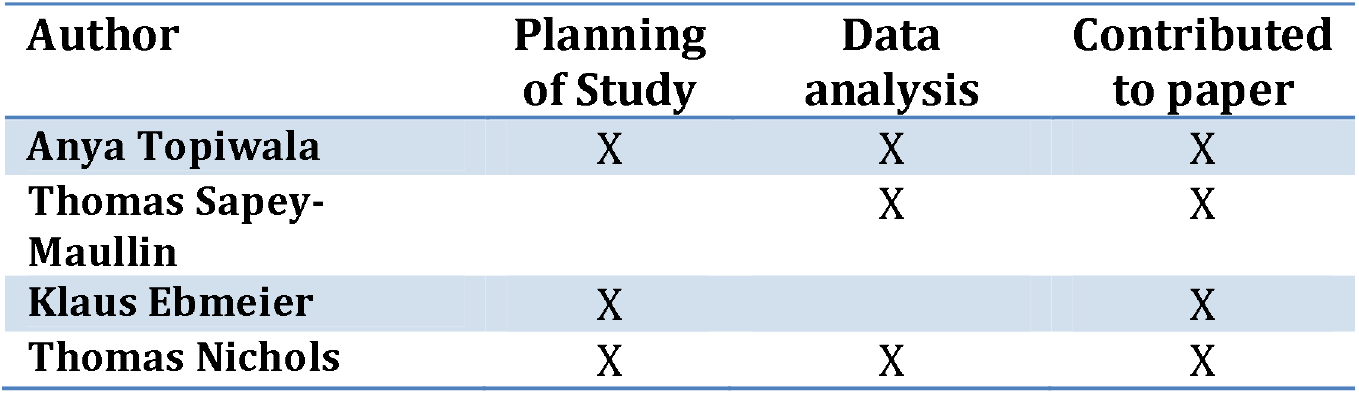

## Ethical approval

UKB approval was obtained from the North West Multi-Centre Research Ethics Committee and the Patient Information Advisory Group. The UKB Board and Access Sub-Committee and the Biobank Ethics and Governance Council reviewed our data access application.

## Transparency

Dr Anya Topiwala (manuscript’s guarantor) affirms that the manuscript is an honest, accurate and transparent account of the study being reported; that no important aspects of the study have been omitted; and that any discrepancies from the study as planned have been explained.

## Data availability

A CC BY licence is required.

## References

1. Statistics N. Statistics on Alcohol, England 2020. 2020.

2. Glynn RW, Shelley E, Lawlor BA. Public knowledge and understanding of dementia—evidence from a national survey in Ireland. Age and ageing. 2017;46(5):865–9.

3. Mukamal KJ, Phillips RS, Mittleman MA. Beliefs, motivations, and opinions about moderate drinking: a cross-sectional survey. FAMILY MEDICINE-KANSAS CITY-. 2008;40(3):188.

4. Wang Y, Pan Y, Li H. What is brain health and why is it important? bmj. 2020;371.

5. Paper UGG. Advancing Our Health Prevention in the 2020s. 2019.

6. Harper C, Kril J, Holloway R. Brain shrinkage in chronic alcoholics: a pathological study. Br Med J (Clin Res Ed). 1985;290(6467):501–4.

7. Schwarzinger M, Pollock BG, Hasan OS, Dufouil C, Rehm J, Group QS. Contribution of alcohol use disorders to the burden of dementia in France 2008–13: a nationwide retrospective cohort study. The Lancet Public Health. 2018.

8. Anstey KJ, Mack HA, Cherbuin N. Alcohol consumption as a risk factor for dementia and cognitive decline: meta-analysis of prospective studies. The American journal of Geriatric psychiatry. 2009;17(7):542–55.

9. Sabia S, Fayosse A, Dumurgier J, Dugravot A, Akbaraly T, Britton A, et al. Alcohol consumption and risk of dementia: 23 year follow-up of Whitehall II cohort study. Bmj. 2018;362.

10. Topiwala A, Allan CL, Valkanova V, Zsoldos E, Filippini N, Sexton C, et al. Moderate alcohol consumption as risk factor for adverse brain outcomes and cognitive decline: longitudinal cohort study. bmj. 2017;357:j2353.

11. Daviet R, Aydogan G, Jagannathan K, Spilka N, Koellinger P, Kranzler HR, et al. Multimodal brain imaging study of 19,825 participants reveals adverse effects of moderate drinking. BioRxiv. 2020.

12. Evangelou E, Suzuki H, Bai W, Pazoki R, Gao H, Matthews PM, et al. Alcohol consumption in the general population is associated with structural changes in multiple organ systems: A population-based study in UK Biobank. medRxiv. 2021.

13. Roerecke M, Tobe SW, Kaczorowski J, Bacon SL, Vafaei A, Hasan OS, et al. Sex-specific associations between alcohol consumption and incidence of hypertension: a systematic review and meta-analysis of cohort studies. Journal of the American Heart Association. 2018;7(13):e008202.

14. Qizilbash N, Gregson J, Johnson ME, Pearce N, Douglas I, Wing K, et al. BMI and risk of dementia in two million people over two decades: a retrospective cohort study. The Lancet Diabetes & Endocrinology. 2015;3(6):431–6.

15. Den Heijer T, Skoog I, Oudkerk M, De Leeuw F-E, De Groot JC, Hofman A, et al. Association between blood pressure levels over time and brain atrophy in the elderly. Neurobiology of aging. 2003;24(2):307–13.

16. Yamazaki Y, Zhao N, Caulfield TR, Liu C-C, Bu G. Apolipoprotein E and Alzheimer disease: pathobiology and targeting strategies. Nature Reviews Neurology. 2019;15(9):501–18.

17. Pignatelli P, Ghiselli A, Buchetti B, Carnevale R, Natella F, Germano G, et al. Polyphenols synergistically inhibit oxidative stress in subjects given red and white wine. Atherosclerosis. 2006;188(1):77–83.

18. Hunt WA. Are binge drinkers more at risk of developing brain damage? Alcohol. 1993;10(6):559–61.

19. Funpack. Funpack.

20. Clarke T-K, Adams MJ, Davies G, Howard DM, Hall LS, Padmanabhan S, et al. Genome-wide association study of alcohol consumption and genetic overlap with other health-related traits in UK Biobank (N= 112 117). Molecular psychiatry. 2017;22(10):1376.

21. Genin E, Hannequin D, Wallon D, Sleegers K, Hiltunen M, Combarros O, et al. APOE and Alzheimer disease: a major gene with semi-dominant inheritance. Molecular psychiatry. 2011;16(9):903–7.

22. Gianaros PJ, Greer PJ, Ryan CM, Jennings JR. Higher blood pressure predicts lower regional grey matter volume: Consequences on short-term information processing. Neuroimage. 2006;31(2):754–65.

23. Maillard P, Seshadri S, Beiser A, Himali JJ, Au R, Fletcher E, et al. Effects of systolic blood pressure on white-matter integrity in young adults in the Framingham Heart Study: a cross-sectional study. The Lancet Neurology. 2012;11(12):1039–47.

24. Whalley L, Staff R, Murray A, Duthie S, Collins A, Lemmon H, et al. Plasma vitamin C, cholesterol and homocysteine are associated with grey matter volume determined by MRI in non-demented old people. Neuroscience letters. 2003;341(3):173–6.

25. Herrmann MJ, Tesar AK, Beier J, Berg M, Warrings B. Grey matter alterations in obesity: A meta-analysis of whole-brain studies. Obesity reviews. 2019;20(3):464–71.

26. Alfaro-Almagro F, Jenkinson M, Bangerter NK, Andersson JL, Griffanti L, Douaud G, et al. Image processing and Quality Control for the first 10,000 brain imaging datasets from UK Biobank. Neuroimage. 2018;166:400–24.

27. Patenaude B, Smith SM, Kennedy DN, Jenkinson M. A Bayesian model of shape and appearance for subcortical brain segmentation. Neuroimage. 2011;56(3):907–22.

28. Mckhann GM, Knopman DS, Chertkow H, Hyman BT, Jack CR, Kawas CH, et al. The diagnosis of dementia due to Alzheimer’s disease: Recommendations from the National Institute on Aging-Alzheimer’s Association workgroups on diagnostic guidelines for Alzheimer’s disease. Alzheimer’s & dementia. 2011;7(3):263–9.

29. Black S, Gao F, Bilbao J. Understanding white matter disease: imaging-pathological correlations in vascular cognitive impairment. Stroke. 2009;40(3 suppl 1):S48–S52.

30. Greicius MD, Srivastava G, Reiss AL, Menon V. Default-mode network activity distinguishes Alzheimer’s disease from healthy aging: evidence from functional MRI. Proceedings of the National Academy of Sciences of the United States of America. 2004;101(13):4637–42.

31. Zhang Y, Brady M, Smith S. Segmentation of brain MR images through a hidden Markov random field model and the expectation-maximization algorithm. IEEE transactions on medical imaging. 2001;20(1):45–57.

32. Zhang H, Schneider T, Wheeler-Kingshott CA, Alexander DC. NODDI: practical in vivo neurite orientation dispersion and density imaging of the human brain. Neuroimage. 2012;61(4):1000–16.

33. Douaud G, Smith S, Jenkinson M, Behrens T, Johansen-Berg H, Vickers J, et al. Anatomically related grey and white matter abnormalities in adolescent-onset schizophrenia. Brain. 2007;130(9):2375–86.

34. Good CD, Johnsrude I, Ashburner J, Henson RN, Friston KJ, Frackowiak RS. Cerebral asymmetry and the effects of sex and handedness on brain structure: a voxel-based morphometric analysis of 465 normal adult human brains. Neuroimage. 2001;14(3):685–700.

35. Smith SM, Jenkinson M, Woolrich MW, Beckmann CF, Behrens TE, Johansen-Berg H, et al. Advances in functional and structural MR image analysis and implementation as FSL. Neuroimage. 2004;23:S208–S19.

36. Andersson JL, Jenkinson M, Smith S. Non-linear registration, aka Spatial normalisation FMRIB technical report TR07JA2. FMRIB Analysis Group of the University of Oxford. 2007.

37. Smith SM, Jenkinson M, Johansen-Berg H, Rueckert D, Nichols TE, Mackay CE, et al. Tract-based spatial statistics: voxelwise analysis of multi-subject diffusion data. Neuroimage. 2006;31(4):1487–505.

38. Smith SM, Jenkinson M, Johansen-Berg H, Rueckert D, Nichols TE, Mackay CE, et al. Tract-based spatial statistics: voxelwise analysis of multi-subject diffusion data. Neuroimage. 2006;31(4):1487–505.

39. Maullin-Sapey T NT. BLM: Parallelised Computing for Big Linear Models.. 2019.

40. Stone CJ. [Generalized additive models]: comment. Statistical Science. 1986;1(3):312–4.

41. Alfaro-Almagro F, Mccarthy P, Afyouni S, Andersson JL, Bastiani M, Miller KL, et al. Confound modelling in UK Biobank brain imaging. NeuroImage. 2021;224:117002.

42. Cinelli C, Hazlett C. Making sense of sensitivity: Extending omitted variable bias. Journal of the Royal Statistical Society: Series B (Statistical Methodology). 2020;82(1):39–67.

43. Fry A, Littlejohns T, Sudlow C, Doherty N, Allen N. OP41 The representativeness of the UK Biobank cohort on a range of sociodemographic, physical, lifestyle and health-related characteristics. BMJ Publishing Group Ltd; 2016.

44. Munafò MR, Tilling K, Taylor AE, Evans DM, Davey Smith G. Collider scope: when selection bias can substantially influence observed associations. International journal of epidemiology. 2018;47(1):226–35.

45. Conigrave KM, Davies P, Haber P, Whitfield JB. Traditional markers of excessive alcohol use. Addiction. 2003;98(2):31–43.

46. Schröck A, Wurst FM, Thon N, Weinmann W. Assessing phosphatidylethanol (PEth) levels reflecting different drinking habits in comparison to the alcohol use disorders identification test-C (AUDIT-C). Drug and Alcohol Dependence. 2017.

47. Paull P, Haber PS, Chitty K, Seth D. Evaluation of a novel method for the analysis of alcohol biomarkers: Ethyl glucuronide, ethyl sulfate and phosphatidylethanol. Alcohol. 2018;67:7–13.

48. Kawano Y. Physio-pathological effects of alcohol on the cardiovascular system: its role in hypertension and cardiovascular disease. Hypertension Research. 2010;33(3):181–91.

49. Luchtmann M, Jachau K, Tempelmann C, Bernarding J. Alcohol induced region-dependent alterations of hemodynamic response: implications for the statistical interpretation of pharmacological fMRI studies. Experimental brain research. 2010;204(1):1–10.

50. Du C, Volkow ND, Koretsky AP, Pan Y. Low-frequency calcium oscillations accompany deoxyhemoglobin oscillations in rat somatosensory cortex. Proceedings of the National Academy of Sciences. 2014;111(43):E4677–E86.

51. Greicius MD, Krasnow B, Reiss AL, Menon V. Functional connectivity in the resting brain: a network analysis of the default mode hypothesis. Proceedings of the National Academy of Sciences. 2003;100(1):253–8.

52. Fawns-Ritchie C, Deary IJ. Reliability and validity of the UK Biobank cognitive tests. PloS one. 2020;15(4):e0231627.

53. Zahr NM, Kaufman KL, Harper CG. Clinical and pathological features of alcohol-related brain damage. Nature Reviews Neurology. 2011;7(5):284–94.

54. Beauchet O, Celle S, Roche F, Bartha R, Montero-Odasso M, Allali G, et al. Blood pressure levels and brain volume reduction: a systematic review and meta-analysis. Journal of hypertension. 2013;31(8):1502–16.

55. Mackenzie ET, Strandgaard S, Graham DI, Jones JV, Harper AM, Farrar JK. Effects of acutely induced hypertension in cats on pial arteriolar caliber, local cerebral blood flow, and the blood-brain barrier. Circulation research. 1976;39(1):33–41.

56. Biancardi VC, Son SJ, Ahmadi S, Filosa JA, Stern JE. Circulating angiotensin II gains access to the hypothalamus and brain stem during hypertension via breakdown of the blood–brain barrier. Hypertension. 2014;63(3):572–9.

57. Montagne A, Nation DA, Sagare AP, Barisano G, Sweeney MD, Chakhoyan A, et al. APOE4 leads to blood–brain barrier dysfunction predicting cognitive decline. Nature. 2020;581(7806):71–6.

58. De La Monte SM, Longato L, Tong M, Denucci S, Wands JR. The liver-brain axis of alcohol-mediated neurodegeneration: role of toxic lipids. International journal of environmental research and public health. 2009;6(7):2055–75.

59. Xu J, Lai KK, Verlinsky A, Lugea A, French SW, Cooper MP, et al. Synergistic steatohepatitis by moderate obesity and alcohol in mice despite increased adiponectin and p-AMPK. Journal of hepatology. 2011;55(3):673–82.

60. Kauhanen J, Kaplan GA, Goldberg DE, Salonen JT. Beer binging and mortality: results from the Kuopio ischaemic heart disease risk factor study, a prospective population based study. Bmj. 1997;315(7112):846–51.

61. White AJ, Deroo LA, Weinberg CR, Sandler DP. Lifetime alcohol intake, binge drinking behaviors, and breast cancer risk. American journal of epidemiology. 2017;186(5):541–9.

62. Mckee M, Britton A. The positive relationship between alcohol and heart disease in eastern Europe: potential physiological mechanisms. Journal of the Royal society of medicine. 1998;91(8):402–7.

63. Brown ME, Anton RF, Malcolm R, Ballenger JC. Alcohol detoxification and withdrawal seizures: clinical support for a kindling hypothesis. Biological psychiatry. 1988;23(5):507–14.

64. Gronbaek M, Deis A, Sorensen TI, Becker U, Schnohr P, Jensen G. Mortality associated with moderate intakes of wine, beer, or spirits. Bmj. 1995;310(6988):1165–9.

65. Mortensen EL, Jensen HH, Sanders SA, Reinisch JM. Better psychological functioning and higher social status may largely explain the apparent health benefits of wine: a study of wine and beer drinking in young Danish adults. Archives of internal medicine. 2001;161(15):1844–8.

66. Tomasi D, Volkow ND. Aging and functional brain networks. Molecular psychiatry. 2012;17(5):549–58.

67. Camchong J, Stenger A, Fein G. Resting-state synchrony during early alcohol abstinence can predict subsequent relapse. Cerebral Cortex. 2012:bhs190.

68. Sullivan EV, Müller-Oehring E, Pitel A-L, Chanraud S, Shankaranarayanan A, Alsop DC, et al. A selective insular perfusion deficit contributes to compromised salience network connectivity in recovering alcoholic men. Biological psychiatry. 2013;74(7):547–55.

69. Müller-Oehring EM, Jung Y-C, Pfefferbaum A, Sullivan EV, Schulte T. The resting brain of alcoholics. Cerebral cortex. 2014:bhu134.

70. Chanraud S, Pitel A-L, Pfefferbaum A, Sullivan EV. Disruption of functional connectivity of the default-mode network in alcoholism. Cerebral Cortex. 2011;21(10):2272–81.

71. Mewton L, Lees B, Rao RT. Lifetime perspective on alcohol and brain health. British Medical Journal Publishing Group; 2020.

72. Schroth G, Naegele T, Klose U, Mann K, Petersen D. Reversible brain shrinkage in abstinent alcoholics, measured by MRI. Neuroradiology. 1988;30(5):385–9.

