## Supplementary methods and tables for "No safe level of alcohol consumption for brain health: observational cohort study of 25,378 UK Biobank participants"

***MRI acquisition***

As part of the protocol [www.fmrib.ox.ac.uk/ukbiobank/protocol/V3_23092014.pdf] T1-weighted images were obtained using an MPRAGE sequence: TR=2000ms, TE=2.0ms, 208 sagittal slices, flip angle=8°, FOV=256mm, matrix=256x256, slice thickness=1.0mm (voxel size 1x1x1mm). Diffusion images were obtained using a spin-echo echo-planar sequence with 10 T2-weighted baseline volumes, 50b = 1000 s mm^-2^ and 50 b=2000 s mm^-2^ diffusion weighted volumes, with 100 diffusion-encoding directions and 2mm isotropic voxels. Resting state functional MRI was acquired with the following parameters: TE=39ms, TR=735ms, MB=8, R=1, flip angle=52°, 490 time points, 2.4x2.4x2.4mm voxel size.

***MRI pre-processing***

T1 structural images were gradient distortion corrected and registered linearly and non-linearly (using FMRIB’s Linear Registration Tool, FLIRT [21] and FMRIB’s Nonlinear Image Registration Tool, FNIRT [22]) to standard space. Brain extraction (using Brain Extraction Tool, BET [23], defacing and segmentation into tissue types (using FMRIB’s Automated Segmentation Tool, FAST [24]) were then performed. Volumes for subcortical structures were generated by modeling using FMRIB’s Integrated Registration and Segmentation Tool (FIRST [25]).

Diffusion images were corrected for eddy currents, head motion and gradient distortion. Using the tool DTIFIT a diffusion tensor was fit generating fractional anisotropy (FA), tensor mode, radial and mean diffusivities. FA images were fed into Tract-Based Spatial Statistics (TBSS [26]). This aligns the image onto a standard-space white matter skeleton. Additionally dMRI was fed into NODDI (Neurite Orientation Dispersion and Density Imaging [27]) to generate white matter microstructural parameters including intra-cellular volume fraction (icvf), isotropic water volume fraction (isovf) and orientation dispersion index (odi). Skeletonised images were averaged within a set of standard-space tract masks to generate mean values [28].

The pipeline for rsfMRI images used MELODIC [29] which performs EPI unwarping, gradient correction unwarping, motion correction, intensity normalization and high pass temporal filtering. Artefacts were removed using independent component analysis and FMRIB’s ICA-based X-noiseifer (FIX [30]). Group-averaged principal components analysis of resting state networks was carried out using a subset of subjects using MELODIC. The ICA spatial maps were mapped onto each subject’s rfMRI timeseries data to derive a timeseries for each subject for each network (‘node’). These timeseries were also used to estimate subject-specific network matrices using FSLNets [31]. Partial temporal correlations (aiming to estimate direct connection strengths better) between nodes’ timeseries were denoted as ‘edges’.

**Index of supplementary figures and tables**

**Figure 1:**	Flow chart of included participants

**Figure 2:**	Cumulative distribution alcohol consumption, whole sample

**Table 1:**	Predictors of binging frequency

**Figure 3:**	Cumulative distribution of alcohol intake, wine, beer and spirits

**Table 2:**	Baseline characteristics according to alcoholic beverage consumed

**Figure 4:**	VBM with additional imaging-related confounder adjustment

**Figure 5:**	VBM results, current drinkers only

**Figure 6:**	Voxelwise variance of grey matter density explained by alcohol intake, all subjects

**Table 3:**	Variance of grey matter volume explained by modifiable risk factors

**Figure 7:**	Sensitivity contour plot for alcohol-grey matter estimate

**Table 4:**	Regression models for grey matter IDPs

**Table 5:**	Regression models for grey matter with and without cardiovascular risk factors

**Figure 8:**	Predicted grey matter IDP volumes according to alcohol intake, drinkers

**Figure 9:**	Predicted DTI indices according to alcohol intake, all subjects

**Table 6:**	Regression models for diffusion IDPs

**Figure 10:**	TBSS - positive correlation alcohol with mean diffusivity, all subjects

**Figure 11:**	TBSS - negative correlation alcohol with mode, all subjects

**Figure 12:**	TBSS - positive correlation alcohol with L2, all subjects

**Figure 13:**	TBSS - negative correlation alcohol with L3, all subjects

**Figure 14:**	TBSS - additional image-related confounder adjustment

**Figure 15:**	TBSS, current drinkers only

**Table 7:**	Regression models including significant interactions

**Table 8:**	Regression models including non-significant interactions

**Table 9:**	Regression models examining binge-drinking frequency

**Figure 16:**	Comparison of wine, beer and spirit drinking in predicting grey matter

**Figure 17:**	Group average spatial maps for the resting state fMRI nodes significantly associated	with alcohol

**Figure 18:** Predicted resting state node amplitude according to alcohol intake

**Figure 19:** Associations between rsfMRI nodes and cognitive test performance

**Table 10:** Associations between alcohol and resting fMRI edges

**Figure 20:** Associations between alcohol and resting fMRI edges (p values)

**Figure 21:** Associations between alcohol and resting fMRI edges (effect sizes)

Structural abnormality, missing alcohol or confounder data. N=23,261

UK Biobank participants with single MRI brain

N=43,572

Subjects in VBM analyses, with usable images, complete alcohol and confounder data

N=25,378

N=550

TBSS could not be run.

N=1348

Subjects in TBSS analyses

N=24,030

Missing FIRST volumes or diffusion parameters

N=1776 write.csv(idp2, "/well/nichols/users/wld330/imaging_48k/brain_idp/idp_quant.csv")

Subjects included in structural IDP analyses

N=22,254

Subjects included in functional IDP analyses

N=17,587

**Figure 1: Flow chart of participants included in analysis.**

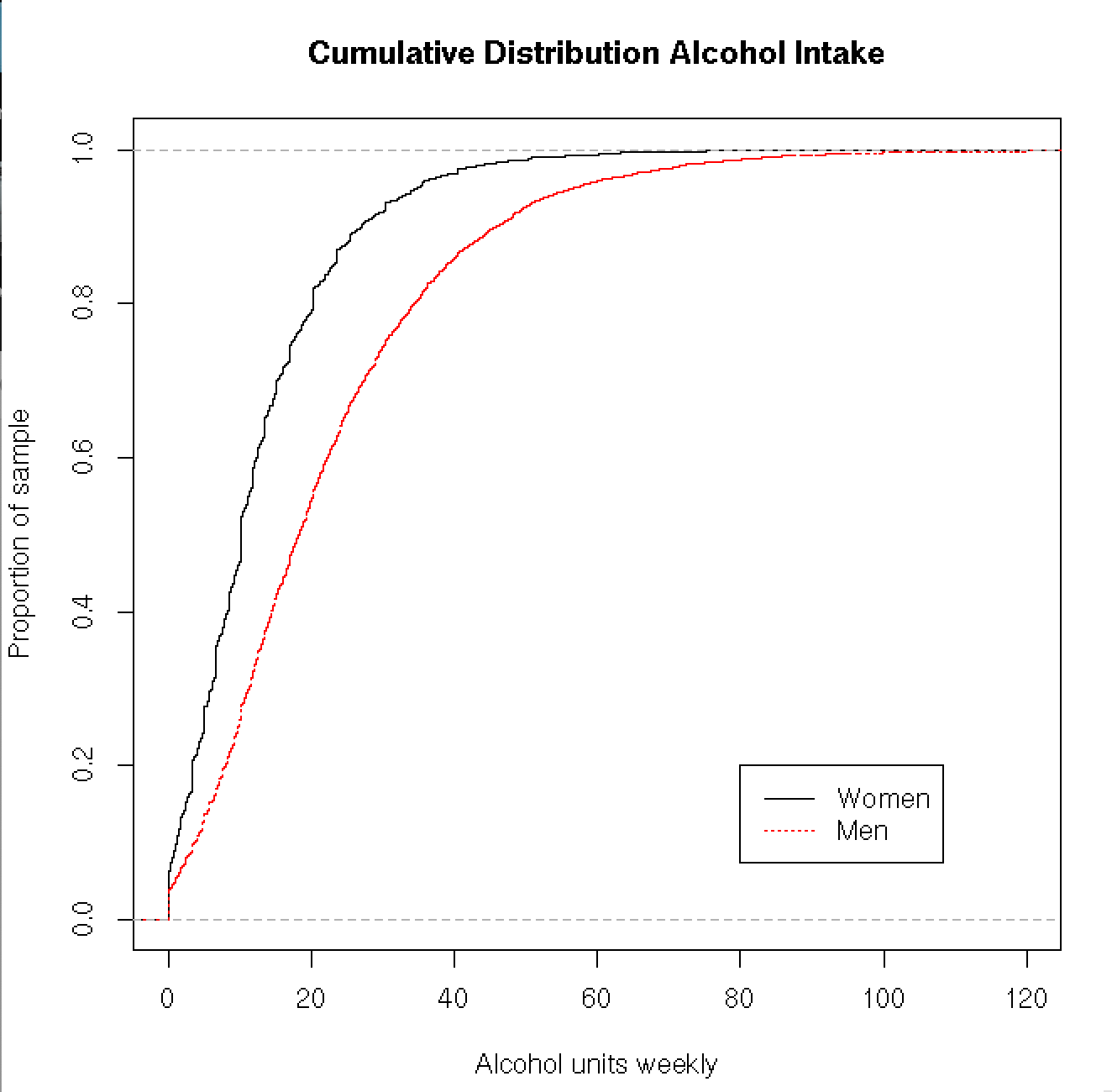

**Figure 2: Cumulative distribution of alcohol consumption separately by sex, weekly units. N=25,378.**

| Sociodemographic factor | Coefficient | 95% CI | P value* |
| --- | --- | --- | --- |
| Age | -0.07 | -0.07 to –0.06 | 4.40 x 10^-210^ |
| Sex | 0.21 | 0.14 to 0.27 | 4.60 x 10^-10^ |
| Current vs. never smoker | 0.52 | 0.39 to 0.65 | 3.30 x 10^-15^ |
| Degree vs. no qualifications | 0.51 | 0.35 to 0.67 | 1.00 x 10^-9^ |
| Townsend Deprivation Index | 0.02 | 0.01 to 0.03 | 3.58 x 10^-4^ |

*** Generated by comparing the t-value against the standard normal distribution**

**Table 1: Predictors of alcohol binging frequency. Binging defined as >6 units (48g) alcohol in one episode. Estimates generated using ordinal logistic regression, with binging frequency as the dependent variable, and each sociodemographic factor entered separately. Models were adjusted for weekly alcohol intake in units. N=14,685.**

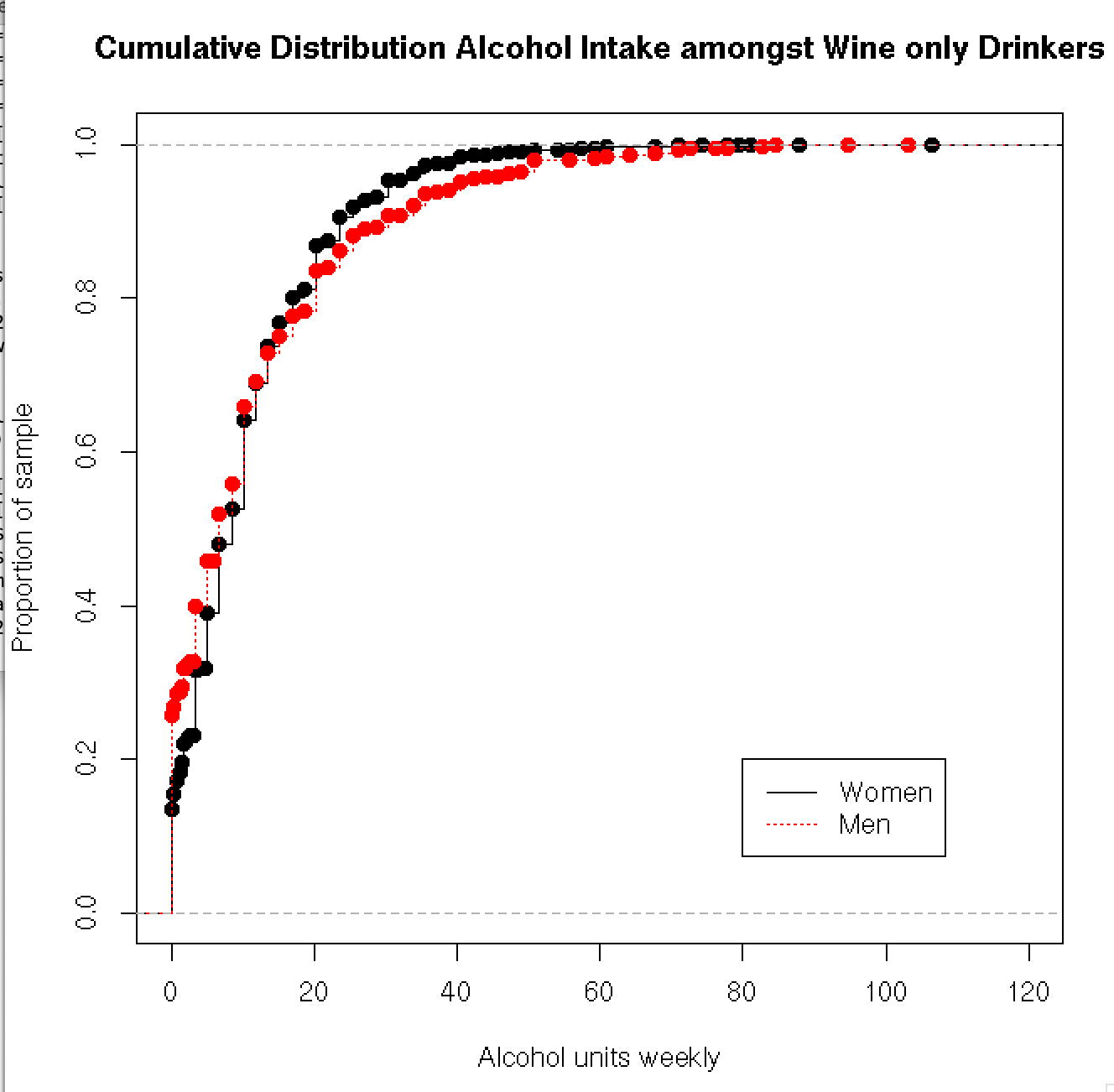

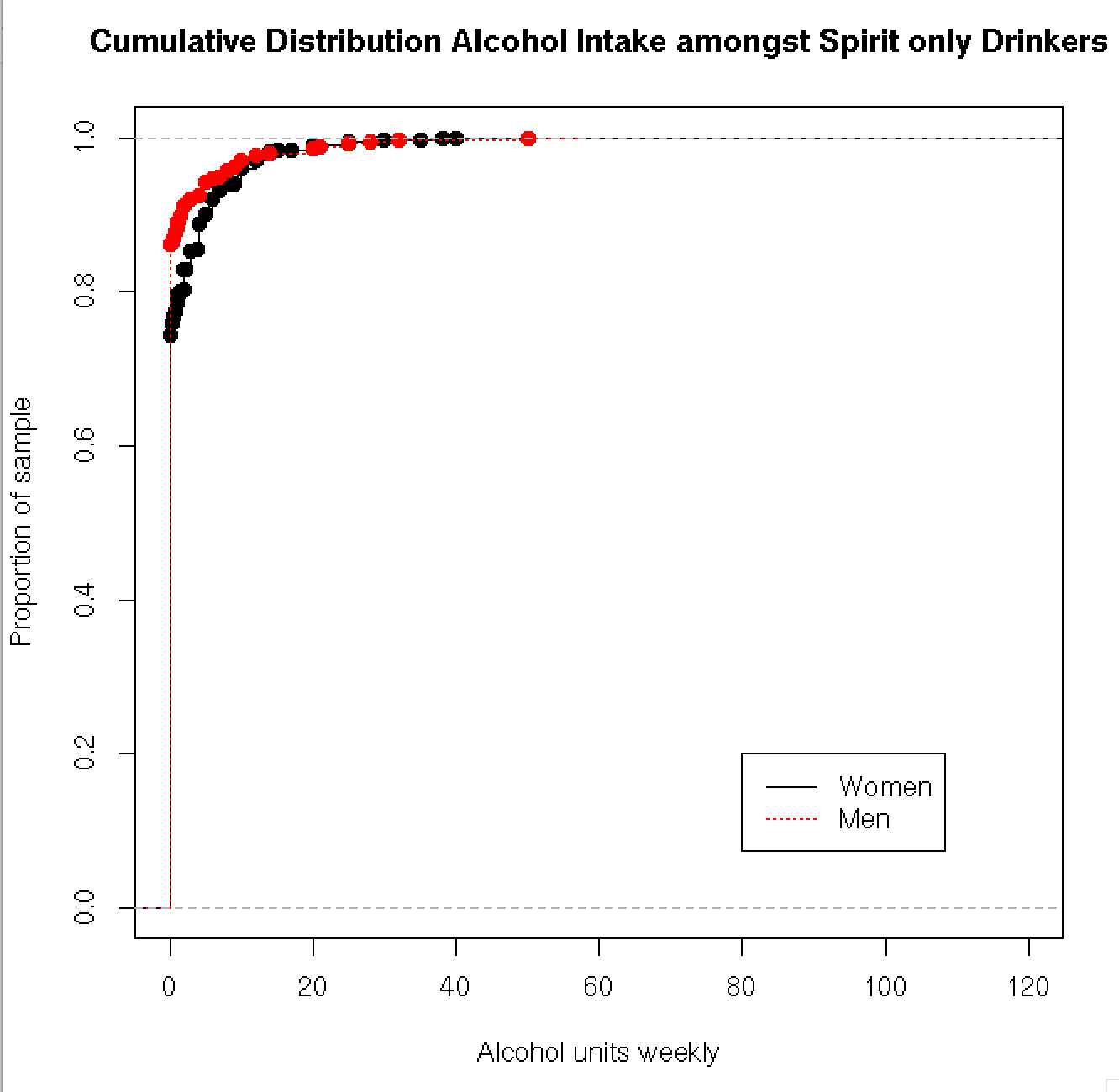

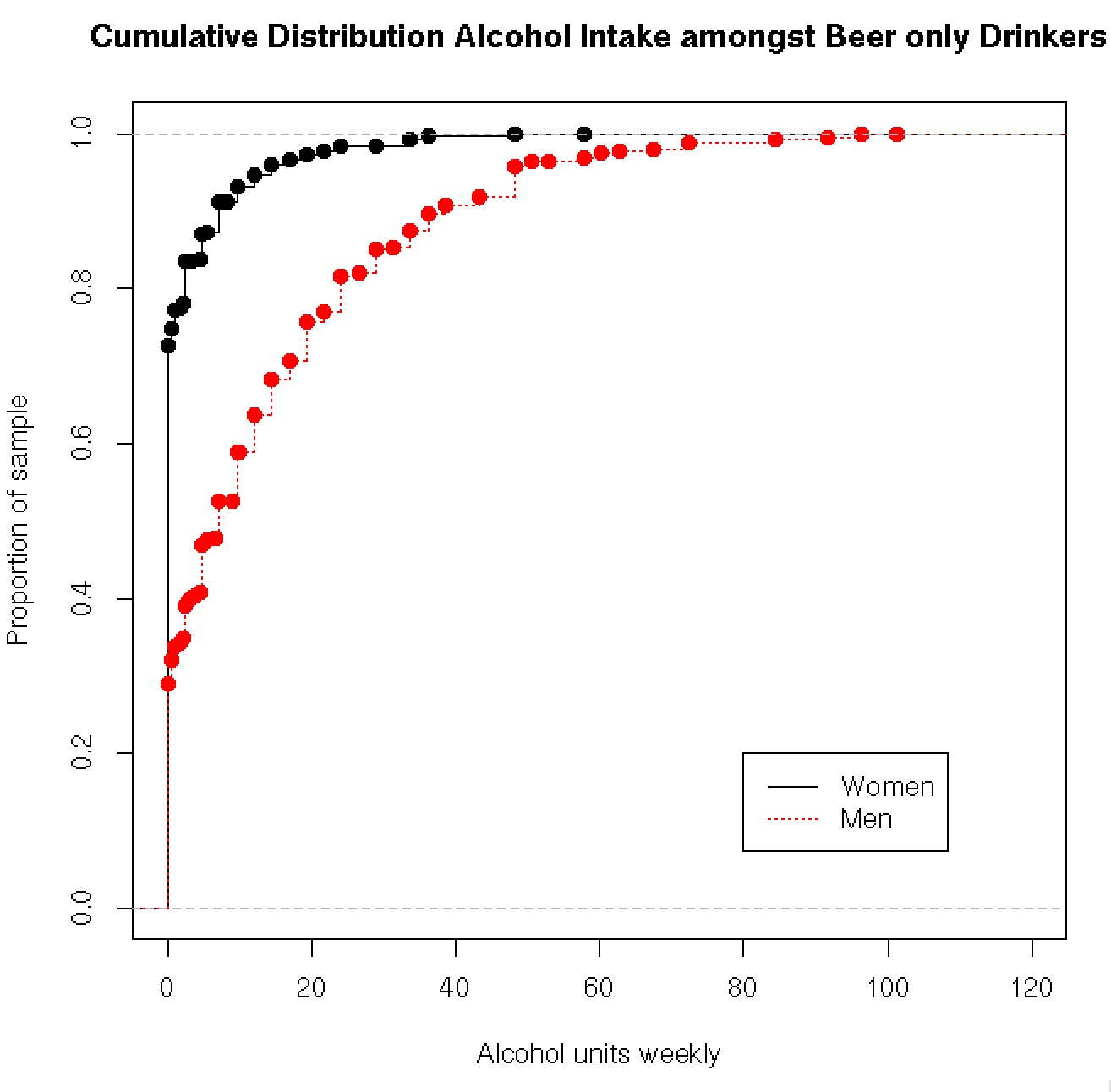

**Figure 3: Cumulative distribution of alcohol intake in weekly units for subjects drinking solely wine, beer and spirits, by sex.**

|  | Wine drinkers (N=5080) | Beer drinkers (N=1193) | Spirits drinkers (N=329) | Group differences^3^ | |
| --- | --- | --- | --- | --- | --- |
| Age^2^, years | 55.24 (7.29) | 53.39 (7.64) | 54.92 (8.00) | | F(1,6600)=28.28,p=1.09x10^-7^ |
| Sex, N(%) female | 3996 (78.66) | 280 (23.47) | 259 (78.72) | | X^2^=1384.6, df=2,p<2.2x10^-16^ |
| Qualifications^4^, *none* | 232 (4.57) | 122 (10.23) | 43 (12.07) | | X^2^=375.55, df=12, p<2.2x10^-16^ |
| *A levels* | 689 (13.56) | 150 (12.57) | 42 (12.77) | |  |
| *Degree* | 2262 (52.40) | 381 (31.94) | 103 (31.32) | |  |
| BMI^2^ | 25.69 (4.05) | 27.26 (4.27) | 26.69 (4.46) | | F(1,6600)=107.9,p<2x10^-16^ |
| SBP^2^ | 134.88 (19.01) | 138.35 (17.67) | 133.97 (18.37) | | F(1,6600)=8.62,p=0.003 |
| DBP^2^ | 80.11 (10.31) | 82.86 (10.49) | 79.78 (10.42) | | F(1,6600)=22.18, p=2.53x10^-6^ |
| Cholesterol^2^ | 5.79 (1.06) | 5.60 (1.08) | 5.80 (1.10) | | F(1,6600)=10.71,p=0.001 |
| Exercise^1^ | 117.40 (86.01) | 141.32 (112.52) | 128.77 (101.07) | | F(1,6600)=42.88, p=6.25x10^-11^ |
| Townsend Deprivation Index^2^ | -2.14 (2.54) | -1.44 (2.80) | -1.64 (2.77) | | F(1,6600)=57.29, p=4.28x10^-14^ |
| Smoking status *never* | 3257 (64.11) | 665 (55.74) | 202 (61.40) | | X^2^=83.62, df=4,p<2.2x10^-16^ |
| *previous* | 1608 (31.64) | 406 (34.03) | 97 (29.48) | |  |
| *current* | 215 (4.23) | 122 (10.23) | 30 (9.12) | |  |
| Diabetes Mellitus | 159 (3.13) | 85 (7.12) | 17 (5.17) | | X^2^=41.95, df=2,p<7.77x10^-10^ |

**^1^ MET minutes of moderate or vigorous activity.**

**^2^ Mean (standard deviation).**

**^3^** **One-way ANOVA for continuous variables, chi-square test for categorical variables.**

**^4^ Only selected qualification categories are presented for brevity.**

**Table 2: Baseline characteristics according to alcohol beverage consumed (those drink only one type).**

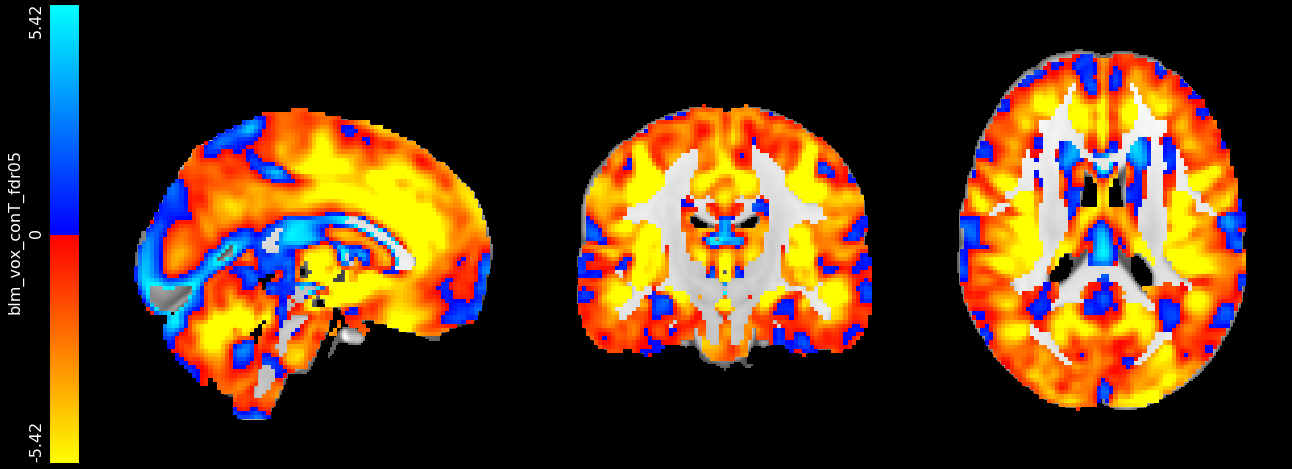

**Figure 4: VBM with additional imaging-related confounder adjustment (n=22,536). Associations between weekly alcohol intake and grey matter density generated using voxel-based morphometry. Adjusted for: age, sex, age^2^, age^3^, age x sex, age^2^ x sex, imaging site, SBP, DBP, cholesterol, HDL, Diabetes Mellitus, smoking, BMI, exercise, TDI, depression, qualifications, head size, head motion, table position, acquisition parameters. FDR-adjusted T statistics are shown according to the colour bar.**

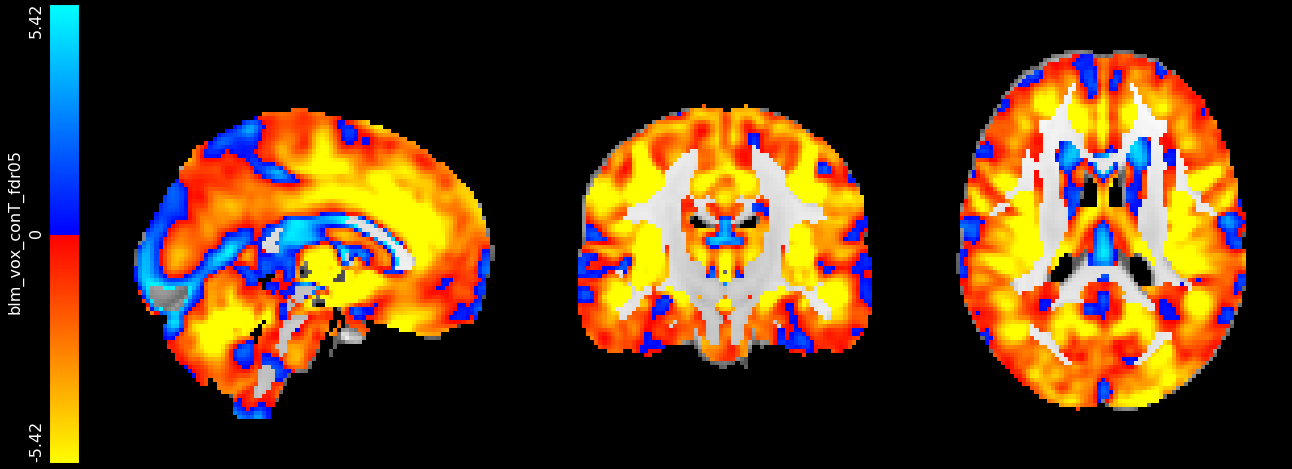

**Figure 5: VBM results with current drinkers only (n=24069). Associations between weekly alcohol intake and grey matter density generated using voxel-based morphometry. Adjusted for: age, sex, age^2^, age^3^, age x sex, age^2^ x sex, imaging site, SBP, DBP, cholesterol, HDL, Diabetes Mellitus, smoking, BMI, exercise, TDI, depression, qualifications, head size. FDR-adjusted T statistics are shown according to the colour bar.**

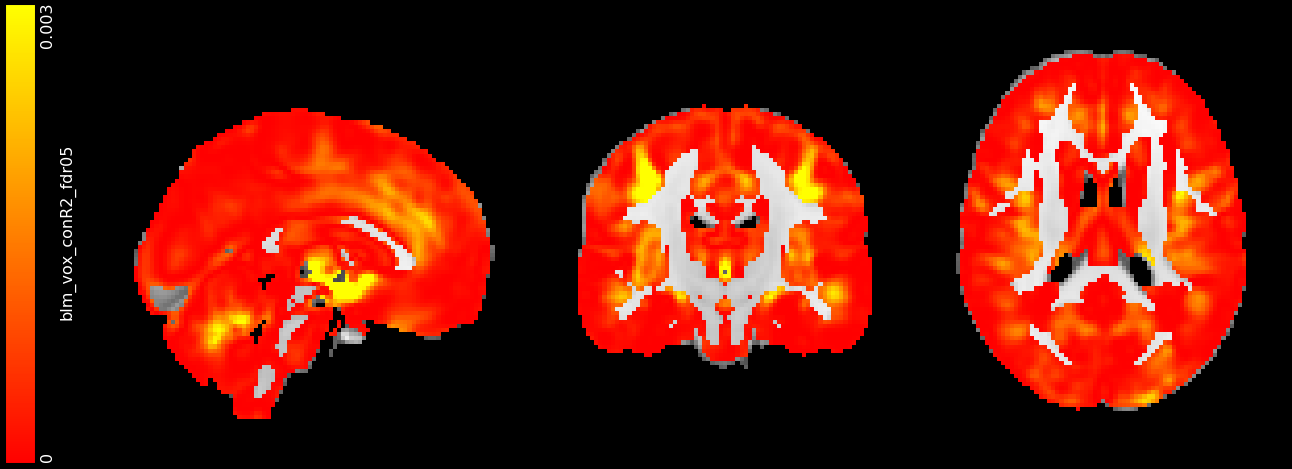

**Figure 6: Variance of grey matter density explained by weekly alcohol intake (R^2^ values) shown voxel-wise N=25,378 subjects. Adjusted for: age, sex, age^2^, age^3^, age x sex, age^2^ x sex, imaging site, SBP, DBP, cholesterol, HDL, Diabetes Mellitus, smoking, BMI, exercise, TDI, depression, qualifications, head size. FDR-adjusted T statistics are shown according to the colour bar.**

| Risk factor | Total R^2^ of model excluding risk factor | R^2^ explained by risk factor^*^ (%) |
| --- | --- | --- |
| Alcohol consumption | 0.5006 | 0.77 |
| Smoking status | 0.5066 | 0.17 |
| BMI | 0.5061 | 0.22 |
| Systolic blood pressure | 0.5082 | 0.0001 |
| Diastolic blood pressure | 0.5075 | 0.0008 |
| Non-HDL cholesterol | 0.5079 | 0.0004 |
| All factors in model | **0.5083** | **50.83** |

**^*^ Calculated by change in adjusted R^2^**

**Table 3: Variance of grey matter volume (normalized) explained by modifiable risk factors. N=22,254 subjects.**

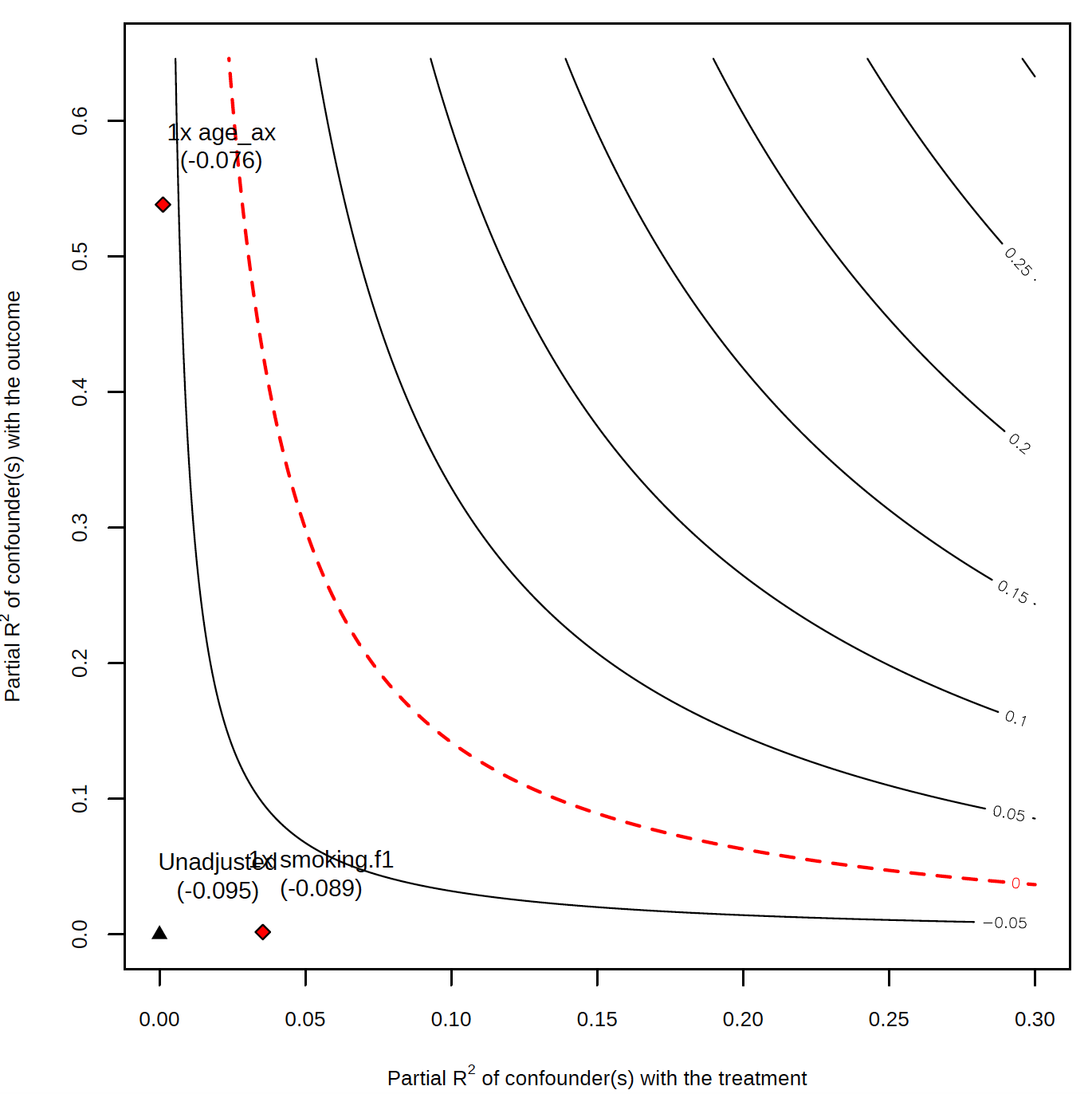

**Figure 7: Sensitivity contour plot of the point estimate for the association of alcohol intake and grey matter volume. The unadjusted estimate is shown by the black triangle. Contours indicate how the estimate would change based on the strength of an unobserved confounder with the treatment (alcohol intake – x axis) and outcome (grey matter volume – y axis). Red diamonds indicate the strength of the observed confounders age and smoking.**

|  | **Total grey matter** | **Right hippocampus** | **Left hippocampus** | **Right putamen** | **Left**  **putamen** | **Right thalamus** | **Left thalamus** |
| --- | --- | --- | --- | --- | --- | --- | --- |
| **Alcohol** | -0.10^***^ (-0.11, -0.09) | -0.04^***^ (-0.05, -0.03) | -0.04^***^ (-0.05, -0.03) | -0.03^***^ (-0.04, -0.02) | -0.03^***^ (-0.04, -0.02) | -0.06^***^ (-0.07, -0.05) | -0.05^***^ (-0.06, -0.04) |
| **Age** | 1.07 (-0.33, 2.47) | 1.16 (-0.61, 2.93) | 0.38 (-1.42, 2.17) | 0.78 (-0.78, 2.33) | 0.95 (-0.63, 2.52) | 1.85^***^ (0.47, 3.23) | 1.32^*^ (-0.09, 2.72) |
| **Sex** | -0.71^***^ (-1.20, -0.23) | 0.28 (-0.33, 0.90) | 0.19 (-0.43, 0.81) | 0.16 (-0.38, 0.70) | 0.41 (-0.14, 0.96) | 0.27 (-0.21, 0.75) | 0.26 (-0.23, 0.76) |
| **Age^2^** | -2.96^**^ (-5.78, -0.14) | -1.63 (-5.21, 1.94) | -0.22 (-3.83, 3.40) | -1.72 (-4.86, 1.42) | -1.89 (-5.07, 1.28) | -4.01^***^ (-6.79, -1.22) | -2.92^**^ (-5.77, -0.08) |
| **Age^3^** | 1.28^*^ (-0.16, 2.72) | 0.27 (-1.55, 2.09) | -0.38 (-2.22, 1.46) | 0.69 (-0.91, 2.29) | 0.67 (-0.95, 2.28) | 1.85^**^ (0.43, 3.27) | 1.31^*^ (-0.14, 2.76) |
| **Age x sex** | 1.12^**^ (0.09, 2.14) | -0.09 (-1.39, 1.21) | 0.12 (-1.19, 1.44) | 0.26 (-0.88, 1.40) | -0.34 (-1.49, 0.82) | -0.17 (-1.18, 0.84) | -0.12 (-1.15, 0.91) |
| **Age^2^ x sex** | -0.45 (-1.01, 0.11) | -0.18 (-0.89, 0.53) | -0.29 (-1.01, 0.42) | -0.28 (-0.90, 0.34) | 0.08 (-0.55, 0.71) | -0.05 (-0.60, 0.50) | -0.11 (-0.67, 0.45) |
| **Higher degree vs. no qualification** | -0.02 (-0.06, 0.03) | 0.03 (-0.02, 0.09) | 0.02 (-0.03, 0.08) | 0.05^**^ (0.002, 0.10) | 0.01 (-0.04, 0.06) | 0.05^**^ (0.01, 0.09) | 0.04^*^ (-0.001, 0.09) |
| **A levels vs. qualification** | 0.01 (-0.04, 0.06) | 0.01 (-0.05, 0.07) | 0.01 (-0.05, 0.08) | 0.04 (-0.01, 0.09) | -0.01 (-0.06, 0.05) | 0.05^**^ (0.0002, 0.10) | 0.03 (-0.01, 0.08) |
| **GCSEs vs. no qualification** | -0.01 (-0.05, 0.04) | -0.01 (-0.07, 0.05) | -0.02 (-0.08, 0.04) | 0.05^*^ (-0.004, 0.10) | 0.001 (-0.05, 0.05) | 0.03 (-0.01, 0.08) | 0.03 (-0.02, 0.07) |
| **CSEs vs. no qualification** | 0.03 (-0.03, 0.09) | -0.004 (-0.08, 0.08) | 0.003 (-0.08, 0.08) | 0.03 (-0.04, 0.10) | -0.01 (-0.08, 0.06) | 0.04 (-0.02, 0.10) | 0.02 (-0.05, 0.08) |
| **NVQ vs. no qualification** | -0.01 (-0.06, 0.05) | -0.003 (-0.07, 0.07) | 0.01 (-0.06, 0.08) | 0.04 (-0.02, 0.10) | -0.003 (-0.07, 0.06) | 0.002 (-0.05, 0.06) | 0.004 (-0.05, 0.06) |
| **Professional qualification vs. no qualification** | 0.05^*^ (-0.005, 0.11) | 0.01 (-0.07, 0.08) | 0.01 (-0.06, 0.09) | 0.07^**^ (0.01, 0.14) | 0.03 (-0.04, 0.10) | 0.02 (-0.04, 0.08) | 0.02 (-0.04, 0.07) |
| **Site 1** | -0.15^***^ (-0.18, -0.12) | -0.07^***^ (-0.10, -0.04) | -0.08^***^ (-0.12, -0.05) | -0.12^***^ (-0.15, -0.09) | -0.13^***^ (-0.16, -0.10) | -0.11^***^ (-0.14, -0.08) | -0.11^***^ (-0.13, -0.08) |
| **Site 2** | -0.12^***^ (-0.14, -0.10) | -0.04^***^ (-0.07, -0.01) | -0.08^***^ (-0.11, -0.05) | -0.08^***^ (-0.10, -0.05) | -0.09^***^ (-0.11, -0.06) | -0.04^***^ (-0.06, -0.01) | -0.05^***^ (-0.07, -0.02) |
| **Previous vs. never smoker** | -0.06^***^ (-0.08, -0.04) | -0.01 (-0.04, 0.02) | -0.003 (-0.03, 0.02) | -0.01 (-0.04, 0.01) | -0.02^*^ (-0.05, 0.0001) | -0.04^***^ (-0.06, -0.02) | -0.04^***^ (-0.06, -0.02) |
| **Current vs. never smoker** | -0.16^***^ (-0.19, -0.12) | -0.06^**^ (-0.11, -0.01) | -0.07^**^ (-0.12, -0.02) | -0.02 (-0.06, 0.03) | -0.03 (-0.08, 0.01) | -0.11^***^ (-0.15, -0.07) | -0.10^***^ (-0.14, -0.06) |
| **TDI** | -0.02^***^ (-0.03, -0.01) | -0.01 (-0.02, 0.003) | -0.01^**^ (-0.02, -0.001) | -0.001 (-0.01, 0.01) | -0.0001 (-0.01, 0.01) | -0.01^**^ (-0.02, -0.0004) | -0.01^**^ (-0.02, -0.002) |
| **SBP** | 0.02^***^ (0.01, 0.04) | 0.002 (-0.02, 0.02) | -0.003 (-0.02, 0.02) | 0.02^**^ (0.004, 0.04) | 0.01 (-0.01, 0.02) | 0.02^**^ (0.003, 0.03) | 0.02^**^ (0.004, 0.03) |
| **DBP** | -0.04^***^ (-0.06, -0.03) | 0.01 (-0.005, 0.03) | 0.004 (-0.01, 0.02) | -0.005 (-0.02, 0.01) | -0.002 (-0.02, 0.01) | -0.02^***^ (-0.04, -0.01) | -0.03^***^ (-0.04, -0.01) |
| **BMI** | -0.05^***^ (-0.06, -0.04) | -0.004 (-0.02, 0.01) | -0.02^**^ (-0.03, -0.002) | -0.04^***^ (-0.05, -0.03) | -0.03^***^ (-0.04, -0.02) | -0.04^***^ (-0.05, -0.03) | -0.04^***^ (-0.05, -0.03) |
| **Exercise** | 0.002 (-0.01, 0.01) | 0.01 (-0.003, 0.02) | 0.01^*^ (-0.002, 0.02) | 0.01 (-0.005, 0.02) | 0.01 (-0.003, 0.02) | 0.01^*^ (-0.001, 0.02) | 0.005 (-0.005, 0.01) |
| **Diabetes** | -0.27^***^ (-0.31, -0.22) | -0.11^***^ (-0.16, -0.05) | -0.11^***^ (-0.16, -0.05) | -0.12^***^ (-0.17, -0.07) | -0.16^***^ (-0.22, -0.11) | -0.18^***^ (-0.22, -0.13) | -0.19^***^ (-0.23, -0.14) |
| **Non-HDL cholesterol** | 0.02^***^ (0.01, 0.03) | 0.0003 (-0.01, 0.01) | 0.004 (-0.01, 0.02) | -0.003 (-0.01, 0.01) | -0.01 (-0.02, 0.003) | 0.001 (-0.01, 0.01) | -0.0005 (-0.01, 0.01) |
| **Head size** | 0.30^***^ (0.28, 0.31) | -0.37^***^ (-0.38, -0.35) | -0.32^***^ (-0.34, -0.31) | -0.47^***^ (-0.48, -0.45) | -0.45^***^ (-0.46, -0.44) | -0.62^***^ (-0.63, -0.61) | -0.61^***^ (-0.62, -0.60) |
| **Constant** | 0.10^***^ (0.06, 0.14) | 0.02 (-0.04, 0.07) | 0.03 (-0.03, 0.08) | 0.002 (-0.05, 0.05) | 0.05^**^ (0.004, 0.10) | 0.01 (-0.03, 0.05) | 0.02 (-0.02, 0.07) |
| **N** | 22,254 | 22,254 | 22,254 | 22,254 | 22,254 | 22,254 | 22,254 |
| **R^2^** | 0.51 | 0.21 | 0.19 | 0.39 | 0.38 | 0.52 | 0.50 |
| **Adjusted R^2^** | 0.51 | 0.21 | 0.19 | 0.39 | 0.38 | 0.52 | 0.50 |
| **Residual Std. Error (df = 22228)** | 0.70 | 0.89 | 0.90 | 0.78 | 0.79 | 0.69 | 0.71 |
| **F Statistic (df = 25; 22228)** | 921.30^***^ | 241.09^***^ | 214.87^***^ | 574.86^***^ | 540.86^***^ | 972.97^***^ | 896.36^***^ |
| ^*^p < .1; ^**^p < .05; ^***^p < .01 **Table 4: Regression models for grey matter IDPs.** \| **Reduced model** \| **Full model**; **Alcohol** \| -0.10^***^ (-0.11, -0.09) \| -0.10^***^ (-0.11, -0.09); **Age** \| 0.96 (-0.45, 2.36) \| 1.07 (-0.33, 2.47); **Sex** \| -0.63^**^ (-1.12, -0.15) \| -0.71^***^ (-1.20, -0.23); **Age^2^** \| -2.71^*^ (-5.54, 0.11) \| -2.96^**^ (-5.78, -0.14); **Age^3^** \| 1.16 (-0.28, 2.59) \| 1.28^*^ (-0.16, 2.72); **Age x sex** \| 0.97^*^ (-0.06, 1.99) \| 1.12^**^ (0.09, 2.14); **Age^2^ x sex** \| -0.38 (-0.94, 0.18) \| -0.45 (-1.01, 0.11); **Higher degree vs. no qualification** \| -0.02 (-0.06, 0.02) \| -0.02 (-0.06, 0.03); **A levels vs. qualification** \| 0.004 (-0.04, 0.05) \| 0.01 (-0.04, 0.06); **GCSEs vs. no qualification** \| -0.01 (-0.06, 0.04) \| -0.01 (-0.05, 0.04); **CSEs vs. no qualification** \| 0.03 (-0.03, 0.09) \| 0.03 (-0.03, 0.09); **NVQ vs. no qualification** \| -0.01 (-0.07, 0.05) \| -0.01 (-0.06, 0.05); **Professional qualification vs. no qualification** \| 0.05^*^ (-0.01, 0.11) \| 0.05^*^ (-0.005, 0.11); **Site 1** \| -0.15^***^ (-0.18, -0.12) \| -0.15^***^ (-0.18, -0.12); **Site 2** \| -0.12^***^ (-0.14, -0.10) \| -0.12^***^ (-0.14, -0.10); **Previous vs. never smoker** \| -0.06^***^ (-0.08, -0.04) \| -0.06^***^ (-0.08, -0.04); **Current vs. never smoker** \| -0.15^***^ (-0.19, -0.11) \| -0.16^***^ (-0.19, -0.12); **TDI** \| -0.02^***^ (-0.03, -0.01) \| -0.02^***^ (-0.03, -0.01); **Exercise** \| 0.003 (-0.01, 0.01) \| 0.002 (-0.01, 0.01); **Diabetes** \| -0.27^***^ (-0.32, -0.23) \| -0.27^***^ (-0.31, -0.22); **Head size** \| 0.30^***^ (0.28, 0.31) \| 0.30^***^ (0.28, 0.31); **BMI** \| -0.06^***^ (-0.07, -0.05) \| -0.05^***^ (-0.06, -0.04); **SBP** \| \| 0.02^***^ (0.01, 0.04); **DBP** \| \| -0.04^***^ (-0.06, -0.03); **Non-HDL cholesterol** \| \| 0.02^***^ (0.01, 0.03); **Constant** \| 0.10^***^ (0.06, 0.14) \| 0.10^***^ (0.06, 0.14); **N** \| 22,254 \| 22,254; **R^2^** \| 0.51 \| 0.51; **Adjusted R^2^** \| 0.51 \| 0.51; **Residual Std. Error** \| 0.70 (df = 22231) \| 0.70 (df = 22228); **F Statistic** \| 1,041.75^***^ (df = 22; 22231) \| 921.30^***^ (df = 25; 22228); ^*^p < .1; ^**^p < .05; ^***^p < .01 \| \|; **Table 5: Regression models predicting grey matter volume, with and without cardiovascular risk factors affected by alcohol.** \| \| | | | | | | | |

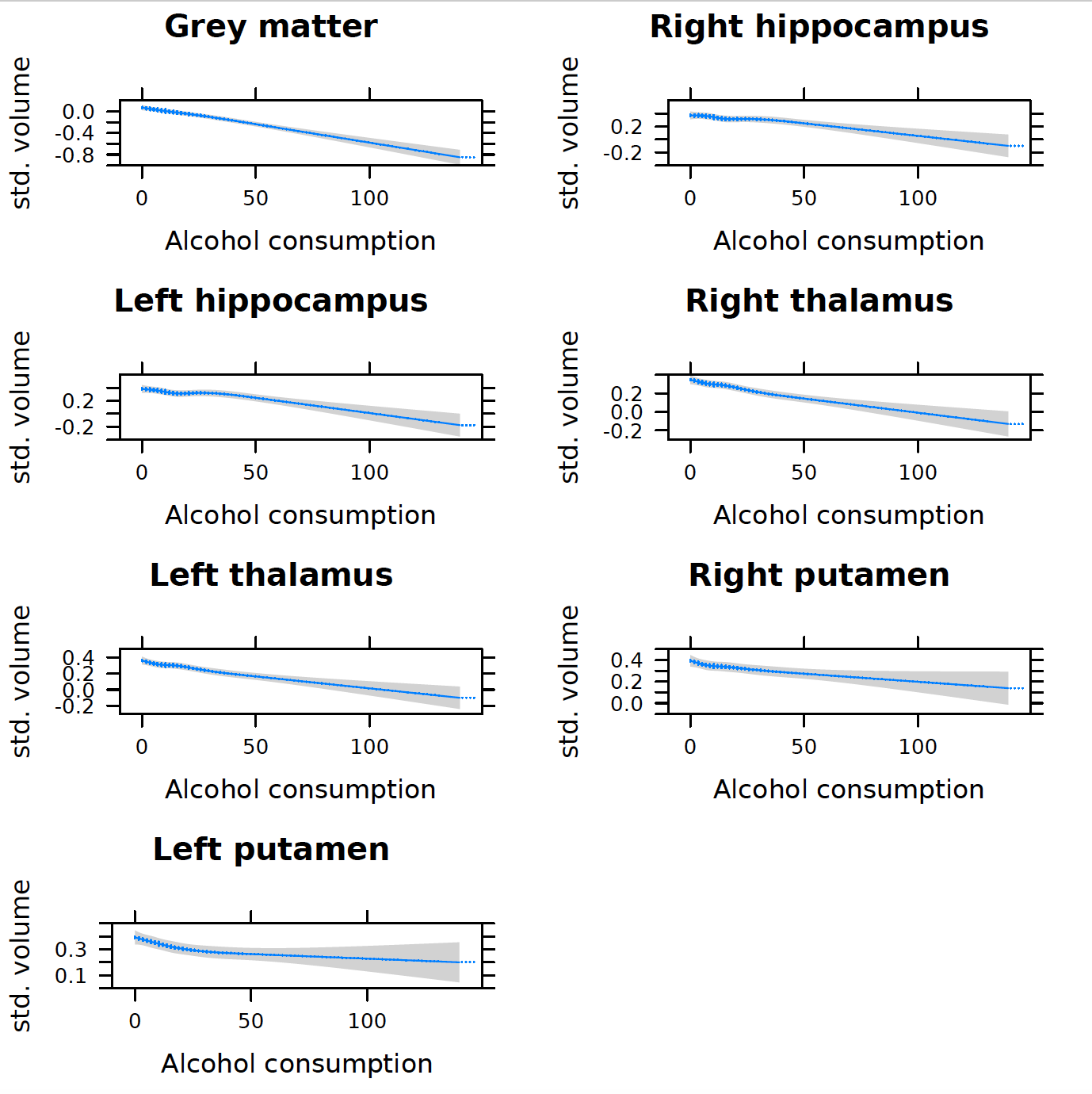

**a) Excluding previous drinkers**

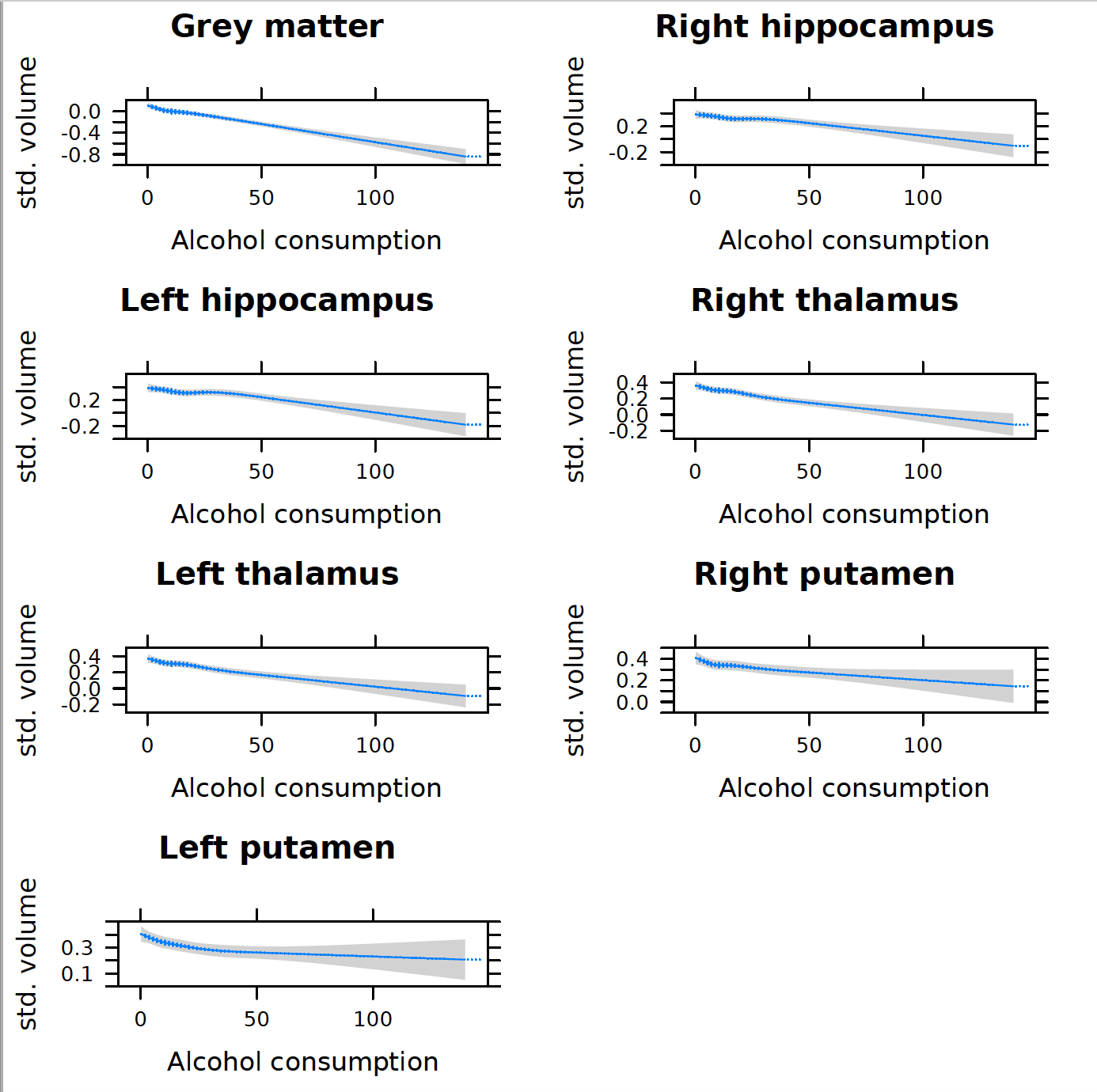

**b) Excluding all non-drinkers**

**Figure 8: Predicted change in selected brain volumes according to weekly alcohol intake in units (grams conversion: 50u=400g, 100u=800g) for excluding a) previous drinkers b) previous and never drinkers. Predictions are based on regression models with alcohol (restricted cubic splines fitted with knots at 5^th^, 25^th^, 50^th^, 75^th^ and 95^th^ percentiles) as an independent variable and standardized brain volume as the dependent variable with n=22,254 subjects. Models are adjusted for: age, sex, age^2^, age^3^, age x sex, age^2^ x sex, SBP, DBP, TDI, smoking, BMI, non-HDL cholesterol, Diabetes Mellitus, head size, exercise. 95% confidence intervals are shaded.**

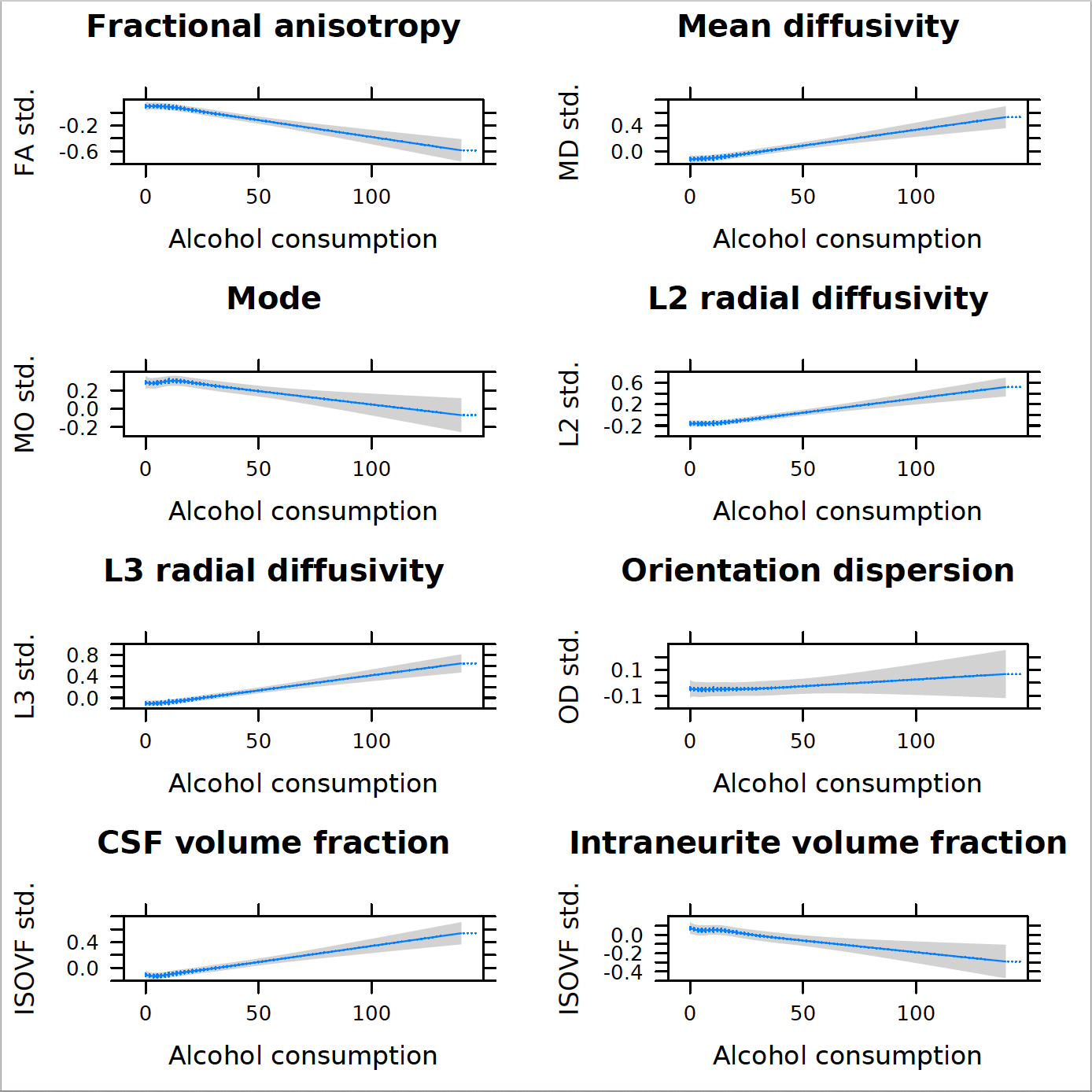

**Abbreviations: FA – fractional anisotropy, MD – mean diffusivity, MO – mode, L2 – radial diffusivity (eigenvalue L2), L3 – radial diffusivity (eigenvalue L3), OD – orientation dispersion, isovf – isotropic volume fraction, icvf – intracellular volume fraction, std. – standardized (z-score).**

**Figure 9: Predicted change in corpus callosum (genu) diffusion metrics according to weekly alcohol intake in units (grams conversion: 50u=400g, 100u=800g). Predictions are based on regression models with alcohol (spline fit with knots at 5^th^, 25^th^, 50^th^, 75^th^ and 95^th^ percentiles) as an independent variable and standardized diffusion metric as the dependent variable with n=22,254 subjects. Models are adjusted for: age, sex, age^2^, age^3^, age x sex, age^2^ x sex, SBP, DBP, TDI, smoking, BMI, non-HDL cholesterol, Diabetes Mellitus, head size, exercise. 95% confidence intervals are shaded.**

|  | **Fractional anisotropy** | **Mean diffusivity** | **Mode** | **L2 radial diffusivity** | **L3 radial diffusivity** | **Intracellular volume fraction** | **Isotropic volume fraction** | **Orientation dispersion** |
| --- | --- | --- | --- | --- | --- | --- | --- | --- |
| **Alcohol** | -0.08^***^ (-0.09, -0.06) | 0.07^***^ (0.06, 0.08) | -0.03^***^ (-0.05, -0.02) | 0.07^***^ (0.06, 0.09) | 0.08^***^ (0.07, 0.10) | -0.04^***^ (-0.06, -0.03) | 0.07^***^ (0.06, 0.09) | 0.01 (-0.01, 0.02) |
| **Age** | -0.37 (-2.20, 1.45) | -1.24 (-3.03, 0.54) | -3.85^***^ (-5.79, -1.92) | 0.70 (-1.12, 2.53) | -0.70 (-2.47, 1.07) | 1.28 (-0.63, 3.20) | -0.70 (-2.50, 1.10) | 3.98^***^ (2.02, 5.94) |
| **Sex** | 0.04 (-0.59, 0.68) | 0.12 (-0.51, 0.74) | -0.23 (-0.90, 0.44) | 0.26 (-0.37, 0.90) | -0.13 (-0.75, 0.49) | -0.40 (-1.07, 0.27) | -0.16 (-0.79, 0.46) | -0.25 (-0.93, 0.43) |
| **Age^2^** | 1.41 (-2.27, 5.08) | 1.67 (-1.93, 5.27) | 8.90^***^ (5.00, 12.81) | -2.31 (-5.99, 1.37) | 0.72 (-2.86, 4.29) | -2.69 (-6.55, 1.17) | 0.19 (-3.45, 3.83) | -8.37^***^ (-12.31, -4.42) |
| **Age^3^** | -1.39 (-3.27, 0.48) | -0.03 (-1.86, 1.80) | -5.18^***^ (-7.16, -3.19) | 1.96^**^ (0.08, 3.83) | 0.40 (-1.42, 2.22) | 1.14 (-0.82, 3.11) | 0.83 (-1.02, 2.69) | 4.44^***^ (2.43, 6.44) |
| **Age x sex** | -0.11 (-1.45, 1.22) | -0.20 (-1.51, 1.11) | 0.94 (-0.48, 2.36) | -0.65 (-1.99, 0.68) | 0.36 (-0.94, 1.66) | 0.73 (-0.67, 2.13) | 0.28 (-1.04, 1.60) | 0.41 (-1.03, 1.84) |
| **Age^2^ x sex** | 0.10 (-0.63, 0.83) | 0.09 (-0.63, 0.80) | -0.70^*^ (-1.47, 0.07) | 0.36 (-0.37, 1.09) | -0.24 (-0.95, 0.47) | -0.29 (-1.05, 0.48) | -0.07 (-0.80, 0.65) | -0.20 (-0.98, 0.59) |
| **Higher degree vs. no qualification** | -0.05^*^ (-0.11, 0.002) | 0.04 (-0.01, 0.10) | -0.01 (-0.07, 0.05) | 0.04 (-0.02, 0.09) | 0.06^**^ (0.004, 0.11) | -0.06^*^ (-0.12, 0.001) | 0.02 (-0.04, 0.07) | 0.002 (-0.06, 0.06) |
| **A levels vs. qualification** | -0.04 (-0.10, 0.02) | 0.04 (-0.02, 0.10) | -0.02 (-0.09, 0.05) | 0.04 (-0.03, 0.10) | 0.04 (-0.02, 0.10) | -0.04 (-0.11, 0.03) | 0.02 (-0.05, 0.08) | 0.0003 (-0.07, 0.07) |
| **GCSEs vs. no qualification** | -0.04 (-0.10, 0.02) | 0.04 (-0.02, 0.10) | 0.0002 (-0.06, 0.06) | 0.04 (-0.02, 0.10) | 0.05 (-0.01, 0.10) | -0.05 (-0.11, 0.01) | 0.01 (-0.05, 0.07) | -0.02 (-0.08, 0.04) |
| **CSEs vs. no qualification** | 0.04 (-0.04, 0.12) | -0.01 (-0.09, 0.07) | 0.05 (-0.04, 0.13) | -0.04 (-0.12, 0.04) | -0.02 (-0.10, 0.06) | -0.01 (-0.09, 0.08) | -0.03 (-0.11, 0.05) | -0.06 (-0.15, 0.03) |
| **NVQ vs. no qualification** | -0.02 (-0.09, 0.05) | 0.03 (-0.04, 0.10) | -0.06 (-0.14, 0.02) | 0.02 (-0.05, 0.10) | 0.03 (-0.04, 0.10) | 0.02 (-0.06, 0.09) | 0.05 (-0.02, 0.12) | 0.04 (-0.04, 0.12) |
| **Professional qualification vs. no qualification** | -0.08^*^ (-0.15, 0.0003) | 0.07^*^ (-0.01, 0.14) | -0.04 (-0.12, 0.04) | 0.07^*^ (-0.01, 0.15) | 0.08^**^ (0.003, 0.15) | -0.08^*^ (-0.16, 0.0003) | 0.03 (-0.04, 0.11) | 0.004 (-0.08, 0.09) |
| **Site 1** | 0.36^***^ (0.33, 0.40) | -0.22^***^ (-0.26, -0.19) | 0.06^***^ (0.02, 0.10) | -0.33^***^ (-0.36, -0.29) | -0.29^***^ (-0.32, -0.26) | -0.12^***^ (-0.16, -0.09) | -0.44^***^ (-0.48, -0.41) | -0.16^***^ (-0.20, -0.12) |
| **Site 2** | 0.03^**^ (0.003, 0.06) | -0.08^***^ (-0.11, -0.05) | 0.04^**^ (0.01, 0.07) | -0.09^***^ (-0.12, -0.06) | -0.03^**^ (-0.06, -0.004) | -0.03^*^ (-0.06, 0.0001) | -0.18^***^ (-0.21, -0.15) | -0.02 (-0.06, 0.01) |
| **Previous vs. never smoker** | -0.04^***^ (-0.06, -0.01) | 0.04^***^ (0.01, 0.06) | 0.004 (-0.02, 0.03) | 0.04^***^ (0.01, 0.06) | 0.04^***^ (0.01, 0.07) | -0.04^***^ (-0.07, -0.01) | 0.02 (-0.01, 0.05) | -0.004 (-0.03, 0.03) |
| **Current vs. never smoker** | -0.14^***^ (-0.19, -0.09) | 0.14^***^ (0.09, 0.19) | -0.005 (-0.06, 0.05) | 0.14^***^ (0.09, 0.19) | 0.16^***^ (0.11, 0.21) | -0.09^***^ (-0.14, -0.04) | 0.14^***^ (0.09, 0.19) | 0.01 (-0.04, 0.07) |
| **TDI** | -0.01 (-0.02, 0.01) | 0.001 (-0.01, 0.01) | 0.01 (-0.01, 0.02) | 0.001 (-0.01, 0.01) | 0.01 (-0.005, 0.02) | -0.002 (-0.02, 0.01) | 0.001 (-0.01, 0.01) | 0.002 (-0.01, 0.01) |
| **SBP** | 0.005 (-0.01, 0.02) | 0.02^*^ (-0.0004, 0.04) | 0.01 (-0.01, 0.03) | 0.01 (-0.01, 0.03) | -0.001 (-0.02, 0.02) | -0.01 (-0.03, 0.01) | 0.01 (-0.01, 0.03) | -0.02^**^ (-0.04, -0.003) |
| **DBP** | -0.07^***^ (-0.09, -0.05) | 0.05^***^ (0.03, 0.07) | -0.02^**^ (-0.04, -0.003) | 0.06^***^ (0.04, 0.08) | 0.06^***^ (0.04, 0.08) | -0.05^***^ (-0.07, -0.03) | 0.03^***^ (0.01, 0.05) | 0.02 (-0.003, 0.04) |
| **BMI** | -0.07^***^ (-0.08, -0.05) | 0.02^***^ (0.01, 0.03) | -0.04^***^ (-0.05, -0.03) | 0.05^***^ (0.04, 0.07) | 0.05^***^ (0.04, 0.06) | 0.03^***^ (0.01, 0.04) | 0.06^***^ (0.05, 0.07) | 0.07^***^ (0.05, 0.08) |
| **Exercise** | 0.01^**^ (0.002, 0.03) | -0.01^**^ (-0.03, -0.003) | 0.004 (-0.01, 0.02) | -0.01^**^ (-0.03, -0.002) | -0.02^***^ (-0.03, -0.004) | 0.005 (-0.01, 0.02) | -0.02^***^ (-0.03, -0.01) | -0.003 (-0.02, 0.01) |
| **Diabetes** | -0.24^***^ (-0.30, -0.18) | 0.25^***^ (0.19, 0.31) | -0.16^***^ (-0.22, -0.09) | 0.26^***^ (0.20, 0.32) | 0.26^***^ (0.20, 0.31) | -0.19^***^ (-0.25, -0.12) | 0.20^***^ (0.14, 0.26) | 0.04 (-0.02, 0.10) |
| **Non-HDL cholesterol** | 0.03^***^ (0.02, 0.04) | -0.03^***^ (-0.04, -0.02) | 0.01 (-0.01, 0.02) | -0.03^***^ (-0.04, -0.02) | -0.03^***^ (-0.04, -0.02) | 0.03^***^ (0.01, 0.04) | -0.02^**^ (-0.03, -0.004) | -0.01 (-0.02, 0.01) |
| **Head size** | -0.06^***^ (-0.08, -0.05) | -0.06^***^ (-0.08, -0.05) | -0.14^***^ (-0.16, -0.12) | 0.03^***^ (0.01, 0.04) | -0.004 (-0.02, 0.01) | 0.09^***^ (0.07, 0.11) | 0.04^***^ (0.02, 0.05) | 0.16^***^ (0.15, 0.18) |
| **Constant** | 0.01 (-0.04, 0.07) | -0.02 (-0.07, 0.04) | -0.001 (-0.06, 0.06) | 0.003 (-0.05, 0.06) | -0.03 (-0.08, 0.02) | 0.10^***^ (0.04, 0.16) | 0.07^**^ (0.01, 0.12) | 0.03 (-0.03, 0.09) |
| **N** | 22,254 | 22,254 | 22,254 | 22,254 | 22,254 | 22,254 | 22,254 | 22,254 |
| **R^2^** | 0.17 | 0.20 | 0.06 | 0.16 | 0.21 | 0.08 | 0.18 | 0.04 |
| **Adjusted R^2^** | 0.17 | 0.20 | 0.06 | 0.16 | 0.21 | 0.08 | 0.18 | 0.04 |
| **Residual Std. Error (df = 22228)** | 0.91 | 0.89 | 0.97 | 0.91 | 0.89 | 0.96 | 0.90 | 0.98 |
| **F Statistic (df = 25; 22228)** | 178.35^***^ | 225.21^***^ | 58.44^***^ | 174.88^***^ | 238.85^***^ | 79.11^***^ | 201.38^***^ | 36.94^***^ |
| ^*^p < .1; ^**^p < .05; ^***^p < .01 | | | | | | | | |

**Table 6: Regression model for diffusion tensor imaging IDPs.**

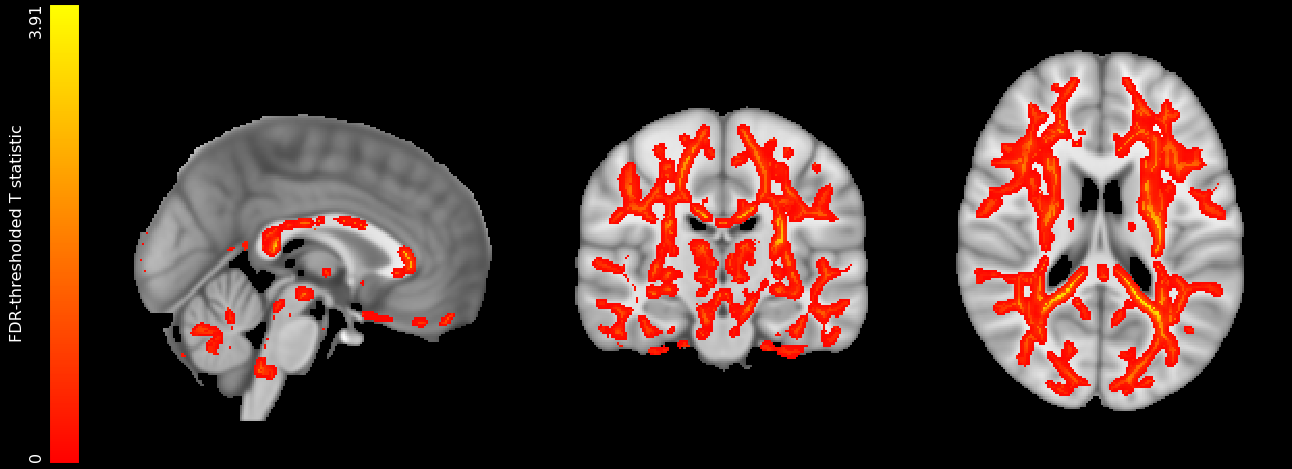

**Figure 10: Positive association between alcohol consumption and mean diffusivity. N=24030 subjects. Red-yellow voxels indicate FDR-thresholded T statistics. Adjusted for: age, sex, age^2^, age^3^, age x sex, age^2^ x sex, imaging site, SBP, DBP, cholesterol, HDL, Diabetes Mellitus, smoking, BMI, exercise, TDI.**

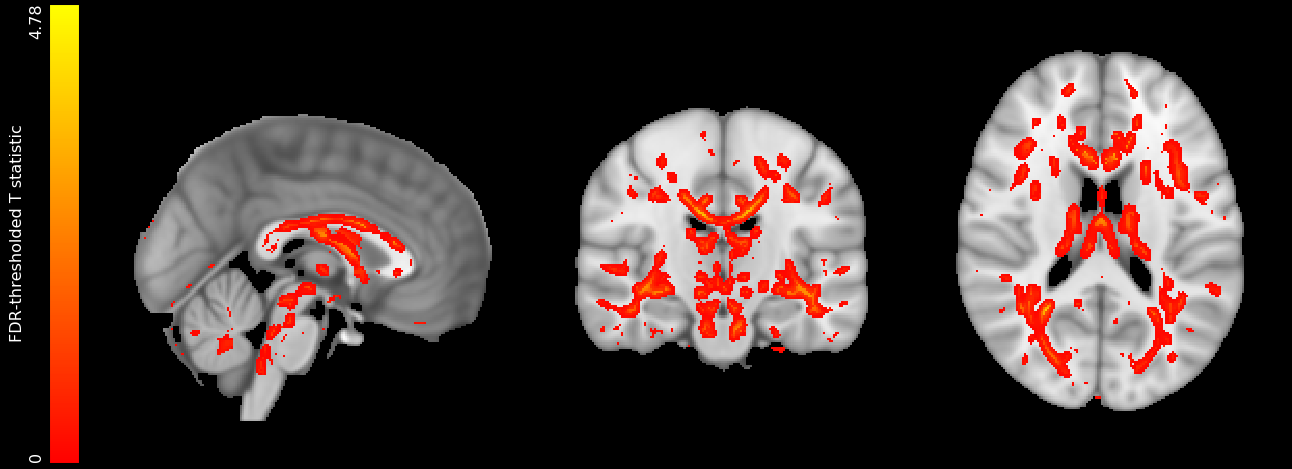

**Figure 11: Negative association between alcohol consumption and mode. N=24030 subjects. Red-yellow voxels indicate FDR-thresholded T statistics. Adjusted for: age, sex, age^2^, age^3^, age x sex, age^2^ x sex, imaging site, SBP, DBP, cholesterol, HDL, Diabetes Mellitus, smoking, BMI, exercise, TDI.**

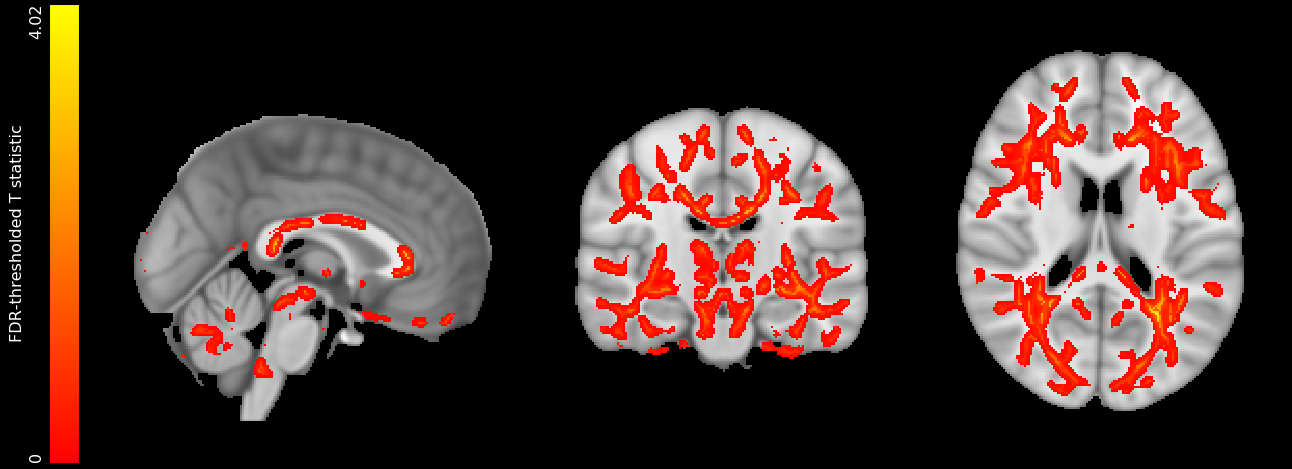

**Figure 12: Positive association between alcohol consumption and L2 (radial diffusivity). N=24030. Red-yellow voxels indicate FDR-thresholded T statistics. Adjusted for: age, sex, age^2^, age^3^, age x sex, age^2^ x sex, imaging site, SBP, DBP, cholesterol, HDL, Diabetes Mellitus, smoking, BMI, exercise, TDI.**

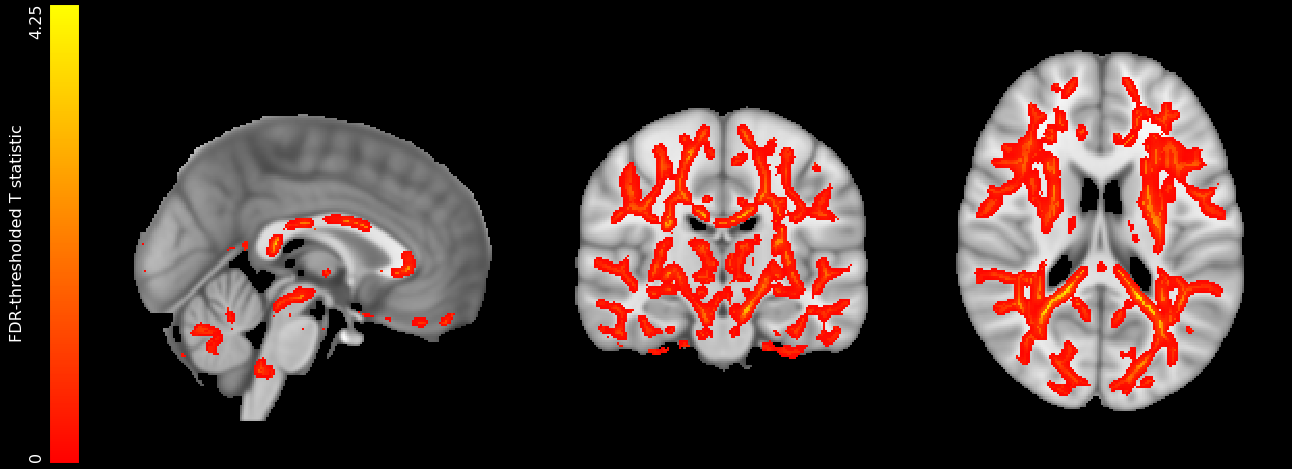

**Figure 13: Positive association between alcohol consumption and L3 (radial diffusivity). N=24030. Red-yellow voxels indicate FDR-thresholded T statistics. Adjusted for: age, sex, age^2^, age^3^, age x sex, age^2^ x sex, imaging site, SBP, DBP, cholesterol, HDL, Diabetes Mellitus, smoking, BMI, exercise, TDI.**

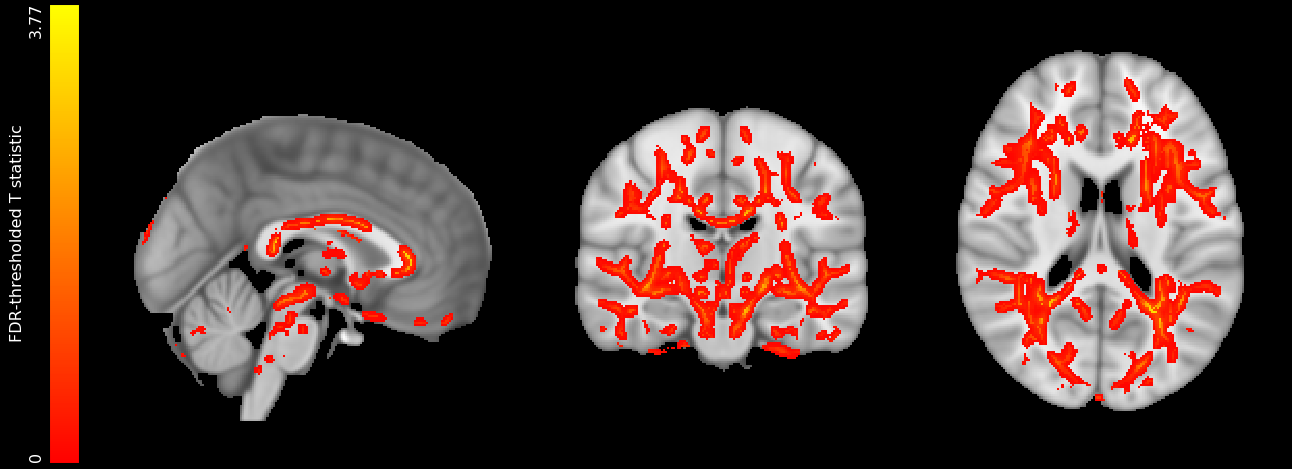

**Figure 14: Additional adjustment for image-related confounders. Negative association between alcohol consumption (weekly units) and fractional anisotropy. Red-yellow voxels indicate FDR-thresholded T statistics. Adjusted for: age, sex, age^2^, age^3^, age x sex, age^2^ x sex, imaging site, SBP, DBP, cholesterol, HDL, Diabetes Mellitus, smoking, BMI, exercise, TDI, table position, head motion during structural scan, acquisition parameters. N=22263.**

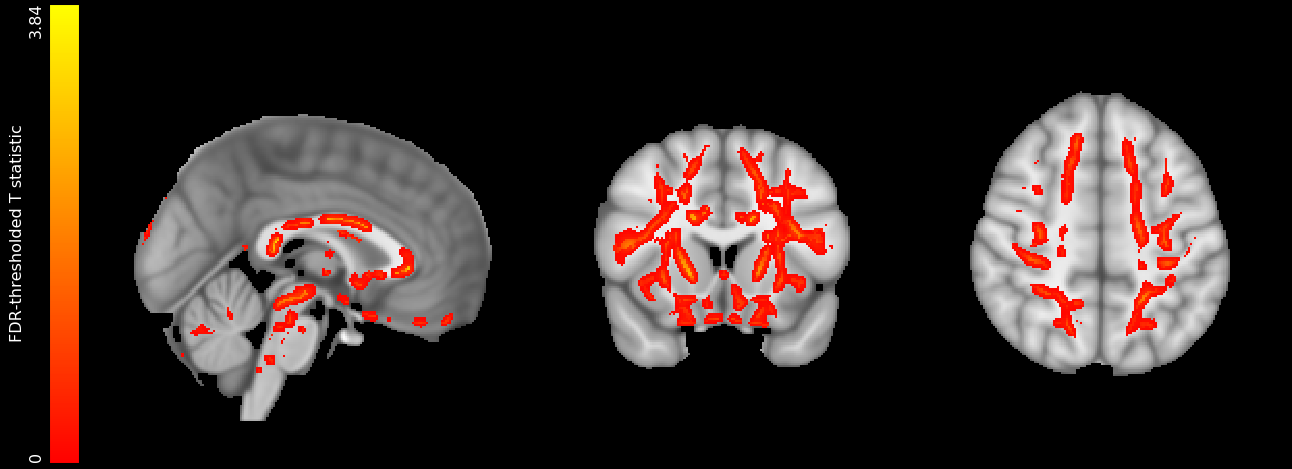

**Figure 15: Negative associations between fractional anisotropy and alcohol intake for current drinkers only (n=22809). Red-yellow voxels indicate FDR-thresholded T statistics. Adjusted for: age, sex, age^2^, age^3^, age x sex, age^2^ x sex, imaging site, SBP, DBP, cholesterol, HDL, Diabetes Mellitus, smoking, BMI, exercise, TDI.**

|  | **Alcohol-BMI interaction** | **Alcohol-SBP interaction** | **Alcohol-DBP interaction** |
| --- | --- | --- | --- |
| **Alcohol** | -0.09^***^ (-0.10, -0.08) | -0.09^***^ (-0.10, -0.08) | -0.09^***^ (-0.10, -0.08) |
| **Age** | -0.05^***^ (-0.06, -0.04) | -0.05^***^ (-0.06, -0.04) | -0.05^***^ (-0.06, -0.04) |
| **Sex** | 1.08 (-0.32, 2.48) | 1.03 (-0.37, 2.43) | 1.06 (-0.34, 2.46) |
| **Age^2^** | -0.73^***^ (-1.21, -0.24) | -0.72^***^ (-1.21, -0.24) | -0.74^***^ (-1.23, -0.25) |
| **Age^3^** | -2.99^**^ (-5.81, -0.16) | -2.89^**^ (-5.71, -0.07) | -2.95^**^ (-5.78, -0.13) |
| **Age x sex** | 1.30^*^ (-0.14, 2.73) | 1.25^*^ (-0.19, 2.68) | 1.28^*^ (-0.15, 2.72) |
| **Age^2^ x sex** | 1.14^**^ (0.11, 2.17) | 1.13^**^ (0.10, 2.16) | 1.16^**^ (0.14, 2.19) |
| **Higher degree vs. no qualification** | -0.46 (-1.02, 0.10) | -0.45 (-1.01, 0.11) | -0.47^*^ (-1.03, 0.09) |
| **A levels vs. qualification** | -0.02 (-0.06, 0.02) | -0.02 (-0.06, 0.03) | -0.02 (-0.06, 0.03) |
| **GCSEs vs. no qualification** | 0.01 (-0.04, 0.06) | 0.01 (-0.04, 0.06) | 0.01 (-0.04, 0.06) |
| **CSEs vs. no qualification** | -0.01 (-0.05, 0.04) | -0.01 (-0.05, 0.04) | -0.01 (-0.05, 0.04) |
| **NVQ vs. no qualification** | 0.03 (-0.03, 0.09) | 0.03 (-0.03, 0.10) | 0.03 (-0.03, 0.09) |
| **Professional qualification vs. no qualification** | -0.01 (-0.06, 0.05) | -0.01 (-0.06, 0.05) | -0.01 (-0.06, 0.05) |
| **Site 1** | 0.05^*^ (-0.01, 0.11) | 0.05^*^ (-0.005, 0.11) | 0.05^*^ (-0.01, 0.11) |
| **Site 2** | -0.15^***^ (-0.18, -0.12) | -0.15^***^ (-0.18, -0.12) | -0.15^***^ (-0.18, -0.12) |
| **Previous vs. never smoker** | -0.12^***^ (-0.14, -0.10) | -0.12^***^ (-0.14, -0.10) | -0.12^***^ (-0.14, -0.10) |
| **Current vs. never smoker** | -0.06^***^ (-0.08, -0.04) | -0.06^***^ (-0.08, -0.04) | -0.06^***^ (-0.08, -0.04) |
| **TDI** | -0.16^***^ (-0.20, -0.12) | -0.16^***^ (-0.20, -0.12) | -0.16^***^ (-0.20, -0.12) |
| **SBP** | -0.02^***^ (-0.03, -0.01) | -0.02^***^ (-0.03, -0.01) | -0.02^***^ (-0.03, -0.01) |
| **DBP** | 0.02^***^ (0.01, 0.04) | 0.02^***^ (0.01, 0.04) | 0.02^***^ (0.01, 0.04) |
| **BMI** | -0.04^***^ (-0.06, -0.03) | -0.05^***^ (-0.06, -0.03) | -0.04^***^ (-0.06, -0.03) |
| **Exercise** | 0.002 (-0.01, 0.01) | 0.002 (-0.01, 0.01) | 0.002 (-0.01, 0.01) |
| **Diabetes** | -0.27^***^ (-0.31, -0.22) | -0.27^***^ (-0.31, -0.22) | -0.27^***^ (-0.31, -0.22) |
| **Non-HDL cholesterol** | 0.02^***^ (0.01, 0.03) | 0.02^***^ (0.01, 0.03) | 0.02^***^ (0.01, 0.03) |
| **Head size** | 0.30^***^ (0.28, 0.31) | 0.30^***^ (0.28, 0.31) | 0.30^***^ (0.28, 0.31) |
| **Alcohol x BMI** | -0.01^**^ (-0.02, -0.002) |  |  |
| **Alcohol x SBP** |  | -0.01^***^ (-0.02, -0.004) |  |
| **Alcohol x DBP** |  |  | -0.01^***^ (-0.02, -0.004) |
| **Constant** | 0.10^***^ (0.06, 0.14) | 0.10^***^ (0.06, 0.14) | 0.10^***^ (0.06, 0.14) |
| **N** | 22,254 | 22,254 | 22,254 |
| **R^2^** | 0.51 | 0.51 | 0.51 |
| **Adjusted R^2^** | 0.51 | 0.51 | 0.51 |
| **Residual Std. Error (df = 22227)** | 0.70 | 0.70 | 0.70 |
| **F Statistic (df = 26; 22227)** | 886.24^***^ | 886.44^***^ | 886.42^***^ |
| ^*^p < .1; ^**^p < .05; ^***^p < .01 | | | |

**Table 7: Regression models for significant interactions with alcohol intake in predicting grey matter volume.**

|  | **Alcohol x age** | **Alcohol x sex** | **Alcohol x Apoe4** | **Alcohol x heart disease** | **Alcohol x GGT** |
| --- | --- | --- | --- | --- | --- |
| **Alcohol** | -0.095^***^ | -0.094^***^ | -0.097^***^ | -0.093^***^ | -0.093^***^ |
|  | (0.005) | (0.006) | (0.006) | (0.005) | (0.007) |
| **Apoe4 one** |  |  | -0.018 |  |  |
|  |  |  | (0.011) |  |  |
| **Apoe4 two** |  |  | 0.040 |  |  |
|  |  |  | (0.032) |  |  |
| **Heart disease** |  |  |  | -0.067^***^ |  |
|  |  |  |  | (0.018) |  |
| **GGT** |  |  |  |  | -0.0004^***^ |
|  |  |  |  |  | (0.0002) |
| **Age** | 1.101 | 1.150 | 1.147 | 1.132 | 1.134 |
|  | (0.718) | (0.717) | (0.717) | (0.717) | (0.717) |
| **Sex** | -0.706^***^ | -0.704^***^ | -0.699^***^ | -0.698^***^ | -0.701^***^ |
|  | (0.250) | (0.250) | (0.250) | (0.250) | (0.250) |
| **Age^2^** | -3.028^**^ | -3.125^**^ | -3.116^**^ | -3.087^**^ | -3.090^**^ |
|  | (1.447) | (1.446) | (1.446) | (1.446) | (1.447) |
| **Age^3^** | 1.315^*^ | 1.367^*^ | 1.361^*^ | 1.349^*^ | 1.347^*^ |
|  | (0.737) | (0.736) | (0.736) | (0.736) | (0.737) |
| **Age x sex** | 1.078^**^ | 1.093^**^ | 1.081^**^ | 1.077^**^ | 1.092^**^ |
|  | (0.526) | (0.526) | (0.526) | (0.526) | (0.526) |
| **Age^2^ x sex** | -0.415 | -0.434 | -0.427 | -0.421 | -0.434 |
|  | (0.287) | (0.287) | (0.287) | (0.287) | (0.287) |
| **Higher degree vs. no qualification** | -0.017 | -0.018 | -0.018 | -0.020 | -0.020 |
|  | (0.022) | (0.022) | (0.022) | (0.022) | (0.022) |
| **A levels vs. qualification** | 0.008 | 0.008 | 0.008 | 0.006 | 0.006 |
|  | (0.025) | (0.025) | (0.025) | (0.025) | (0.025) |
| **GCSEs vs. no qualification** | -0.004 | -0.004 | -0.005 | -0.006 | -0.006 |
|  | (0.024) | (0.024) | (0.024) | (0.024) | (0.024) |
| **CSEs vs. no qualification** | 0.033 | 0.033 | 0.033 | 0.033 | 0.031 |
|  | (0.032) | (0.032) | (0.032) | (0.032) | (0.032) |
| **NVQ vs. no qualification** | -0.002 | -0.002 | -0.001 | -0.001 | -0.003 |
|  | (0.029) | (0.029) | (0.029) | (0.029) | (0.029) |
| **Professional qualification vs. no qualification** | 0.056^*^ | 0.056^*^ | 0.056^*^ | 0.056^*^ | 0.054^*^ |
|  | (0.030) | (0.030) | (0.030) | (0.030) | (0.030) |
| **Site 1** | -0.150^***^ | -0.150^***^ | -0.150^***^ | -0.151^***^ | -0.150^***^ |
|  | (0.014) | (0.014) | (0.014) | (0.014) | (0.014) |
| **Site 2** | -0.118^***^ | -0.118^***^ | -0.118^***^ | -0.118^***^ | -0.118^***^ |
|  | (0.012) | (0.012) | (0.012) | (0.012) | (0.012) |
| **Previous vs. never smoker** | -0.064^***^ | -0.065^***^ | -0.064^***^ | -0.064^***^ | -0.064^***^ |
|  | (0.011) | (0.011) | (0.011) | (0.011) | (0.011) |
| **Current vs. never smoker** | -0.158^***^ | -0.157^***^ | -0.157^***^ | -0.155^***^ | -0.158^***^ |
|  | (0.020) | (0.020) | (0.020) | (0.020) | (0.020) |
| **TDI** | -0.016^***^ | -0.016^***^ | -0.016^***^ | -0.015^***^ | -0.016^***^ |
|  | (0.005) | (0.005) | (0.005) | (0.005) | (0.005) |
| **SBP** | 0.020^***^ | 0.020^***^ | 0.020^***^ | 0.020^***^ | 0.021^***^ |
|  | (0.007) | (0.007) | (0.007) | (0.007) | (0.007) |
| **DBP** | -0.044^***^ | -0.044^***^ | -0.045^***^ | -0.045^***^ | -0.044^***^ |
|  | (0.007) | (0.007) | (0.007) | (0.007) | (0.007) |
| **BMI** | -0.052^***^ | -0.052^***^ | -0.052^***^ | -0.050^***^ | -0.050^***^ |
|  | (0.005) | (0.005) | (0.005) | (0.005) | (0.005) |
| **Exercise** | 0.001 | 0.001 | 0.001 | 0.001 | 0.001 |
|  | (0.005) | (0.005) | (0.005) | (0.005) | (0.005) |
| **Diabetes** | -0.268^***^ | -0.268^***^ | -0.268^***^ | -0.265^***^ | -0.264^***^ |
|  | (0.023) | (0.023) | (0.023) | (0.023) | (0.023) |
| **Non HDL cholesterol** | 0.022^***^ | 0.022^***^ | 0.022^***^ | 0.021^***^ | 0.023^***^ |
|  | (0.005) | (0.005) | (0.005) | (0.005) | (0.005) |
| **Head size** | 0.296^***^ | 0.296^***^ | 0.296^***^ | 0.296^***^ | 0.297^***^ |
|  | (0.006) | (0.006) | (0.006) | (0.006) | (0.006) |
| **Alcohol x age** | -0.009^*^ |  |  |  |  |
|  | (0.005) |  |  |  |  |
| **Alcohol x sex** |  | -0.002 |  |  |  |
|  |  | (0.006) |  |  |  |
| **Alcohol x Apoe 1** |  |  | 0.006 |  |  |
|  |  |  | (0.011) |  |  |
| **Alcohol x Apoe 2** |  |  | 0.052 |  |  |
|  |  |  | (0.033) |  |  |
| **Alcohol x heart disease** |  |  |  | -0.018 |  |
|  |  |  |  | (0.016) |  |
| **Alcohol x GGT** |  |  |  |  | 0.00001 |
|  |  |  |  |  | (0.0001) |
| **Constant** | 0.098^***^ | 0.099^***^ | 0.102^***^ | 0.105^***^ | 0.115^***^ |
|  | (0.022) | (0.022) | (0.022) | (0.022) | (0.023) |
| **Observations** | 22,007 | 22,007 | 22,007 | 22,007 | 21,993 |
| **R^2^** | 0.510 | 0.509 | 0.510 | 0.510 | 0.510 |
| **Adjusted R^2^** | 0.509 | 0.509 | 0.509 | 0.509 | 0.509 |
| **Residual Std. Error** | 0.701 (df = 21980) | 0.701 (df = 21980) | 0.701 (df = 21977) | 0.701 (df = 21979) | 0.701 (df = 21965) |
| **F Statistic** | 878.252^***^ (df = 26; 21980) | 878.029^***^ (df = 26; 21980) | 787.592^***^ (df = 29; 21977) | 846.638^***^ (df = 27; 21979) | 845.480^***^ (df = 27; 21965) |
| *Note:* | ^*^p<0.1; ^**^p<0.05; ^***^p<0.01 | | | | |

**Table 8: Regression models for non-significant interactions with alcohol in predicting grey matter volume.**

|  | **Quantile 2 drinkers** | **Quantile 3 drinkers** | **Quantile 4 drinkers** | **Quantile 5 drinkers** |
| --- | --- | --- | --- | --- |
| **Alcohol total weekly volume** | -0.02^**^ (-0.03, -0.004) | -0.02^**^ (-0.03, -0.004) | 0.01 (-0.005, 0.01) | -0.01^***^ (-0.01, -0.004) |
| **<Monthly binge** | -0.02 (-0.09, 0.04) | -0.02 (-0.09, 0.04) | -0.12^***^ (-0.20, -0.05) | -0.05 (-0.15, 0.05) |
| **Monthly binge** | -0.01 (-0.10, 0.08) | -0.01 (-0.10, 0.08) | -0.13^***^ (-0.22, -0.05) | -0.01 (-0.12, 0.09) |
| **Weekly binge** | -0.01 (-0.10, 0.08) | -0.01 (-0.10, 0.08) | -0.07^*^ (-0.15, 0.01) | -0.05 (-0.14, 0.04) |
| **Daily binge** | -0.23 (-0.58, 0.12) | -0.23 (-0.58, 0.12) | -0.20^**^ (-0.36, -0.04) | -0.19^***^ (-0.30, -0.08) |
| **SBP** | 0.002^*^ (-0.0002, 0.004) | 0.002^*^ (-0.0002, 0.004) | -0.0004 (-0.003, 0.002) | 0.002 (-0.001, 0.004) |
| **Age** | -0.10 (-0.65, 0.45) | -0.10 (-0.65, 0.45) | -0.17 (-0.71, 0.38) | 0.03 (-0.50, 0.55) |
| **Diabetes** | -0.28^***^ (-0.43, -0.13) | -0.28^***^ (-0.43, -0.13) | -0.22^***^ (-0.37, -0.08) | -0.30^***^ (-0.41, -0.18) |
| **Sex** | -3.40^**^ (-6.31, -0.50) | -3.40^**^ (-6.31, -0.50) | -0.48 (-3.37, 2.41) | -3.12^*^ (-6.35, 0.10) |
| **Age^2^** | 0.001 (-0.01, 0.01) | 0.001 (-0.01, 0.01) | 0.002 (-0.01, 0.01) | -0.002 (-0.01, 0.01) |
| **Age^3^** | -0.0000 (-0.0001, 0.0001) | -0.0000 (-0.0001, 0.0001) | -0.0000 (-0.0001, 0.0000) | 0.0000 (-0.0000, 0.0001) |
| **DBP** | -0.01^***^ (-0.01, -0.002) | -0.01^***^ (-0.01, -0.002) | -0.003^*^ (-0.01, 0.001) | -0.01^***^ (-0.01, -0.003) |
| **TDI** | -0.01 (-0.02, 0.002) | -0.01 (-0.02, 0.002) | -0.01^*^ (-0.02, 0.001) | -0.01^*^ (-0.02, 0.0003) |
| **BMI** | -0.01^***^ (-0.02, -0.004) | -0.01^***^ (-0.02, -0.004) | -0.02^***^ (-0.03, -0.01) | -0.01^***^ (-0.02, -0.004) |
| **Non-HDL cholesterol** | 0.05^***^ (0.02, 0.07) | 0.05^***^ (0.02, 0.07) | 0.03^*^ (-0.001, 0.05) | 0.01 (-0.02, 0.03) |
| **Site 1** | -0.16^***^ (-0.24, -0.09) | -0.16^***^ (-0.24, -0.09) | -0.14^***^ (-0.22, -0.07) | -0.13^***^ (-0.21, -0.06) |
| **Site 2** | -0.13^***^ (-0.19, -0.06) | -0.13^***^ (-0.19, -0.06) | -0.11^***^ (-0.17, -0.04) | -0.13^***^ (-0.19, -0.07) |
| **Exercise** | -0.0000 (-0.0004, 0.0003) | -0.0000 (-0.0004, 0.0003) | -0.0000 (-0.0004, 0.0003) | -0.0001 (-0.0004, 0.0002) |
| **Head size** | 2.42^***^ (2.13, 2.70) | 2.42^***^ (2.13, 2.70) | 2.55^***^ (2.26, 2.84) | 2.99^***^ (2.71, 3.26) |
| **Previous vs. never smoker** | -0.12^***^ (-0.18, -0.06) | -0.12^***^ (-0.18, -0.06) | -0.01 (-0.06, 0.05) | -0.06^**^ (-0.11, -0.003) |
| **Current vs. never smoker** | -0.18^***^ (-0.30, -0.06) | -0.18^***^ (-0.30, -0.06) | -0.20^***^ (-0.31, -0.09) | -0.10^**^ (-0.19, -0.02) |
| **Higher degree vs. no qualification** | -0.004 (-0.14, 0.13) | -0.004 (-0.14, 0.13) | -0.07 (-0.21, 0.07) | -0.07 (-0.20, 0.05) |
| **A levels vs. qualification** | 0.002 (-0.15, 0.15) | 0.002 (-0.15, 0.15) | -0.09 (-0.24, 0.07) | -0.07 (-0.21, 0.06) |
| **GCSEs vs. no qualification** | 0.02 (-0.13, 0.16) | 0.02 (-0.13, 0.16) | -0.06 (-0.21, 0.09) | -0.05 (-0.18, 0.08) |
| **CSEs vs. no qualification** | 0.09 (-0.13, 0.30) | 0.09 (-0.13, 0.30) | 0.09 (-0.11, 0.29) | -0.05 (-0.23, 0.12) |
| **NVQ vs. no qualification** | -0.04 (-0.21, 0.14) | -0.04 (-0.21, 0.14) | -0.15^*^ (-0.33, 0.02) | 0.0001 (-0.15, 0.15) |
| **Professional qualification vs. no qualification** | 0.10 (-0.07, 0.28) | 0.10 (-0.07, 0.28) | 0.01 (-0.17, 0.20) | 0.16^*^ (-0.02, 0.33) |
| **Age x sex** | 0.10^*^ (-0.01, 0.21) | 0.10^*^ (-0.01, 0.21) | 0.001 (-0.11, 0.11) | 0.11^*^ (-0.01, 0.23) |
| **Age^2^ x sex** | -0.001 (-0.002, 0.0003) | -0.001 (-0.002, 0.0003) | 0.0001 (-0.001, 0.001) | -0.001^*^ (-0.002, 0.0001) |
| **Constant** | 2.38 (-7.39, 12.16) | 2.38 (-7.39, 12.16) | 3.44 (-6.32, 13.19) | 0.01 (-9.39, 9.41) |
| **N** | 2,879 | 2,879 | 2,898 | 2,911 |
| **R^2^** | 0.48 | 0.48 | 0.49 | 0.52 |
| **Adjusted R^2^** | 0.48 | 0.48 | 0.48 | 0.52 |
| **Residual Std. Error** | 0.72 (df = 2849) | 0.72 (df = 2849) | 0.72 (df = 2868) | 0.70 (df = 2881) |
| **F Statistic** | 91.62^***^ (df = 29; 2849) | 91.62^***^ (df = 29; 2849) | 94.08^***^ (df = 29; 2868) | 108.18^***^ (df = 29; 2881) |
| ^*^p < .1; ^**^p < .05; ^***^p < .01 | | | | |

**Table 9: Regression models examining binging frequency in predicting grey matter volume, independently of total alcohol intake consumed.**

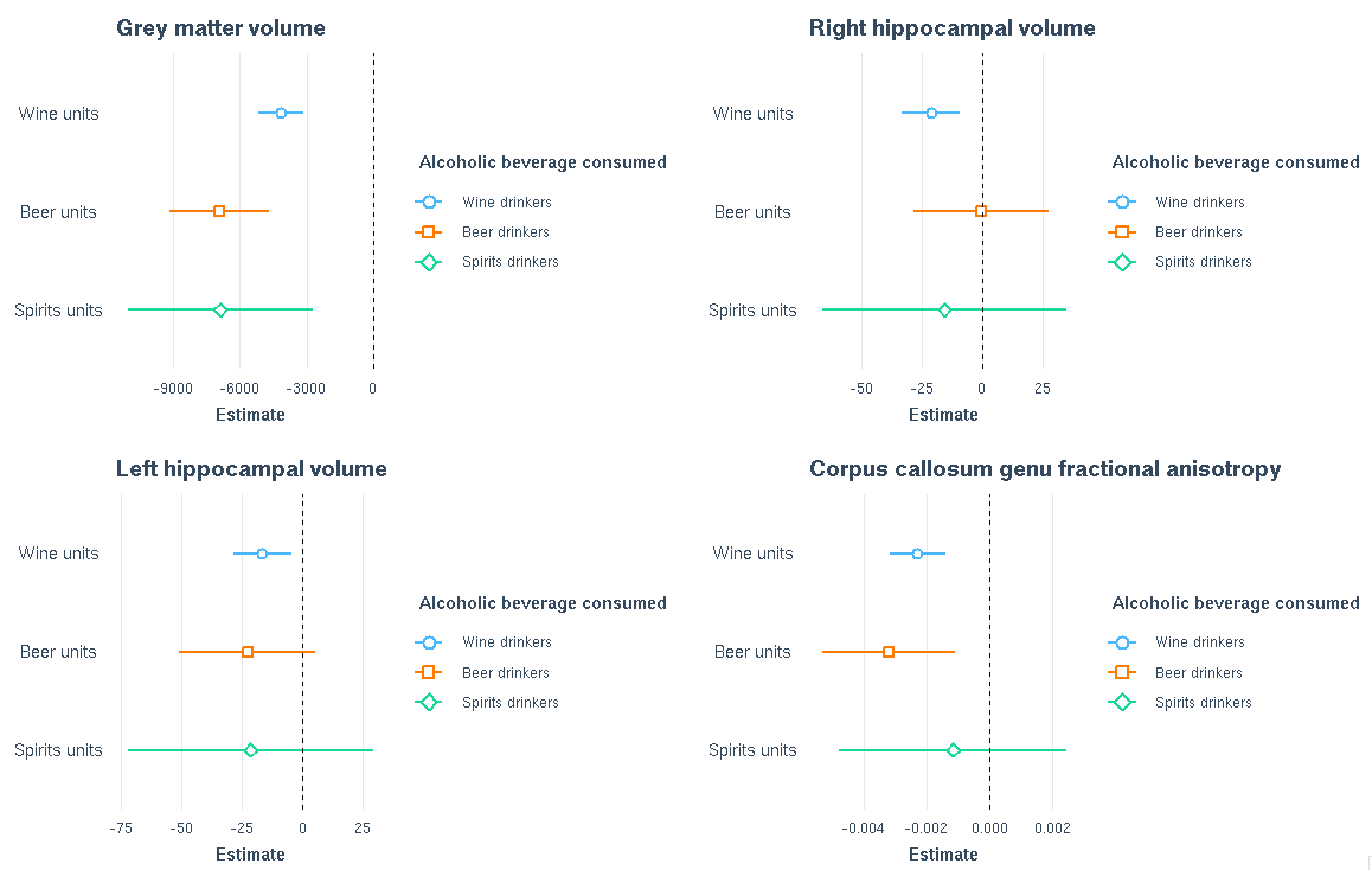

**Figure 16: Associations between weekly units of alcohol intake and selected brain measures for each alcoholic beverage type consumed. Subjects included are those solely drink one alcoholic beverage type. Coloured bars show regression coefficients (estimates) and 95% CI separately for wine (n=5080), beer (n=1193) and spirits (n=329) drinkers. Models were adjusted for: age, sex, age*sex, age^2^, age^3^, age^2^*sex, qualifications, SBP, DBP, Diabetes Mellitus, smoking status, TDI, BMI, cholesterol, HDL, imaging site, head size and exercise.**

**Node 3: DMN/attention/salience/CEN (inferior parietal)**

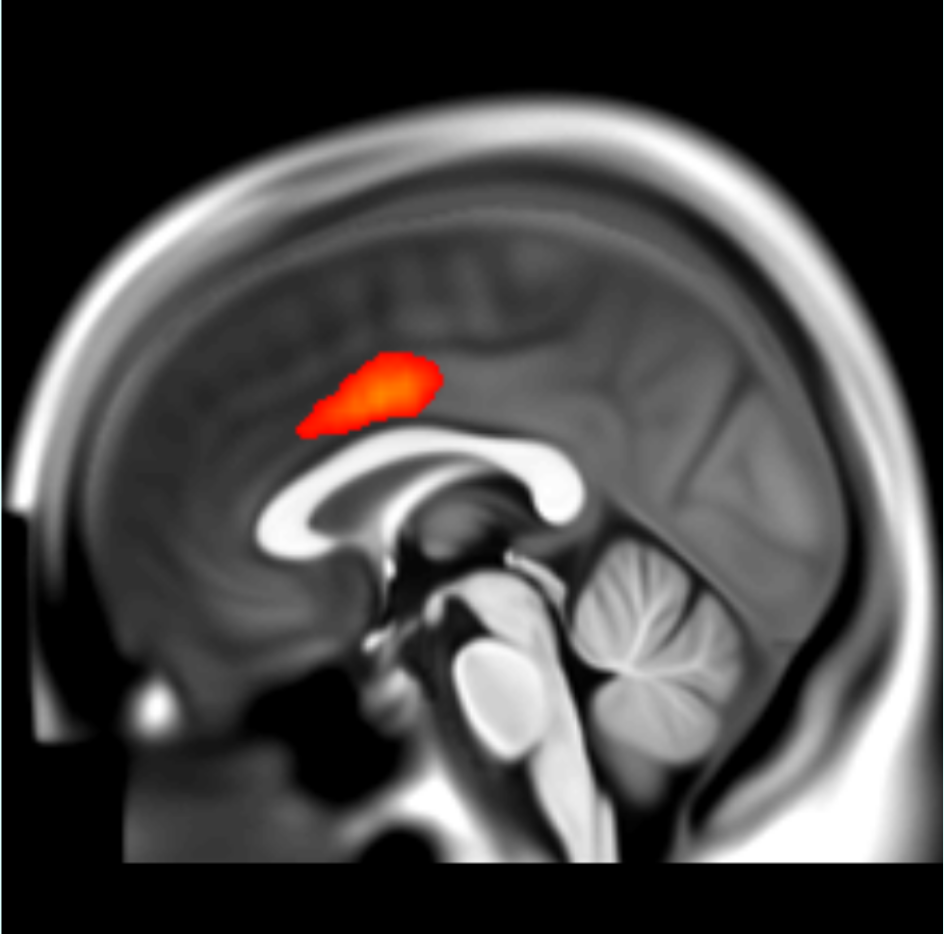

**Node 4: visual (lingual gyrus)**

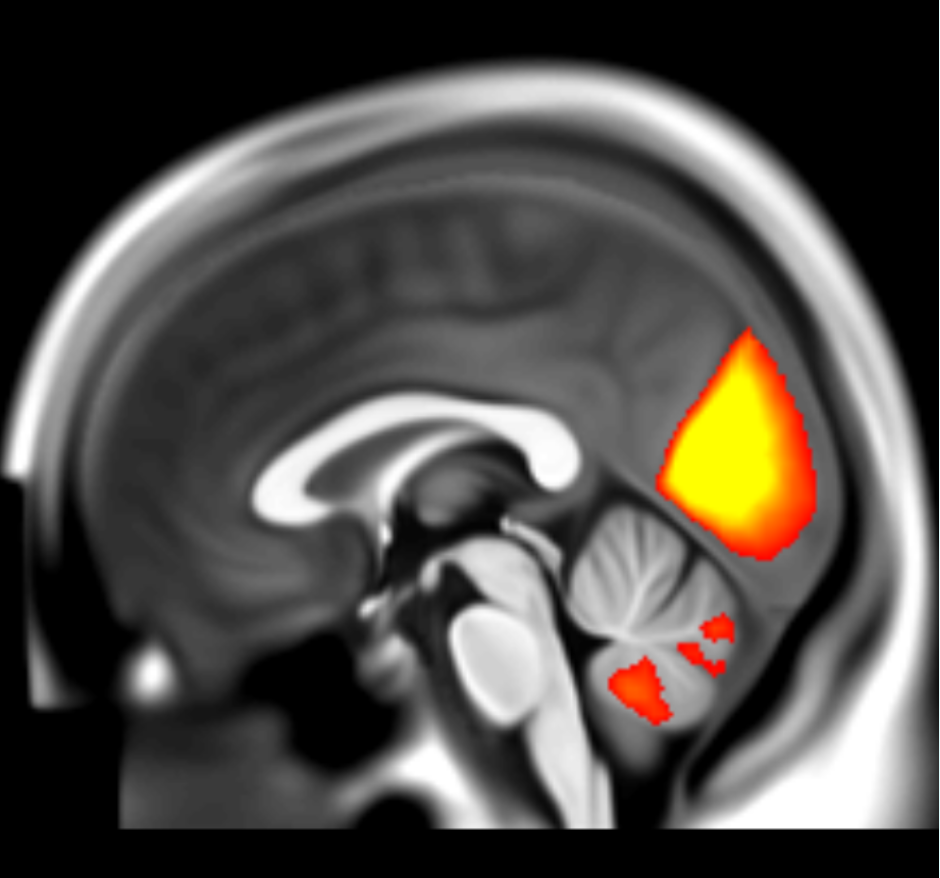

**Node 5: DMN (inferior parietal), CEN, attention**

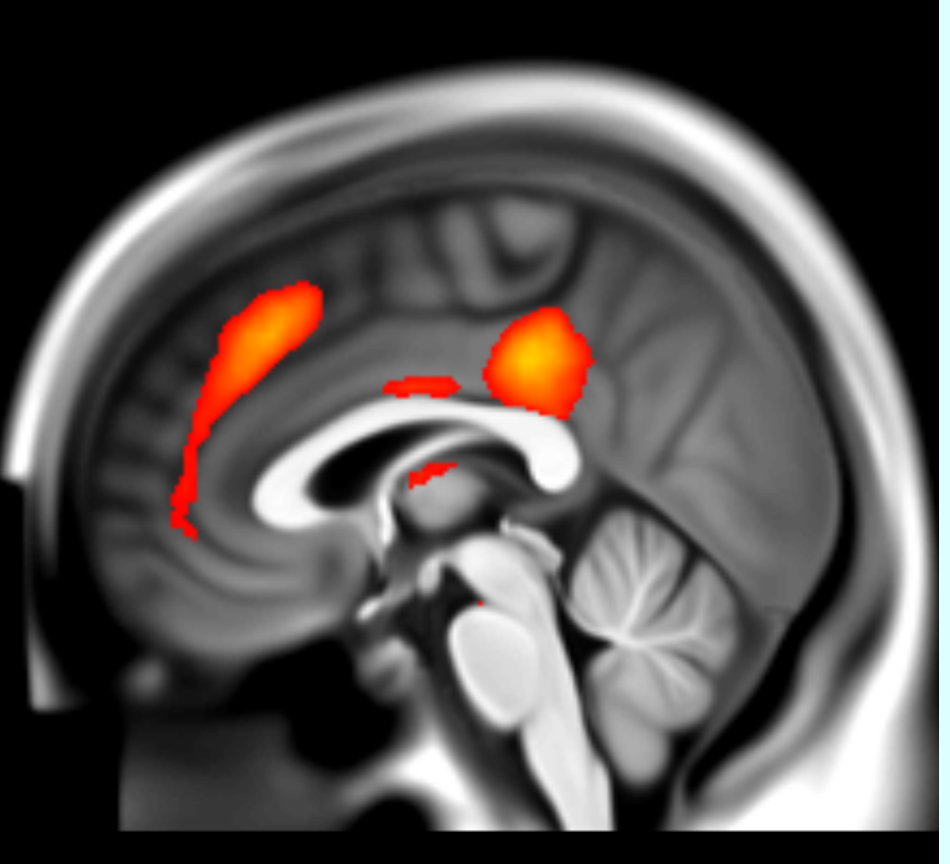

**Node 6: DMN/CEN (inferior and middle frontal)**

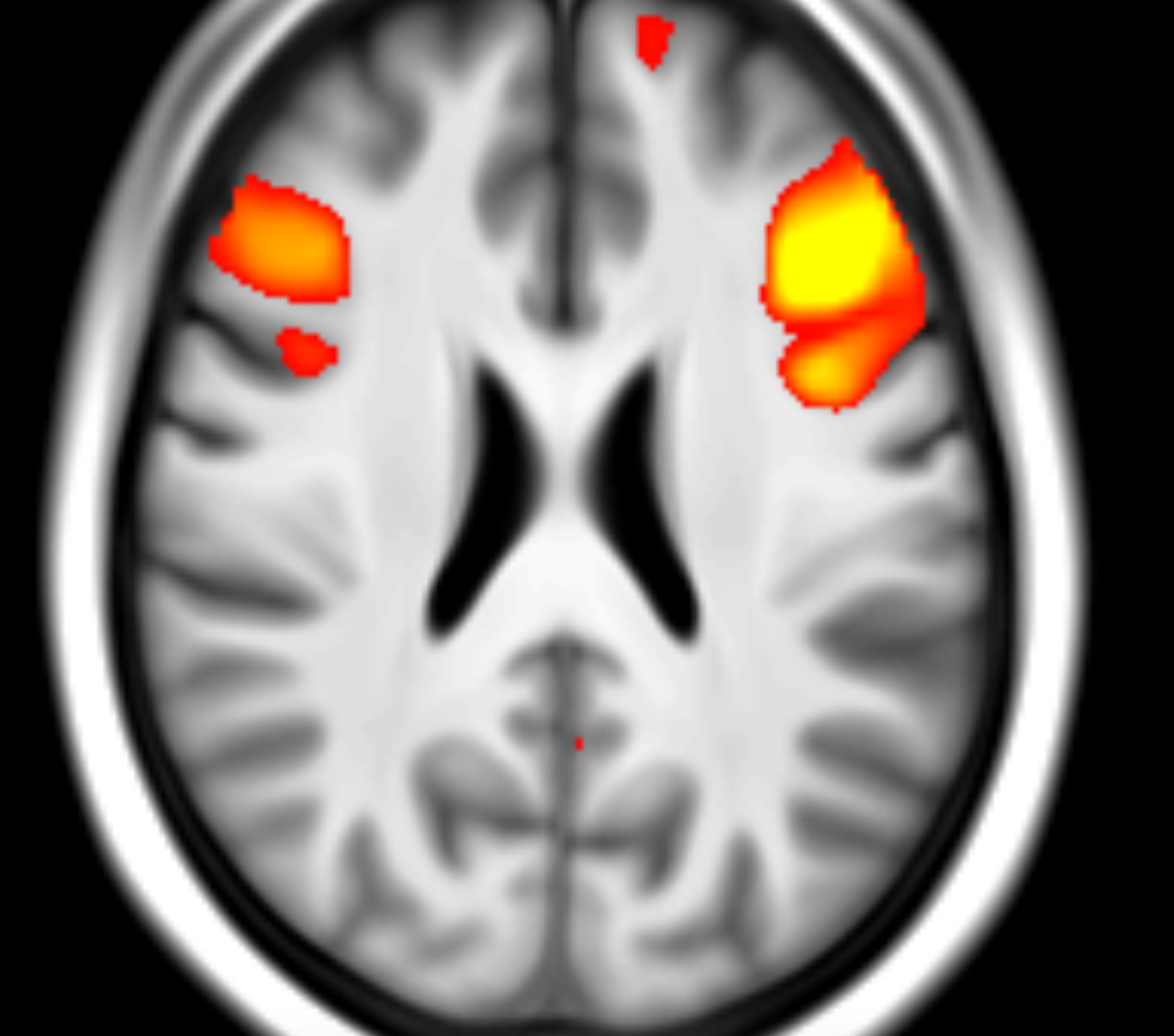

**Node 9: DMN/CEN**

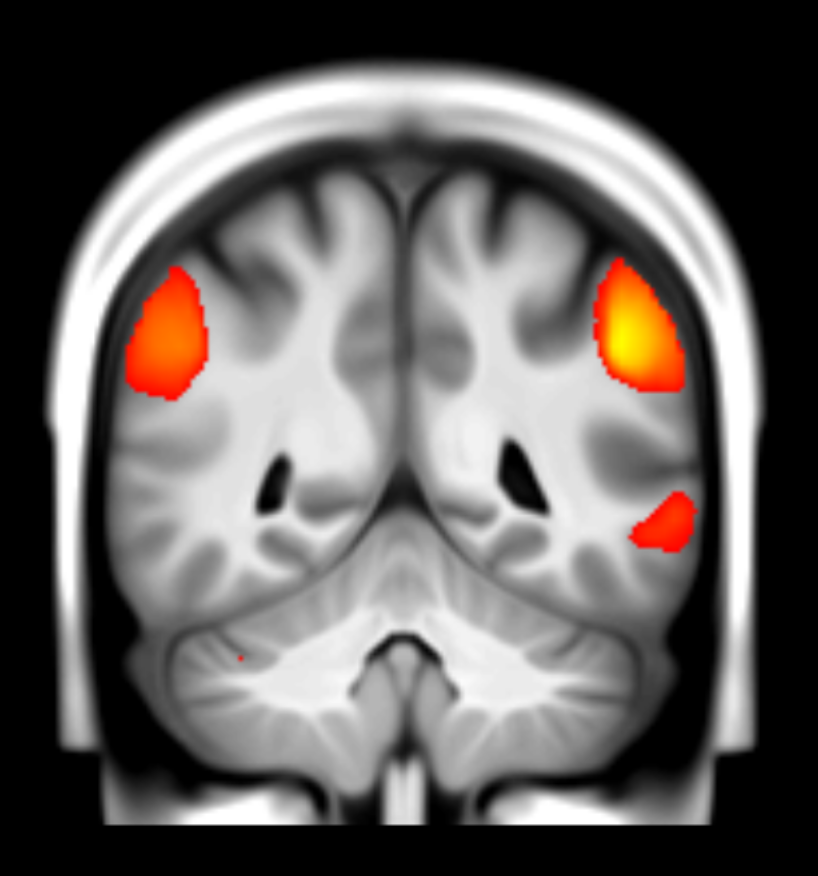

**Node 13: DMN/salience**

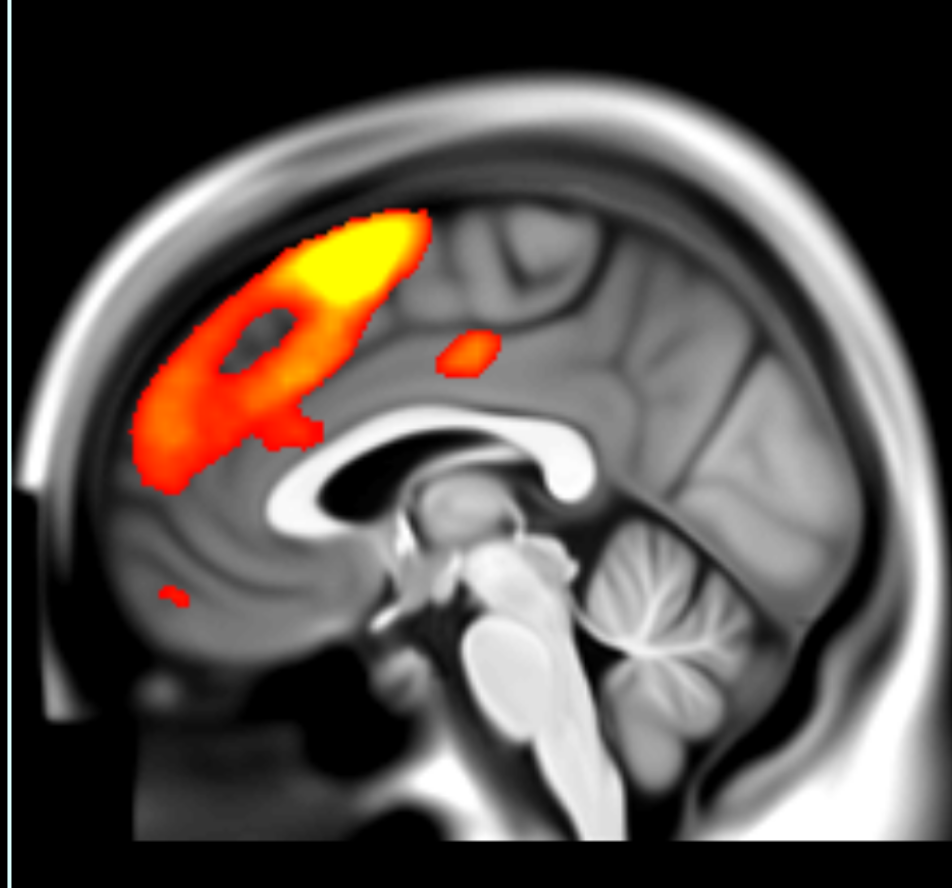

**Node 21: DMN/CEN**

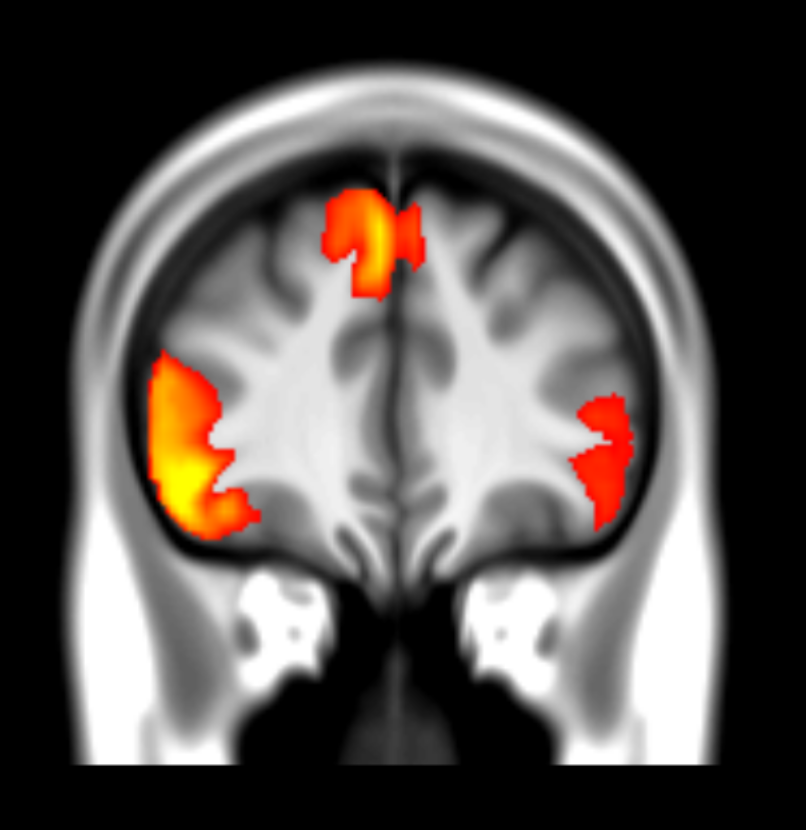

**Figure 17: Group average spatial maps for the resting state fMRI nodes significantly associated with alcohol intake. 3D versions of these maps can be visualized here: https://www.fmrib.ox.ac.uk/ukbiobank/group_means/rfMRI_ICA_d25_good_nodes.html)**

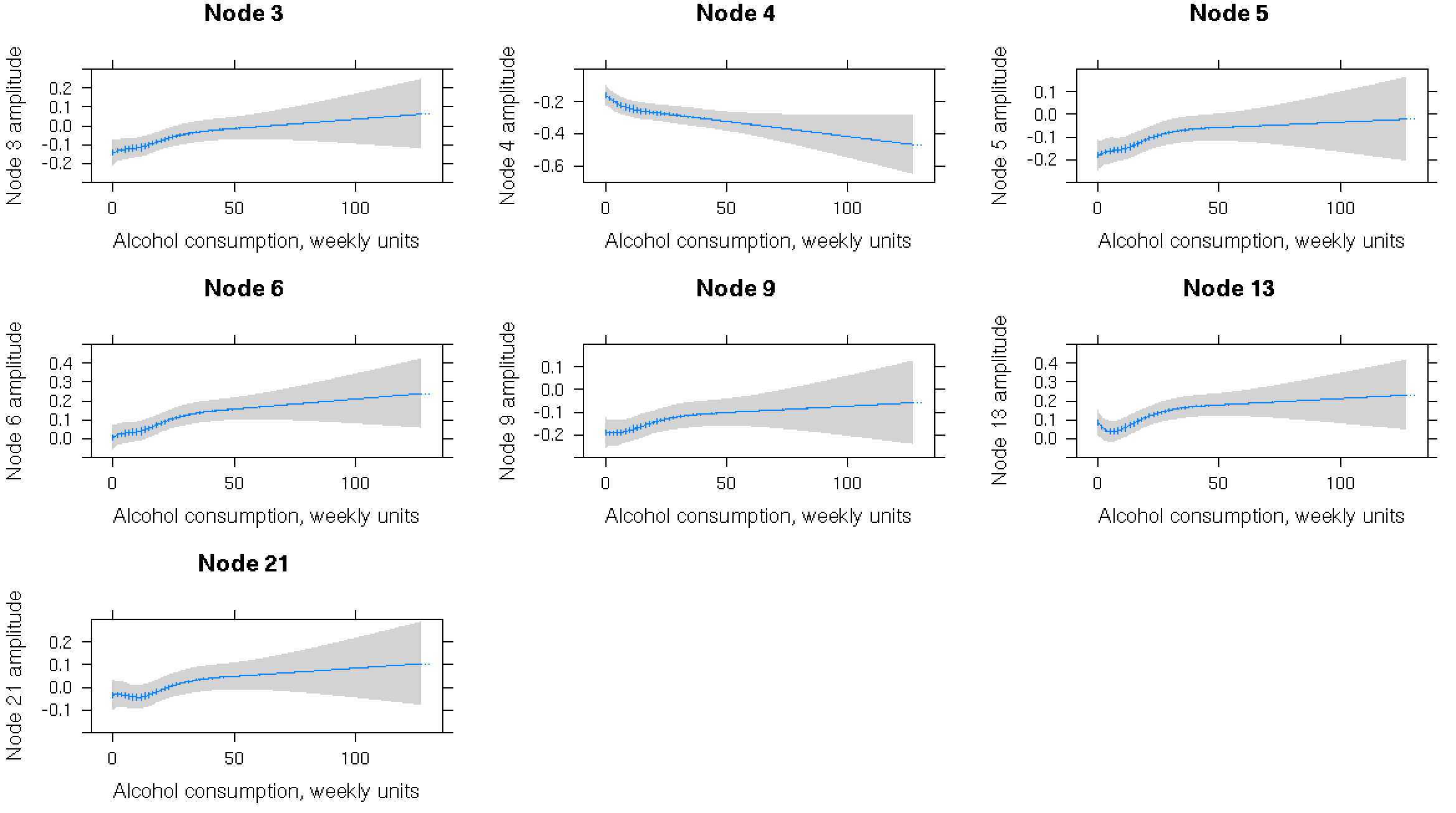

**Figure 18: Associations between alcohol intake weekly (units shown, grams conversion: 50u=400g, 100u=800g) and resting state fMRI node amplitude (standarised). Only significant (defined by Bonferroni corrected p<0.002) associations are shown. Spline fit with 5 knots to model alcohol intake flexibly. Models adjusted for: age, sex, age^2^, age^3^, age*sex, age^2^*sex, SBP, DBP, Diabetes Mellitus, smoking status, TDI, BMI, cholesterol, HDL, imaging site, head size, head motion and exercise. 95% CI are shaded. N=17,587.**

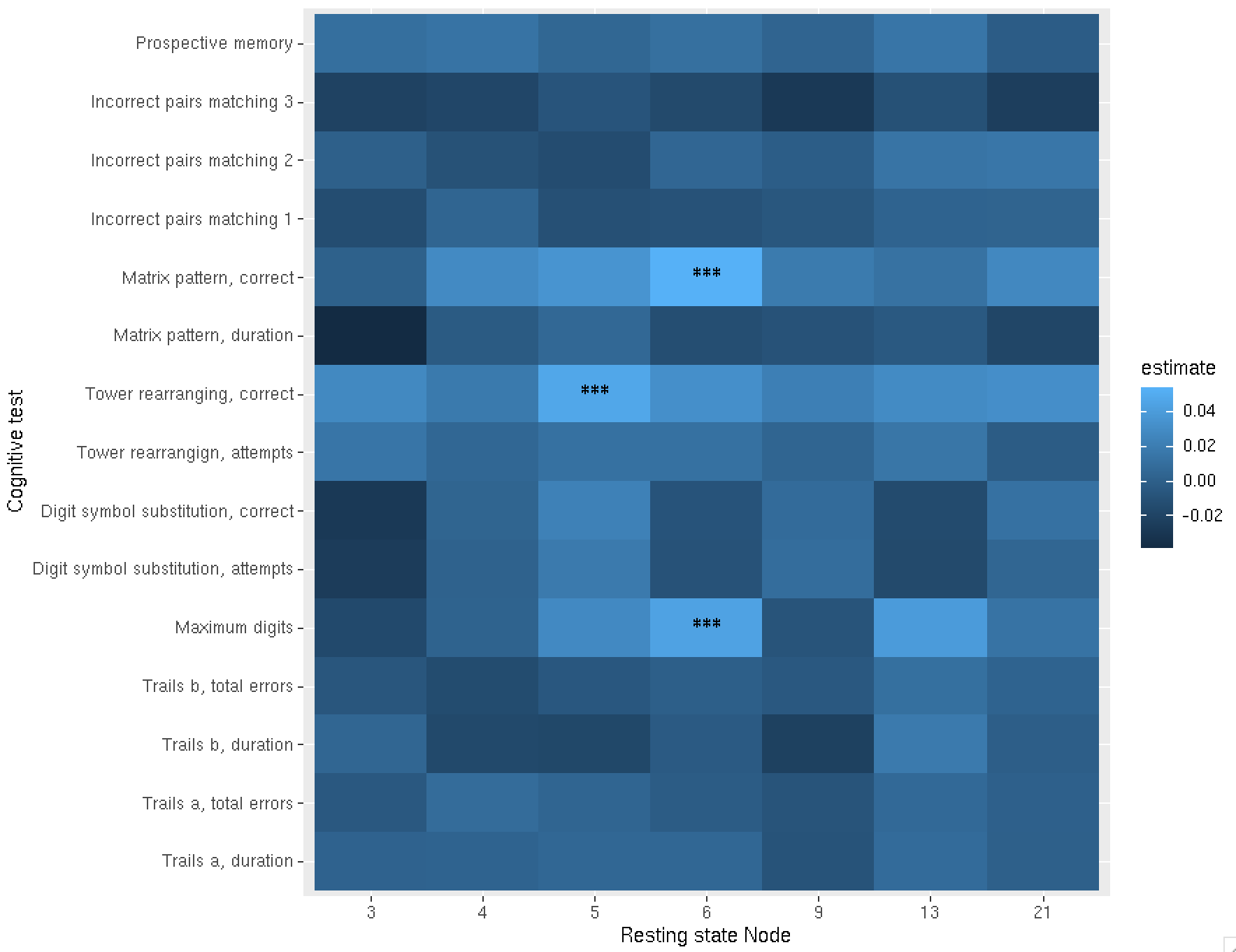

**Figure 19: Heatmap showing estimates (standardized regression coefficients) for associations between resting state node functional connectivity (node amplitudes significantly associated with alcohol consumption in supplementary Figure 18) and cognitive tests performed at the imaging visit. Estimates generated from regression models adjusted for: age, sex, age*sex, age^2^, age^2^*sex, SBP, DBP, Diabetes Mellitus, smoking status, TDI, BMI, cholesterol, HDL, imaging site, head size, head motion, qualifications and exercise. Stars represent Bonferroni significant p values (<0.0048). N=7086 subjects included.**

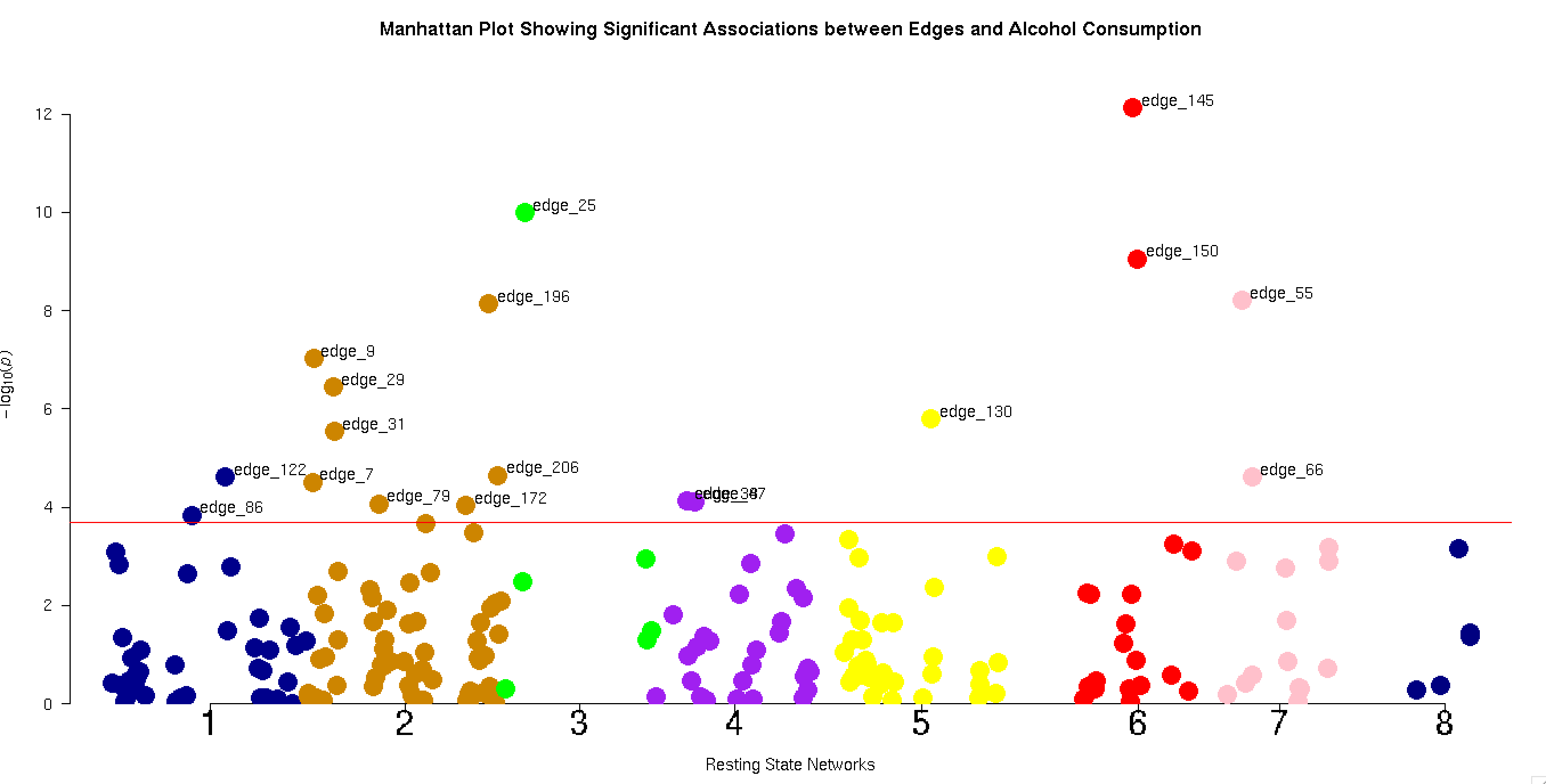

**Figure 20: Manhattan plot showing –log_10_(p) values for regression models with edge functional connectivity as the dependent variable and alcohol intake as an independent variable, adjusted for: age, sex, age*sex, age^2^, age^2^*sex, SBP, DBP, Diabetes Mellitus, smoking status, TDI, BMI, cholesterol, HDL, imaging site, head size, head motion and exercise. Bonferroni threshold is shown by the red line (p<0.0002) and edges passing this threshold are labelled. N=17,587. Edges are represented by dots, coloured according to the resting state networks their compromising nodes reside in: blue (1) – default mode ⬄ visual/attention, orange (2) – DMN ⬄ CEN, green (3) = visual ⬄visual, purple (4) – visual ⬄ CEN/attention/salience/motor/subcortical-cerebellum, yellow (5) – DMN ⬄ motor, red (6) – DMN ⬄ subcortical-cerebellum, pink (7) – motor ⬄ attention/salience/CEN/motor/subcortical-cerebellum, dark blue (8) – subcortical-cerebellum ⬄ attention/salience/CEN.**

| Edge | Nodes connected | Regions | Networks |
| --- | --- | --- | --- |
| 145 | 9-18 | Inferior parietal, angular, middle temporal ⬄ putamen/caudate/SMA | DMN/CEN ⬄ subcortical-cerebellum |
| 25 | 4-8 | Calcarine/lingual/cuneus ⬄ lingual/calcarine/superior occipital | Visual ⬄ visual |
| 150 | 14-18 | Anterior cingulate/superior frontal ⬄putamen/caudate/SMA | DMN/salience/CEN ⬄ subcortical-cerebellum |
| 55 | 10-11 | Postcentral/precentral ⬄ postcentral/precentral | Motor ⬄ motor |
| 195 | 5-21 | Inferior parietal/cerebellum/angular ⬄ inferior frontal/superior frontal/middle temporal | CEN/attention/DMN ⬄ DMN/CEN |

**Table 10: Top five most significantly associated edges (as defined by p values) with alcohol intake, together with the regions and networks of their constituent nodes.**

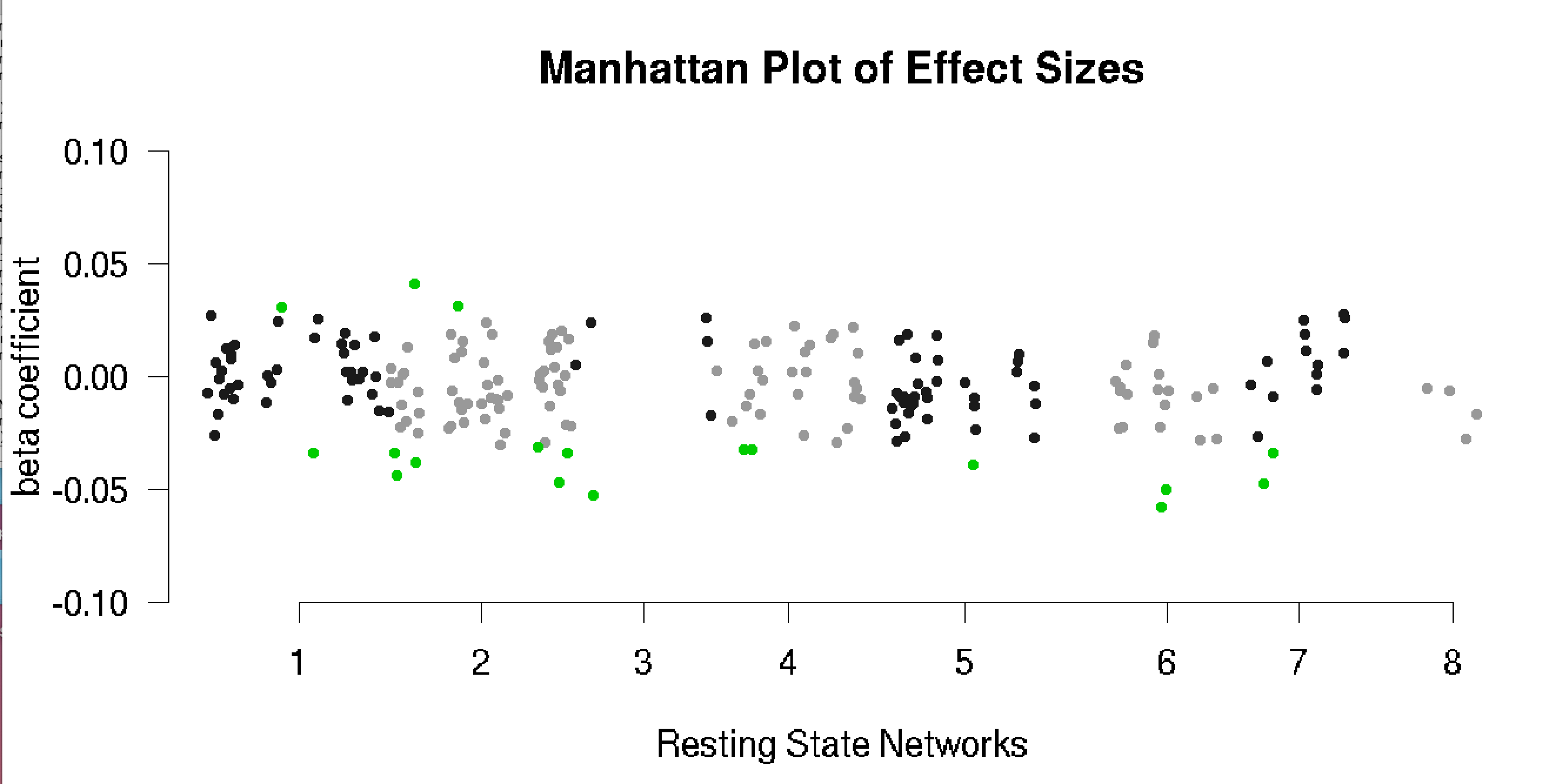

**Figure 21: Manhattan plot of beta coefficients from regression of alcohol intake onto edge functional connectivity. Dots represent edges (grouped by resting state network as in main Figure 12). Green coloured dots signify those edges whose association with alcohol reached Bonferroni significance (p<0.0002). Estimates generated using regression models with edge functional connectivity as the dependent variable and alcohol intake as an independent variable, adjusted for: age, sex, age*sex, age^2^, age^2^*sex, SBP, DBP, Diabetes Mellitus, smoking status, TDI, BMI, cholesterol, HDL, imaging site, head size, head motion and exercise.**
